# Mid-life leukocyte telomere length and dementia risk: a prospective cohort study of 435,046 UK Biobank participants

**DOI:** 10.1101/2022.06.30.22277121

**Authors:** Rui Liu, Meiruo Xiang, Luke C. Pilling, David Melzer, Lihong Wang, Kevin J. Manning, David C. Steffens, Jack Bowden, Richard H. Fortinsky, George A. Kuchel, Taeho G Rhee, Breno S. Diniz, Chia-Ling Kuo

## Abstract

**Importance:** Telomere attribution is one of biological aging hallmarks and may be intervened to target multiple aging-related diseases, including Alzheimer’s disease and Alzheimer’s disease related dementias (AD/ADRD).

**Objective:** To assess associations of leukocyte telomere length (TL) with AD/ADRD and early markers of AD/ADRD, including cognitive performance and brain magnetic resonance imaging (MRI) phenotypes.

**Design, Setting, and Participants:** Data from European-ancestry participants in the UK Biobank (n=435,046) were used to evaluate whether mid-life leukocyte TL is associated with incident AD/ADRD over a mean follow-up of 12.2 years. In a subsample without AD/ADRD and with brain imaging data (n=43,390), we associated TL with brain MRI phenotypes related to AD or vascular dementia pathology.

**Exposures:** Relative mean leukocyte TL at baseline was measured in T/S ratio, using a multiplex quantitative polymerase chain reaction technique by comparing the amount of the telomere amplification product (T) to that of a single-copy gene (S).

**Main Outcomes and Measures:** AD/ADRD was confirmed by mapping multi-source data to 3-charcter ICD codes (G30; F00; F01; F03; G31), excluding other-cause dementias (F02).

**Results:** Longer TL was associated with a lower risk of incident AD/ADRD (adjusted HR [aHR] per SD=0.93, 95% CI 0.90 to 0.96, *P*=3.37×10^−7^). Longer TL also was associated with better cognitive performance in specific cognitive domains, larger hippocampus volume (adjusted standardized *β* [a*β*] per SD=0.012, 95% CI 0.003 to 0.021, *P*=0.012), lower total volume of white matter hyperintensities (a*β* per SD= -0.015, 95% CI -0.024 to -0.006, *P*=0.002), and higher fractional anisotropy (FA; a*β* per SD=0.018, 95% CI 0.009 to 0.027, *P*=9.44×10^−5^) and lower mean diffusivity (MD; a*β* per SD=0.045, 95% CI 0.035 to 0.056, *P*=1.07×10^−17^) in the fornix.

**Conclusions and Relevance:** Longer TL is a protective factor against AD/ADRD, cognitive impairment, and brain structural lesions toward the development of AD/ADRD. A better understanding of underlying mechanisms may help improve diagnosis and management of AD/ADRD in older adults.

**Key Points:** *Question:* Is leukocyte telomere length (TL) associated with Alzheimer’s disease (AD) and AD related dementias (AD/ADRD) and early markers of AD/ADRD, including cognitive performance and brain magnetic resonance imaging phenotypes?

*Findings:* Longer TL was associated with a lower risk of incident AD/ADRD, larger hippocampus volume, lower total volume of white matter hyperintensities, higher fractional anisotropy and lower mean diffusivity in the fornix.

*Meaning:* Longer TL in midlife may play a protective role against AD/ADRD, and a better understanding of underlying mechanisms may help improve diagnosis and management of AD/ADRD in older adults.

## Introduction

Telomeres are repetitive sequences at the end of chromosomes, where they protect DNA from damage and preserving genome stability.^1^ In somatic cells, telomeres shorten with each cell division and are replenished by the telomerase enzyme activity.^2^ Critically short telomere length (TL) signals cells to stop replicating and can trigger cellular senescence changes.^3^ A significant consequence of cellular senescence is the change in the cellular secretome and a shift towards a pro-inflammatory state (i.e., the Senescence-Associated Secretory Phenotype, SASP) that can exert deleterious effects in different tissues and organs, including the brain.^4^ Shorter leukocyte TL is associated with increased mortality risks^5^ and cardiovascular disease.^6^ In contrast, longer leukocyte TL may increase risks of certain cancers, including glioma, ovarian, and lung cancer.^7^

Recent studies suggest that TL may also play an important role in the development of neurodegeneration and neurodegenerative disorders.^8^ Alzheimer’s disease (AD) is the most prevalent neurodegenerative disease associated with aging. Previous conflicting associations between TL and AD^9^ may be explained by small samples, pathology of other neurodegenerative disorders in AD patients, and controls including possible preclinical AD cases. Due to shared pathological features, AD, frontotemporal, Lewy body, vascular, and mixed dementia are classified as AD and AD-related dementias (AD/ADRD) for research purposes.^10^ Cognitive decline and brain changes occur many years before AD diagnosis.^11^ Recent meta-analyses reported that longer TL was associated with better general cognition^12^ and performance in several cognitive domains.^13,14^ Few population-based studies have explored TL in relation to brain MRI imaging features.^14–16^ A recent meta-analysis showed that longer leukocyte TL is associated with whole brain and hippocampus volumes but not with white matter hyperintensities (WMH).^17^

We leveraged data from a large prospective cohort in the UK Biobank (UKB) to not only estimate associations between TL and risk of incident AD/ADRD, but to also assess associations with early markers of AD/ADRD, including cognitive performance and brain imaging derived phenotypes (IDPs)^18,19^ in participants who were dementia-free at the time of multi-modality imaging assessment. As a means of avoiding confounding, we also carried out Mendelian randomization (MR) analyses to estimate causal effects of genetically determined TL on AD/ADRD and related measures.

## Methods

### UK Biobank

UKB is a volunteer community cohort recruiting over 500,000 volunteers aged 40 to 69 years between 2006 and 2010.^20,21^ At recruitment, participants completed an extensive questionnaire and provided biological samples for genetic and other future assays. Since 2014, UKB re-invited participants to undergo a multimodal imaging assessment of the brain, heart, and body. During the visit, baseline and additional cognitive tests were administered online.

### Inclusion and Exclusion Criteria

Data were from active UKB participants. We excluded from our analysis participants with 1) non-European ancestry (n=51,131) to avoid confounding from ancestry differences; 2) extreme TL <0.01^st^ or >99.9^th^ percentile (n=15,644); 3) diagnosed with AD/ADRD prior to or at baseline (n=393); 4) diagnosed with other cause-dementia at any time before the last follow-up (n=190) (Figure 1). The baseline cohort (N=435,046) was used to assess associations of TL with incident AD/ADRD and baseline cognitive performance. A subsample who attended the first imaging visit between 2014 and 2019 and were free of AD/ADRD, termed “imaging cohort”, was used to test for associations of TL with IDPs and performance of cognitive tests first implemented at the first imaging visit. For specific analyses, participants with any missing outcome or covariates were further excluded.

**Figure 1.**
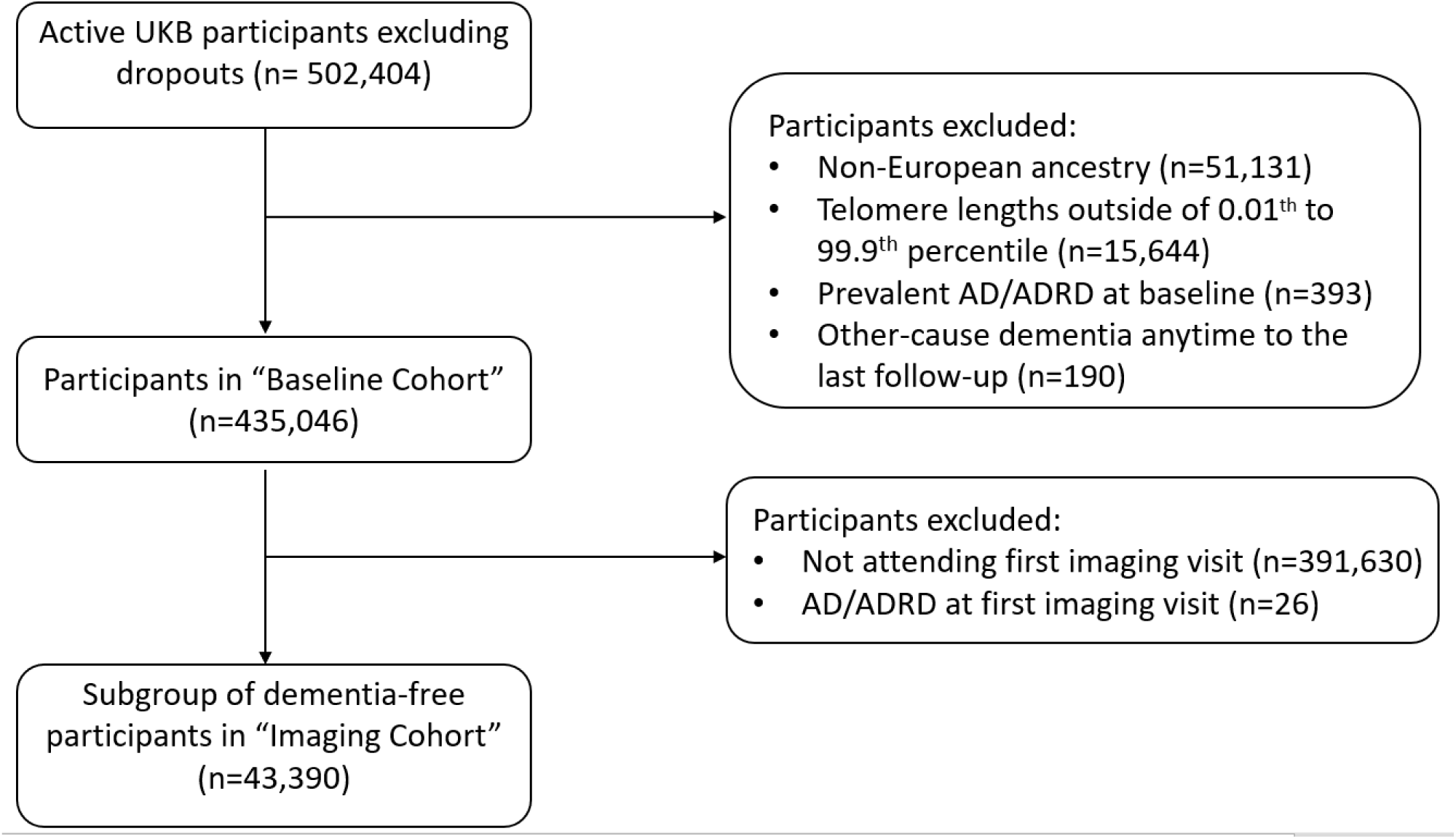
Flowchart of study participant selection.

## Data

### Telomere Length Measurement

Relative mean TL was measured from peripheral blood leukocytes in T/S ratio using a multiplex quantitative polymerase chain reaction technique in T/S ratio by comparing the amount of the telomere amplification product (T) to that of a single-copy gene (S).^22^ TL was adjusted for the influence of technical parameters and was log- and z-transformed among the included samples before analysis.

### Genotype Data

UKB participants were genotyped using DNA extracted from the baseline blood samples.^20^ Genetic principal components (PCs) to account for ancestry differences were derived within participants of European descent using the genome-wide genotype data. *APOE* genotypes were determined based on the genotypes of the two single nucleotide polymorphisms (SNPs), rs429358 and rs7412, on chromosome 19. Genetic variants strongly associated with leukocyte TL at the genome-wide significance level (p<8.31×10^−9^) in the UKB GWAS^23^ were selected as genetic instruments (n=130) (eTable 1 in Supplement 1) to estimate TL-outcome associations using MR methods. These genetic variants were uncorrelated and robust to pleiotropy, and they were enriched with functional variants near genes underlying telomere and telomerase biology.^23^

### Assessment of Outcomes

AD/ADRD was confirmed using the first occurrence data of UKB by mapping multi-source data to 3-charcter ICD codes (eTable 1 in Supplement 2). A dementia case was first diagnosed with AD/ADRD or other-cause dementia (ICD-10: F02) depending on which occurred first. Other-cause dementias as described previously were excluded from our analyses. AD and vascular dementia (VD) as subtypes of AD/ADRD were analyzed, allowing the cases to have other diagnoses in AD/ADRD, e.g., a patient may have both AD and VD diagnoses.

We selected UKB cognitive tests *a priori*, previously shown to have moderate to high concurrent validity with well-validated reference tests and test-retest reliability (specifically, reaction time, numeric memory, symbol digit substitution, trail making part B, and matrix pattern completion).^24^ We also included tests mostly correlated with general cognitive ability, i.e., fluid intelligence,^24^ and derived general cognitive ability scores combining five baseline cognitive test scores via the first principal component from Fawns-Ritchie et al.^24^ Data were from baseline or the first imaging visit depending on when the cognitive test was first implemented. Cognitive assessments are detailed in eMethods 1 in Supplement 3. Their associated cognitive domains, data field IDs, and weblinks are provided in eTable 2 in Supplement 2.

We utilized the IDPs generated by an image-processing pipeline developed and run on behalf of UKB.^25^ Specifically, we selected IDPs associated with dementia^18,19^: 1) AD-signature region volumes from T1-weighted structural imaging: hippocampus, parahippocampal cortex, entorhinal cortex, inferior parietal lobule, precuneus, and cuneus; 2) total volume of WMH derived from combined T1 and T2-weighted fluid-attenuated inversion recovery (FLAIR) structural imaging; and 3) weighted mean fractional anisotropy (FA) and mean diffusivity (MD) of white matter tracts from diffusion-weighted imaging. Left and right hemisphere measurements for the same tract were averaged before analysis. A description of the selected IDPs is provided in eTable 3 in Supplement 2.

### Covariates

Baseline covariates included demographics (age, sex, assessment center near residence), socioeconomic status (education, Townsend deprivation index), lifestyle factors (body mass index [BMI], smoking status, alcohol intake frequency, physical activity), *APOE* genotype, and top 10 genetic PCs (PC1-PC10). Townsend deprivation index score was a measure of material deprivation at the postcode level based on the preceding national census data. Higher scores represent greater levels of deprivation. Smoking status and alcohol intake frequency were accessed through a touchscreen questionnaire. Physical activity (low, moderate, or high) was self-reported and measured following the short International Physical Activity Questionnaire guidelines.^26^

### Statistical Methods

The association between TL and time from baseline to first incident AD/ADRD diagnosis was estimated using a Cox proportional hazards model. Participants who did not develop AD/ADRD but died during follow-up were censored at date of death; otherwise, at the last follow-up date (March 31, 2021). Sensitivity analyses were performed by *APOE* genotype (e3e3, e4 [e3e4 or e4e4], or e2 [e2e3 or e2e2]), for AD and VD, separately. Linear regression models were used to examine the associations of TL with cognitive measures and IDPs. Before modeling, each continuous outcome was transformed by the rank-based inverse normal transformation to correct for the distributional skewness, followed by a z-transformation to unify the scales. Each of the above models adjusting for all as described in “Covariates” (full model [primary]). Unadjusted results (unadjusted model) and results adjusted for age and sex only (base model) are presented for comparison. IDPs were adjusted for head size additionally regardless of the models.

To estimate the causal effects of TL on AD/ADRD and related outcomes, we applied several MR methods to ensure our results are robust to MR assumptions: 1) inverse-variance weighting (IVW) method^27^ (primary) that meta-analyzes causal estimates from individual genetic instruments; 2) a weighted median-based method^28^ that assumes that the majority of genetic variants are valid instrumental variables; 3) MR-Egger^29^ that allows us to assess horizontal pleiotropy additionally; 4) robust adjusted profile score (MR-RAPS) method,^30^ accounting for any residual weak instrument bias, pleiotropy, and extreme outliers. Both instrument-TL associations and instrument-outcome associations from the present study were adjusted for age, sex, genotyping array and PC1-PC10, plus head size additionally for IDPs.

Prior to the MR analysis, the instrument-TL associations (regression coefficients and standard errors) were adjusted for winner’s curse using (see the method in eMethods 2 and R code in eMethods 3 in Supplement 3). Before and after winner’s curse adjustment, we calculated the mean F-statistic^31^ and percent of variance in TL attributed to the genetic instruments to evaluate the weak instrument bias, and the I^2^-statistic^32^ to evaluate the no measurement error (NOME) assumption of MR-Egger.

Observational p-values or MR p-values from the primary IVW method were evaluated at the level of false discovery rate less than 5%. All the statistical analyses were performed in R version 3.4.1.

## Results

Participant characteristics at recruitment of the baseline cohort (n=435,046) are presented in eTable 4 in Supplement 2. The mean follow-up time from baseline to first imaging visit for participants in the imaging cohort (n=43,390) was 8.97 years (SD=1.75). Participants who subsequently underwent the first imaging visit were healthier than the baseline sample (eTable 4 in Supplement 2), which had also been shown by a previous study.^33^ In the baseline cohort, incident AD/ADRD cases and normal controls were compared for TL and covariates one at a time (eTable 5 in Supplement 2).

### Observational Association Analysis

During a mean follow-up of 12.2 years, we identified 6,424 incident AD/ADRD cases (mean age at diagnosis 72.8 years, SD=6.06): 2,527 AD cases (mean age at diagnosis 73.5 years, SD=5.09) and 1,330 VD cases (mean age at diagnosis 73.5 years, SD=5.03). The incidence of AD/ADRD or its subtype linearly decreased as TL increased (eFigure 1 in Supplement 2). Compared with normal controls (0.83±0.13), AD/ADRD cases (0.80±0.12, *P*<2.2×10^−16^) had a shorter mean TL and similarly, AD (0.80±0.12, *P*<2.2×10^−16^) and VD (0.79±0.12, *P*<2.2×10^−16^) cases.

Next, we present the full model results adjusted for a full set of covariates, which are similar to the base model results adjusted for age and sex only. In contrast, the unadjusted models tended to show larger effect sizes, which may be of the same or opposite direction. Longer leukocyte TL at baseline was associated with a lower risk of incident AD/ADRD (Figure 2). The adjusted hazard ratio (aHR) of AD/ADRD was 0.93 per SD longer in TL (95% CI 0.90 to 0.96, *P*=3.37×10^−7^). Similar results were found for different *APOE* genotypes, as well as for VD and AD (Figure 2). One SD of TL corresponds to approximately 650 base pairs in a European adult population, which is approximately 26 years of telomere attrition given the telomere shortening rate in the population is about 25 base pairs per year.^34^

**Figure 2.**
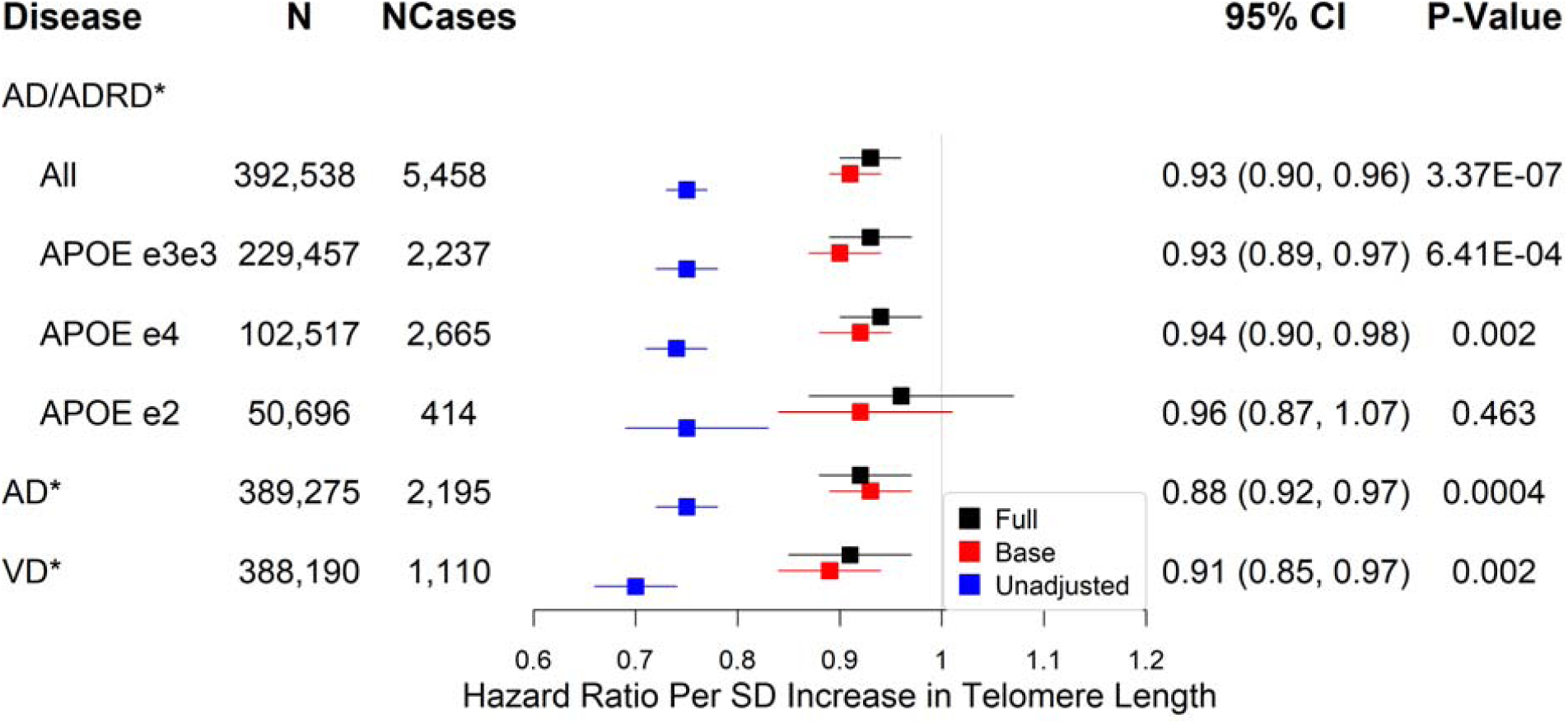
Associations between telomere length and AD/ADRD or related phenotypes. *Significant at the false discovery rate < 0.05 level; AD: Alzheimer’s disease (AD) or dementia in AD; VD: vascular dementia; N (full model): sample size; NCases (full model): number of cases; Full: Cox proportional hazards models adjusting for age, sex, education, Townsend deprivation index, BMI, smoking status, alcohol intake frequency, IPAQ physical activity group, *APOE* genotype, PC1-PC10, and baseline assessment center; Base: Cox proportional hazards models adjusting for age and sex only; Unadjusted: Cox regression models with no adjustment for covariates; P-Value: unadjusted p-value for multiple testing.

Longer TL was significantly associated with better cognitive performance at baseline, including faster reaction time (adjusted standardized *β* [a*β*] per SD= -0.005, 95% CI -0.008 to - 0.002, *P*=0.003), higher fluid intelligence (a*β* per SD=0.011, 95% CI 0.006 to 0.017, *P*=1.41×10^−5^), and higher numeric memory (a*β* per SD=0.013, 95% CI 0.003 to 0.023, *P*=0.009). Although not statistically significant at the level of FDR<5%, longer TL was suggestive of higher general cognitive ability (a*β* per SD=0.009, 95% CI -0.001 to 0.018, *P*=0.066) (Figure 3). For cognitive tests measured at the first imaging visit, each SD increase in TL was associated with better scores on tests of processing speed/executive functioning, including more symbol digit substitution (a*β* per SD=0.012, 95% CI 0.001 to 0.023, *P*=0.029) and shorter duration to complete trail making part B (a*β* per SD= -0.013, 95% CI -0.024 to -0.002, *P*=0.022). Additionally, each SD increase in TL was associated with better non-verbal reasoning measured with matrix pattern completion (a*β* per SD=0.013, 95% CI 0.001 to 0.024, *P*=0.027).

**Figure 3.**
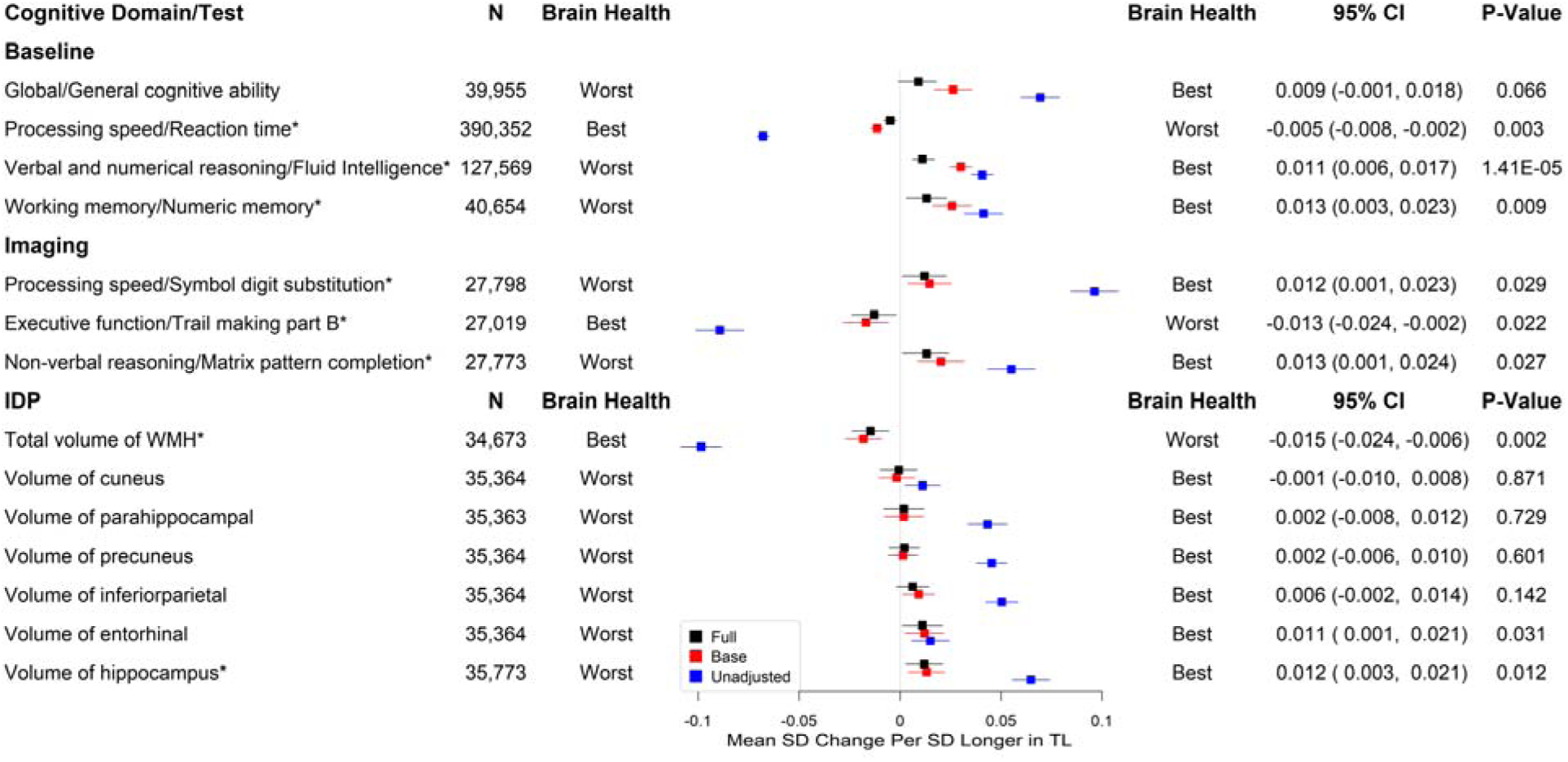
Associations between telomere length and cognitive function measures. *Significant at the false discovery rate <0.05 level; N (full model): sample size; Full: linear regression models adjusting for age, sex, education, Townsend deprivation index, BMI, smoking status, alcohol intake frequency, IPAQ physical activity group, *APOE* genotype, PC1-PC10, and baseline assessment center; Base: linear regression models adjusting for age and sex only; Unadjusted: linear regression models with no adjustment for covariates; Head size was included in each of the models for IDPs; P-Value: unadjusted p-value for multiple testing; The two ends representing best and worst brain health are labelled.

Longer TL was significantly associated with higher hippocampus volume (a*β* per SD=0.012, 95% CI 0.003 to 0.021, *P*=0.012) and lower total WMH volume (a*β* per SD= -0.015, 95% CI -0.024 to -0.006, *P*=0.002). There were no significant associations between TL and other AD signature volumes (Figure 3). We also evaluated the associations between TL and markers of white matter microstructural damage assessed by diffusion tensor imaging. Longer TL was associated with higher FA (indicating better white matter fiber integrity) and lower MD (indicating more glial cellularity and lower inflammatory burden) in the fornix. Interestingly, most of the other white matter tracts analyzed showed the opposite direction of association between TL and FA or MD. (Figures 4 and 5).

**Figure 4.**
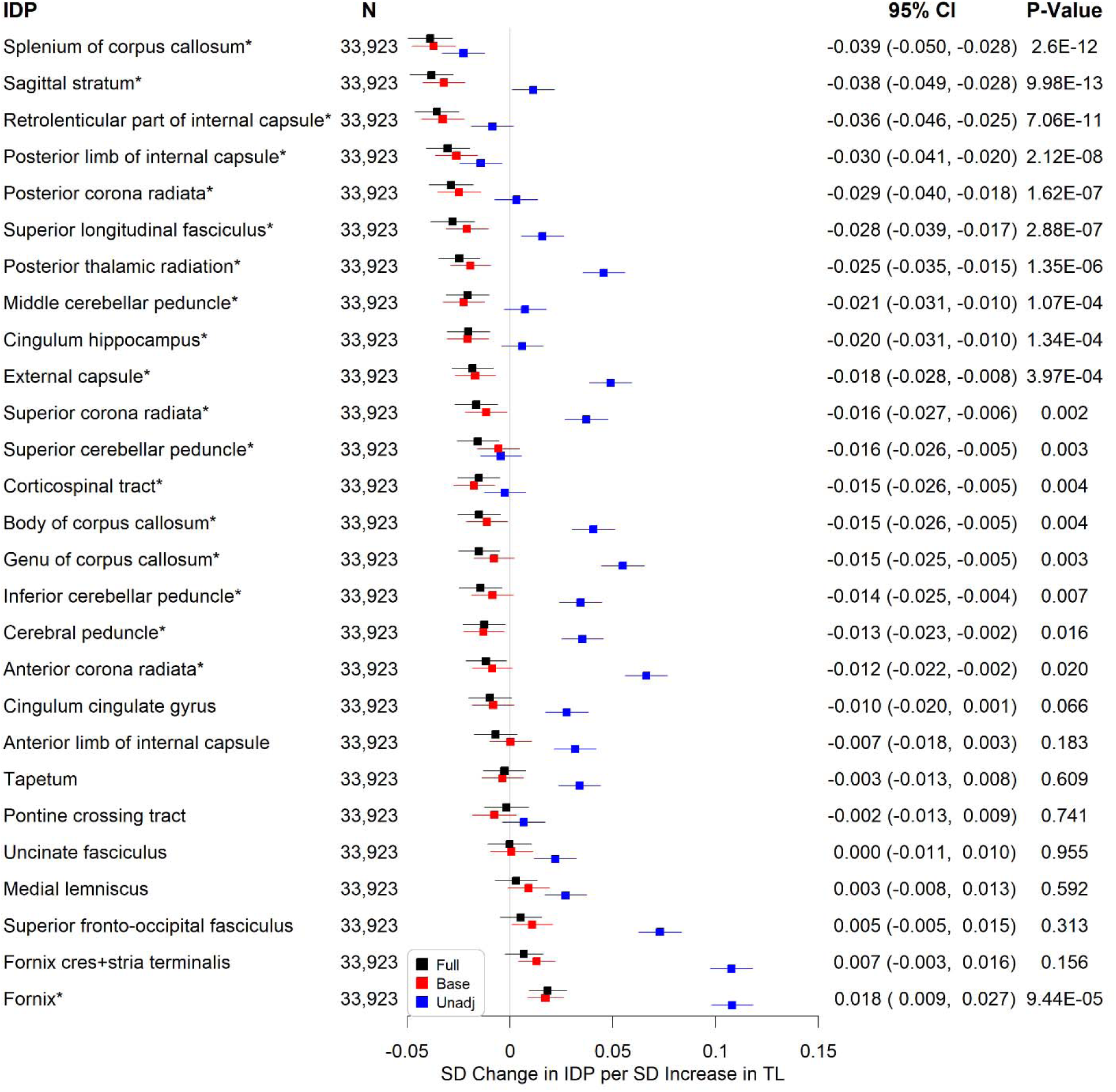
Associations between telomere length and weighted-mean fractional anisotropy IDPs. *Significant at the false discovery rate < 0.05 level; N (full model): sample size; Full: linear regression models adjusting for age, sex, education, Townsend deprivation index, BMI, smoking status, alcohol intake frequency, IPAQ physical activity group, *APOE* genotype, PC1-PC10, baseline assessment center, and head size; Base: linear regression models adjusting for age, sex, and head size only; Unadjusted: linear regression models adjusting for head size only; P-Value: unadjusted p-value for multiple testing.

**Figure 5.**
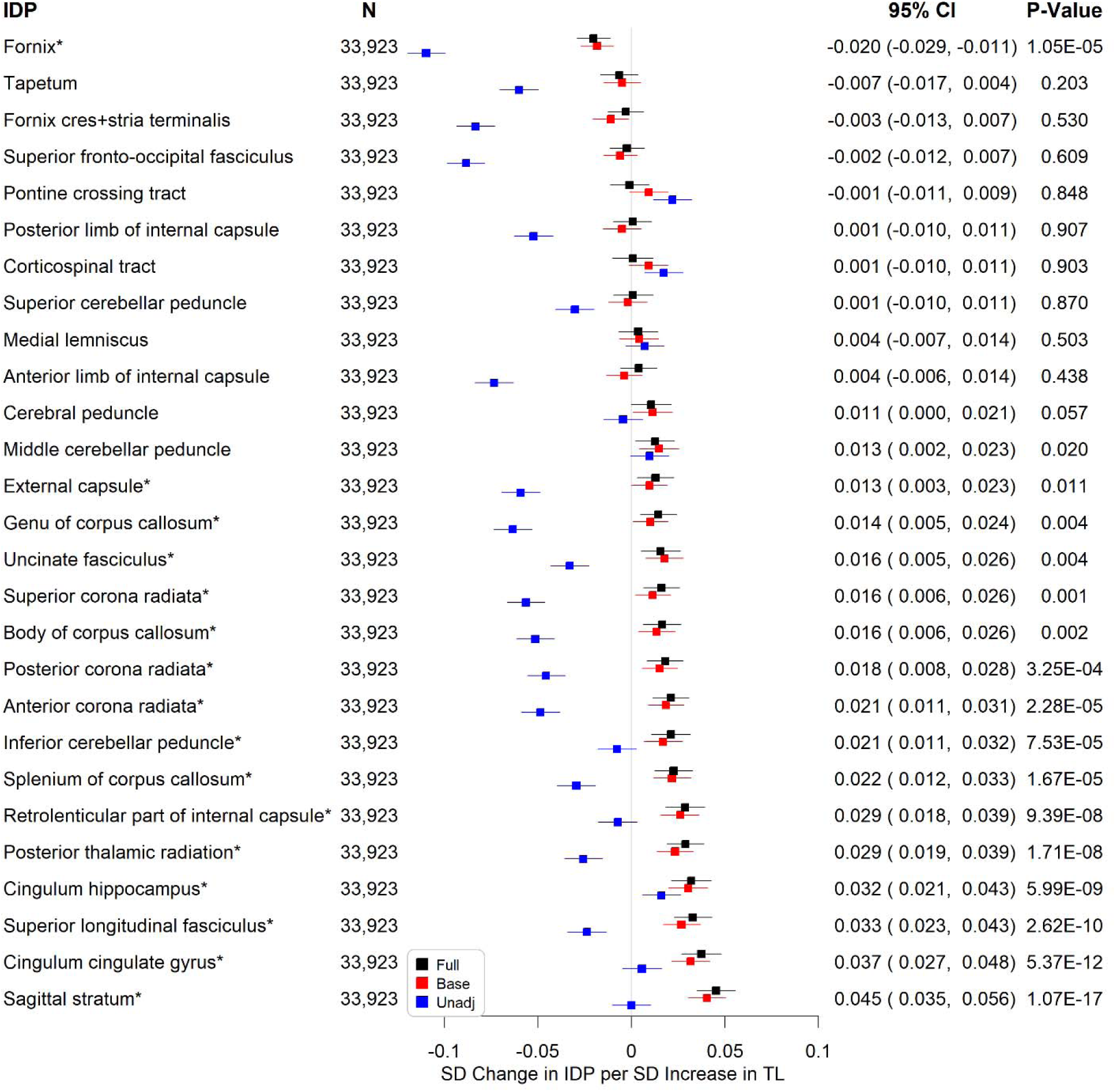
Associations between telomere length and weighted-mean mean diffusivity IDPs. *Significant at the false discovery rate < 0.05 level; N (full model): sample size; Full: linear regression models adjusting for age, sex, education, Townsend deprivation index, BMI, smoking status, alcohol intake frequency, IPAQ physical activity group, *APOE* genotype, PC1-PC10, baseline assessment center, and head size; Base: linear regression models adjusting for age, sex, and head size only; Unadjusted: linear regression models adjusting for head size only; P-Value: unadjusted p-value for multiple testing.

### Mendelian Randomization Analysis

A single instrument (rs35671754) was dropped from the analysis after winner’s curse correction due to its sign reversing. The corrected effect sizes and standard errors were presented in eTable 1 in Supplement 1. Our instrument set remained strong after winner’s curse correction (mean F-statistic 127.11 [before] versus 120.25 [after]; percent of the variance explained: 3.61% [before] versus 3.42% [after]). There was a low risk of violating the NOME assumption of MR-Egger, with the I^2^ statistic close to 1 before (97.6%) and after (97.4%) winner’s curse adjustment.

Using the IVW MR method, there was no evidence showing that genetically determined TL was associated with AD/ADRD or its subtypes (eFigure 2 in Supplement 2). Genetically determined TL was not associated with any of the cognitive measures (eFigure 3 in Supplement 2), volumetric IDPs of AD signatures, or total volume of WMH (eFigure 4 in Supplement 2). Genetically determined TL was not associated with FA or MD in the fornix. The MR analysis, however, confirmed observational associations of longer TL with lower FA (eFigure 5 in Supplement 2), and higher MD (eFigure 6 in Supplement 2) in several tracts. Different MR methods showed consistent results (eFigures 7-11 in Supplement 2). MR-Egger tended to produce a larger effect size than the other methods, which may be inflated by horizontal pleiotropy (indicated by an estimated intercept significantly different from zero; see eFigures 1-71 in Supplement 4). MR-RAPS accounted for pleiotropy and the results of IVW and MR-RAPS were similar across outcomes.

## Discussion

Using middle-aged adults with a mean follow-up of 12.2 years, we found that longer leukocyte TL was associated with a lower risk of AD/ADRD, and specifically of AD and VD. In the population without dementia diagnoses, longer TL was associated with better performance. We also found that longer TL was associated with brain MRI features linked to a lower risk of dementia, including higher hippocampus volume, lower total volume of WMH, along with higher FA and lower MD in the fornix.

Our results are consistent with previous observational studies suggesting a protective effect of longer TL on AD.^9^ Different previous MR studies,^35,36^ we did not find significant genetic associations of TL with AD/ADRD or its subtypes. Our samples were younger and we used more genetic instruments of TL to increase statistical power. However, the percent of variance in TL explained by the genetic instruments is still insufficient to power the outcomes for the observed effect sizes due to limited numbers of cases in this young cohort. Perhaps TL in brain tissues may show a stronger causal effect.

The relationship we observed between TL and alterations in the fornix is commensurate with the study by Staffaroni et al.^37^ that showed TL attrition over time was associated with decreased fornix FA, increased fornix MD, and greater hippocampal volume loss. The fornix is a white matter bundle in the limbic system that functions as the principal outflow pathway from the hippocampus.^38^ Microstructural changes in the fornix had been reported as an early predictor of cognitive decline in older adults with normal cognition and an indicator of AD progression.^39^ Moreover, reduced FA and increased MD in the fornix have been suggested as promising imaging markers for AD.^38^ Our findings on associations of longer TL with lower FA and higher MD in several white matter tracts suggest that longer TL may also have deleterious effects on related health outcomes including dementia.

The mechanism linking leukocyte TL with AD/ADRD and related brain MRI markers is unclear. Telomere dysfunction has a major impact on stem cell exhaustion and genomic instability, where the biological changes including reduced neurogenesis and increased mosaic DNA content variation have been lined to AD.^40–42^ Additionally, shorter TL may reflect an increased pool of senescent cells, which is associated with SASP.^4^ Neuroinflammation plays an important role in exacerbating amyloid-*β* burden and tau hyperphosphorylation that are two core pathologies of AD.^43^ Pro-inflammatory cytokines and other SASP factors are associated with brain structural abnormalities, cerebrovascular pathology, cognitive impairment, and elevated risk of AD.^44,45^ Future studies are necessary to address by which mechanisms TL can have protective or harmful effects.

Several limitations need to be considered when interpreting the results of the current study. First, TL was measured in leukocytes in peripheral blood rather than in the brain. However, peripheral blood TL is positively correlated with cerebellum TL and plays a direct role in AD pathogenesis.^46^ Second, study participants were relatively young, with many not yet old enough to have developed AD/ADRD. Third, our results may not be generalized to non-European populations. Lastly, healthy volunteers are over-represented in our baseline and imaging cohorts, which could lead to underestimated exposure-outcome associations but the impact is alleviated by significant heterogeneity of exposures.^47^ Overall, we believe that the above limitations would diminish rather than enhance our ability to reject the null hypothesis, thus raising the robustness of our statistically significant findings.

Together with our findings that longer TL is associated with better cognitive performance and lower risks of incident AD, VD, and their related brain markers, this suggests that TL is a robust indicator of neurodegeneration or cognitive impairment toward the development of AD/ADRD. Further research is needed to elucidate the biological mechanisms linking TL and dementia and to understand the health impact of lower FA and higher MD associated with longer TL in several white matter tracts.

## Data Availability

All data produced in the present work are contained in the manuscript.

## Acknowledgements

This research was conducted using the UK Biobank resources under application no. 14631 and was funded by the National Institute of Nursing Research, National Institutes of Health, USA (R21NR018963-01A1S1). K.J.M., D.C.S, R.H.F., G.A.K. and C.L.K. were supported by P30AG067988. All the authors have no conflict of interest to disclose.

## Author Contributions

C.L.K., L.C.P, D.M., L.W., D.C.S, and R.L. designed the study; M.X. and C.L.K. performed statistical analyses; J.B. guided Mendelian randomization analyses and developed the method to adjust for the winner’s curse; R.L. and C.L.K drafted the initial manuscript, and all the authors reviewed the manuscript.

