## Supplement 1 for "Mid-life leukocyte telomere length and dementia risk: a prospective cohort study of 435,046 UK Biobank participants"

**eTable 1 Genetic intruments selected for Mendelian randomization analyses**

Modified table based on Supplementary Table 1 of Codd et al. (Nature Genetics 2021)

A1 is the effect allele,  A2 the other allele

N_imp is the sample size multiplied by the imputation quality to indicate approximate sample size for the test.

Beta is the estimated mean standard deviation change in leukocye telomere length per copy increase in the effect allele.

SE is the standard error of beta.

F=(Beta/SE)^2

%var explained' is the percent of variance explained by a particular genetic variant.

| | | | | | | | | Uncorrected for the winner's curse | | | | | Corrected for the winner's curse | | | |
|---|---|---|---|---|---|---|---|---|---|---|---|---|---|---|---|---|
| **Chr** | **Genetic Variant** | **BP_GRCh37** | **GENE** | **A1** | **A2** | **N_imp** | **FREQ (A1)** | **Beta** | **SE** | **Pvalue** | **F** | **%var explained** | **Beta** | **SE** | **F** | **%var explained** |
| 1 | rs187540244 | 11,224,327 | EXOSC10 | G | A | 455,136 | 0.995 | 0.091 | 0.015 | 1.2E-09 | 36.55 | 0.008 | 0.04 | 0.06 | 0.55 | 0.000 |
| 1 | rs66731853 | 20,916,238 | CDA | G | A | 463,432 | 0.683 | 0.018 | 0.002 | 1.5E-16 | 77.79 | 0.017 | 0.02 | 0.00 | 76.32 | 0.016 |
| 1 | rs17185038 | 28,219,658 | RPA2 | C | G | 464,716 | 0.935 | 0.026 | 0.004 | 2.1E-10 | 40.58 | 0.009 | 0.02 | 0.01 | 6.59 | 0.001 |
| 1 | rs6669563 | 32,279,629 | SPOCD1 | A | G | 464,716 | 0.562 | 0.018 | 0.002 | 2.1E-19 | 77.79 | 0.017 | 0.02 | 0.00 | 76.32 | 0.016 |
| 1 | 1:41236837_CT_C | 41,236,837 | NFYC | C | CT | 458,273 | 0.775 | 0.015 | 0.002 | 1.7E-09 | 42.68 | 0.009 | 0.01 | 0.00 | 11.31 | 0.002 |
| 1 | rs41269079 | 45,252,015 | BEST4 | A | T | 464,215 | 0.811 | 0.015 | 0.003 | 1.7E-09 | 34.57 | 0.007 | 0.00 | 0.01 | 0.00 | 0.000 |
| 1 | rs139795227 | 92,842,367 | RPAP2 | C | A | 437,231 | 0.986 | 0.06 | 0.009 | 6.7E-12 | 47.85 | 0.011 | 0.06 | 0.01 | 24.16 | 0.006 |
| 1 | rs4498805 | 110,910,397 | SLC16A4 | T | G | 464,158 | 0.453 | 0.015 | 0.002 | 5.7E-14 | 54.02 | 0.012 | 0.01 | 0.00 | 38.31 | 0.008 |
| 1 | rs3838300 | 114,442,355 | MAGI3 | C | CA | 463,050 | 0.820 | 0.033 | 0.003 | 6.1E-36 | 138.29 | 0.030 | 0.03 | 0.00 | 138.29 | 0.030 |
| 1 | rs11579626 | 146,741,960 | CHD1L | C | A | 464,433 | 0.915 | 0.027 | 0.004 | 1.3E-13 | 49.79 | 0.011 | 0.03 | 0.00 | 28.90 | 0.006 |
| 1 | rs61818036 | 151,364,199 | PSMB4 | G | A | 461,745 | 0.176 | 0.019 | 0.003 | 2.1E-12 | 55.47 | 0.012 | 0.02 | 0.00 | 41.71 | 0.009 |
| 1 | rs932002 | 226,577,306 | PARP1 | C | T | 463,856 | 0.849 | 0.04 | 0.003 | 7.3E-47 | 203.18 | 0.044 | 0.04 | 0.00 | 203.18 | 0.044 |
| 2 | rs9752694 | 17,874,177 | SMC6 | C | G | 461,608 | 0.571 | 0.014 | 0.002 | 2.7E-12 | 47.06 | 0.010 | 0.01 | 0.00 | 21.96 | 0.005 |
| 2 | rs56178008 | 29,098,543 | TRMT61B | A | T | 463,319 | 0.563 | 0.014 | 0.002 | 9.7E-13 | 47.06 | 0.010 | 0.01 | 0.00 | 21.96 | 0.005 |
| 2 | rs202034370 | 54,488,018 | | T | TA | 456,276 | 0.975 | 0.103 | 0.007 | 2.6E-56 | 241.15 | 0.053 | 0.10 | 0.01 | 241.15 | 0.053 |
| 2 | rs12613375 | 58,984,109 | LINC01122 | T | C | 461,376 | 0.862 | 0.018 | 0.003 | 9.1E-10 | 41.14 | 0.009 | 0.01 | 0.01 | 7.83 | 0.002 |
| 2 | rs775145631 | 210,667,432 | UNC80 | TC | T | 462,415 | 0.409 | 0.027 | 0.002 | 2.0E-41 | 175.03 | 0.038 | 0.03 | 0.00 | 175.03 | 0.038 |
| 2 | rs35671754 | 216,220,870 | ATIC | T | G | 462,765 | 0.704 | 0.012 | 0.002 | 1.4E-08 | 27.32 | 0.006 | 0.00 | 0.01 | 0.01 | 0.000 |
| 3 | rs869785 | 24,347,800 | THRB | T | C | 463,558 | 0.328 | 0.015 | 0.002 | 4.4E-12 | 54.02 | 0.012 | 0.01 | 0.00 | 38.31 | 0.008 |
| 3 | rs575032615 | 47,638,657 | SMARCC1 | G | A | 416,872 | 0.985 | 0.061 | 0.008 | 1.1E-12 | 52.50 | 0.013 | 0.06 | 0.01 | 35.54 | 0.009 |
| 3 | rs78491606 | 72,891,547 | SHQ1 | A | C | 464,716 | 0.982 | 0.076 | 0.007 | 1.9E-24 | 105.53 | 0.023 | 0.08 | 0.01 | 105.52 | 0.023 |
| 3 | rs6776756 | 128,215,821 | GATA2 | G | A | 463,785 | 0.402 | 0.017 | 0.002 | 1.1E-17 | 69.39 | 0.015 | 0.02 | 0.00 | 65.51 | 0.014 |
| 3 | 3:138398778_TA_T | 138,398,778 | PIK3CB | T | TA | 454,254 | 0.441 | 0.015 | 0.002 | 9.1E-13 | 54.02 | 0.012 | 0.01 | 0.00 | 38.31 | 0.008 |
| 3 | rs41272947 | 160,119,525 | | G | A | 464,361 | 0.458 | 0.017 | 0.002 | 5.9E-18 | 69.39 | 0.015 | 0.02 | 0.00 | 65.51 | 0.014 |
| 3 | rs2293607 | 169,482,335 | TERC | T | C | 464,283 | 0.758 | 0.094 | 0.002 | <1.0E-314 | 1676.20 | 0.360 | 0.09 | 0.00 | 1676.20 | 0.360 |
| 4 | rs753936006 | 2,191,750 | POLN | AAAA | C | 461,329 | 0.886 | 0.025 | 0.003 | 1.1E-14 | 66.69 | 0.014 | 0.02 | 0.00 | 62.09 | 0.013 |
| 4 | rs871134 | 7,044,380 | CCDC96 | C | T | 462,176 | 0.431 | 0.018 | 0.002 | 1.7E-19 | 77.79 | 0.017 | 0.02 | 0.00 | 76.32 | 0.017 |
| 4 | rs13129697 | 9,926,967 | SLC2A9 | G | T | 464,716 | 0.721 | 0.017 | 0.002 | 1.5E-14 | 54.82 | 0.012 | 0.02 | 0.00 | 40.20 | 0.009 |
| 4 | rs35500378 | 122,729,413 | EXOSC9 | ACTT | C | 463,888 | 0.611 | 0.014 | 0.002 | 2.0E-12 | 47.06 | 0.010 | 0.01 | 0.00 | 21.96 | 0.005 |
| 4 | rs4435700 | 164,020,174 | NAF1 | A | C | 459,000 | 0.235 | 0.054 | 0.002 | 2.2E-112 | 553.17 | 0.120 | 0.05 | 0.00 | 553.17 | 0.120 |
| 5 | rs112951499 | 50,697 | LOC105374602 | ACCCT | A | 415,296 | 0.937 | 0.034 | 0.004 | 8.1E-15 | 61.46 | 0.015 | 0.03 | 0.00 | 53.79 | 0.013 |
| 5 | rs7705526 | 1,285,974 | | A | C | 451,006 | 0.673 | 0.078 | 0.002 | 2.4E-282 | 1154.14 | 0.255 | 0.08 | 0.00 | 1154.14 | 0.255 |
| 5 | rs61748181 | 1,294,166 | | C | T | 463,851 | 0.971 | 0.059 | 0.006 | 2.8E-23 | 101.11 | 0.022 | 0.06 | 0.01 | 101.09 | 0.022 |
| 5 | 5:78951569_GT_G | 78,951,569 | TENT2 (PAPD4) | GT | G | 463,081 | 0.899 | 0.025 | 0.003 | 1.9E-13 | 56.83 | 0.012 | 0.02 | 0.00 | 44.77 | 0.010 |
| 5 | rs34255404 | 138,935,580 | UBE2D2 | A | G | 463,012 | 0.943 | 0.037 | 0.004 | 6.8E-18 | 72.79 | 0.016 | 0.04 | 0.00 | 70.69 | 0.015 |
| 6 | rs80324517 | 204,031 | LOC285766 | A | G | 464,716 | 0.952 | 0.04 | 0.005 | 1.8E-17 | 75.88 | 0.016 | 0.04 | 0.00 | 74.50 | 0.016 |
| 7 | rs117247304 | 69,010 | LOC102723672 | G | A | 372,378 | 0.976 | 0.064 | 0.007 | 4.2E-18 | 74.84 | 0.020 | 0.06 | 0.01 | 73.33 | 0.020 |
| 7 | rs13230646 | 23,930,316 | STK31 | T | C | 458,458 | 0.751 | 0.017 | 0.002 | 8.9E-14 | 54.82 | 0.012 | 0.02 | 0.00 | 40.20 | 0.009 |
| 7 | rs11769630 | 50,257,703 | IKZF1 | T | A | 455,587 | 0.928 | 0.026 | 0.004 | 4.3E-11 | 46.17 | 0.010 | 0.02 | 0.01 | 19.86 | 0.004 |
| 7 | rs2538745 | 76,310,784 | UPK3B | T | C | 459,153 | 0.397 | 0.013 | 0.002 | 3.1E-10 | 40.58 | 0.009 | 0.01 | 0.00 | 6.63 | 0.001 |

| Chr | Genetic Variant | BP_GRCh37 | GENE | A1 | A2 | N_imp | FREQ (A1) | Uncorrected for the winner's curse: Beta | SE | Pvalue | F | %var explained | Corrected for the winner's curse: Beta | SE | F | %var explained |
|---|---|---|---|---|---|---|---|---|---|---|---|---|---|---|---|---|
| 7 | rs2056726 | 99,780,283 | STAG3 | G | A | 464,552 | 0.786 | 0.023 | 0.003 | 7.9E-21 | 81.29 | 0.017 | 0.02 | 0.00 | 80.48 | 0.017 |
| 7 | rs609953 | 123,422,444 | RNU6-2 | A | T | 459,026 | 0.618 | 0.013 | 0.002 | 7.0E-11 | 32.06 | 0.007 | 0.00 | 0.01 | 0.01 | 0.000 |
| 7 | rs7790856 | 124,459,852 | POT1 | C | T | 464,345 | 0.711 | 0.044 | 0.002 | 1.8E-87 | 367.26 | 0.079 | 0.04 | 0.00 | 367.26 | 0.079 |
| 7 | rs4731541 | 128,678,236 | TNPO3 | C | G | 464,268 | 0.375 | 0.021 | 0.002 | 1.4E-23 | 105.88 | 0.023 | 0.02 | 0.00 | 105.86 | 0.023 |
| 7 | rs1985369 | 159,119,220 | | A | G | 445,833 | 0.132 | 0.031 | 0.003 | 3.6E-25 | 102.55 | 0.023 | 0.03 | 0.00 | 102.52 | 0.023 |
| 8 | rs2306646 | 21,846,586 | XPO7 | G | C | 461,926 | 0.441 | 0.021 | 0.002 | 3.3E-25 | 105.88 | 0.023 | 0.02 | 0.00 | 105.86 | 0.023 |
| 8 | rs762679 | 48,885,436 | MCM4 | A | T | 464,716 | 0.143 | 0.031 | 0.003 | 1.4E-27 | 102.55 | 0.022 | 0.03 | 0.00 | 102.52 | 0.022 |
| 8 | rs564224004 | 56,667,353 | TGS1 | C | TATTAT | 457,659 | 0.115 | 0.031 | 0.003 | 2.1E-22 | 102.55 | 0.022 | 0.03 | 0.00 | 102.52 | 0.022 |
| 8 | rs7012816 | 70,964,743 | PRDM14 | A | G | 464,716 | 0.870 | 0.018 | 0.003 | 3.0E-09 | 41.14 | 0.009 | 0.01 | 0.01 | 7.83 | 0.002 |
| 8 | rs10112752 | 73,958,718 | TERF1 | G | A | 462,003 | 0.570 | 0.029 | 0.002 | 9.5E-46 | 201.92 | 0.044 | 0.03 | 0.00 | 201.92 | 0.044 |
| 8 | rs540491189 | 74,150,379 | | T | G | 385,475 | 0.998 | 0.191 | 0.024 | 3.8E-16 | 64.81 | 0.017 | 0.19 | 0.02 | 59.64 | 0.015 |
| 8 | rs1023767 | 95,530,969 | VIRMA | G | A | 464,716 | 0.762 | 0.018 | 0.002 | 5.0E-15 | 61.46 | 0.013 | 0.02 | 0.00 | 53.20 | 0.011 |
| 9 | rs4742448 | 826,585 | DMRT1 | G | C | 443,556 | 0.531 | 0.015 | 0.002 | 6.4E-14 | 54.02 | 0.012 | 0.01 | 0.00 | 38.31 | 0.009 |
| 9 | rs4743037 | 109,639,970 | ZNF462 | T | C | 462,541 | 0.769 | 0.015 | 0.002 | 5.1E-10 | 42.68 | 0.009 | 0.01 | 0.00 | 11.31 | 0.002 |
| 10 | rs762222726 | 5,816,070 | ASB13 | ACAT | C | 462,547 | 0.399 | 0.018 | 0.002 | 1.9E-19 | 77.79 | 0.017 | 0.02 | 0.00 | 76.32 | 0.016 |
| 10 | rs12572897 | 96,114,835 | NOC3L | G | A | 464,716 | 0.870 | 0.032 | 0.003 | 3.6E-27 | 109.27 | 0.024 | 0.03 | 0.00 | 109.26 | 0.024 |
| 10 | 10:101274251_CT_C | 101,274,251 | NKX2-3 | CT | C | 463,870 | 0.620 | 0.022 | 0.002 | 1.9E-27 | 116.20 | 0.025 | 0.02 | 0.00 | 116.20 | 0.025 |
| 10 | rs4919611 | 103,894,939 | PPRC1 | C | A | 464,716 | 0.113 | 0.026 | 0.003 | 5.1E-16 | 61.46 | 0.013 | 0.03 | 0.00 | 53.63 | 0.012 |
| 10 | rs9419958 | 105,675,946 | | T | C | 463,885 | 0.139 | 0.081 | 0.003 | 2.6E-167 | 700.11 | 0.151 | 0.08 | 0.00 | 700.11 | 0.151 |
| 11 | rs939916 | 202,253 | ODF3 | A | G | 458,773 | 0.330 | 0.024 | 0.002 | 6.6E-29 | 138.29 | 0.030 | 0.02 | 0.00 | 138.29 | 0.030 |
| 11 | rs10840270 | 9,629,553 | WEE1 | G | C | 460,657 | 0.344 | 0.014 | 0.002 | 1.3E-11 | 37.18 | 0.008 | 0.01 | 0.01 | 1.11 | 0.000 |
| 11 | rs611646 | 108,177,097 | | T | A | 464,305 | 0.591 | 0.037 | 0.002 | 3.5E-73 | 328.68 | 0.071 | 0.04 | 0.00 | 328.68 | 0.071 |
| 11 | rs6590343 | 128,500,215 | FLI1 | G | A | 463,852 | 0.484 | 0.012 | 0.002 | 1.5E-09 | 34.57 | 0.007 | 0.00 | 0.01 | 0.01 | 0.000 |
| 12 | rs10845387 | 11,757,743 | LINC01252 | G | A | 464,716 | 0.647 | 0.014 | 0.002 | 1.5E-11 | 47.06 | 0.010 | 0.01 | 0.00 | 21.96 | 0.005 |
| 12 | rs12369950 | 24,762,109 | LINC00477 | T | C | 462,282 | 0.859 | 0.018 | 0.003 | 8.0E-10 | 34.57 | 0.007 | 0.00 | 0.02 | 0.00 | 0.000 |
| 12 | rs79977579 | 54,694,560 | SMUG1 | A | C | 457,062 | 0.904 | 0.028 | 0.004 | 2.3E-16 | 61.46 | 0.013 | 0.03 | 0.00 | 53.68 | 0.012 |
| 12 | rs1907702 | 88,955,469 | KITLG | A | G | 462,976 | 0.233 | 0.015 | 0.003 | 5.9E-10 | 34.57 | 0.007 | 0.00 | 0.01 | 0.00 | 0.000 |
| 12 | rs76666449 | 120,904,895 | SRSF9 | C | T | 461,128 | 0.899 | 0.03 | 0.003 | 8.2E-19 | 81.83 | 0.018 | 0.03 | 0.00 | 81.18 | 0.018 |
| 12 | rs4758644 | 122,943,915 | ZCCHC8 | A | C | 464,129 | 0.269 | 0.017 | 0.002 | 2.7E-13 | 54.82 | 0.012 | 0.02 | 0.00 | 40.20 | 0.009 |
| 12 | rs79228077 | 133,046,343 | MUC8 | G | T | 394,231 | 0.369 | 0.015 | 0.002 | 2.3E-11 | 54.02 | 0.014 | 0.01 | 0.00 | 38.31 | 0.010 |
| 13 | rs1332941 | 41,695,100 | KBTBD6 | G | A | 460,016 | 0.180 | 0.026 | 0.003 | 5.9E-21 | 85.85 | 0.019 | 0.03 | 0.00 | 85.45 | 0.019 |
| 13 | rs35017269 | 73,340,177 | DIS3 | A | G | 447,674 | 0.984 | 0.067 | 0.008 | 1.6E-16 | 67.36 | 0.015 | 0.07 | 0.01 | 63.46 | 0.014 |
| 14 | rs3093888 | 20,812,951 | TEP1 | G | A | 463,795 | 0.949 | 0.029 | 0.005 | 1.5E-10 | 39.88 | 0.009 | 0.02 | 0.01 | 5.21 | 0.001 |
| 14 | rs73581419 | 21,941,148 | RAB2B | T | C | 462,410 | 0.893 | 0.023 | 0.003 | 1.3E-12 | 56.45 | 0.012 | 0.02 | 0.00 | 43.93 | 0.009 |
| 14 | rs12884911 | 65,027,871 | PPP1R36 | C | T | 463,054 | 0.497 | 0.013 | 0.002 | 2.9E-11 | 40.58 | 0.009 | 0.01 | 0.00 | 6.63 | 0.001 |
| 14 | rs762810 | 65,544,367 | MAX | C | A | 463,076 | 0.648 | 0.02 | 0.002 | 4.3E-22 | 96.04 | 0.021 | 0.02 | 0.00 | 95.93 | 0.021 |
| 14 | rs137901416 | 73,418,095 | DCAF4 | A | G | 463,864 | 0.900 | 0.046 | 0.003 | 4.7E-43 | 192.39 | 0.041 | 0.05 | 0.00 | 192.39 | 0.041 |
| 14 | 14:91970514_GA_G | 91,970,514 | PPP4R3A | G | GA | 463,412 | 0.544 | 0.019 | 0.002 | 6.3E-22 | 86.67 | 0.019 | 0.02 | 0.00 | 86.22 | 0.019 |
| 14 | rs1957937 | 96,181,360 | TCL1A | T | A | 463,272 | 0.840 | 0.021 | 0.003 | 1.9E-14 | 67.76 | 0.015 | 0.02 | 0.00 | 63.49 | 0.014 |
| 15 | rs17677991 | 42,032,383 | MGA | G | C | 464,716 | 0.658 | 0.022 | 0.002 | 4.4E-26 | 116.20 | 0.025 | 0.02 | 0.00 | 116.20 | 0.025 |
| 15 | rs181647350 | 50,379,219 | ATP8B4 | T | C | 455,018 | 0.761 | 0.034 | 0.002 | 7.7E-46 | 219.30 | 0.048 | 0.03 | 0.00 | 219.30 | 0.048 |
| 15 | rs1980240 | 56,774,018 | TEX9 | C | A | 462,804 | 0.595 | 0.013 | 0.002 | 2.2E-10 | 40.58 | 0.009 | 0.01 | 0.00 | 6.63 | 0.001 |
| 16 | rs80116508 | 3,650,970 | SLX4 | G | A | 458,739 | 0.938 | 0.035 | 0.004 | 2.0E-17 | 73.53 | 0.016 | 0.03 | 0.00 | 71.60 | 0.016 |
| 16 | rs11646283 | 9,073,060 | USP7 | C | T | 461,847 | 0.589 | 0.015 | 0.002 | 5.3E-14 | 54.02 | 0.012 | 0.01 | 0.00 | 38.31 | 0.008 |
| 16 | rs182059586 | 14,652,220 | PARN | T | C | 408,252 | 0.975 | 0.057 | 0.007 | 4.9E-17 | 73.85 | 0.018 | 0.06 | 0.01 | 72.11 | 0.018 |
| 16 | rs450962 | 28,413,517 | EIF3CL | G | A | 384,323 | 0.716 | 0.014 | 0.003 | 5.9E-09 | 30.12 | 0.008 | 0.00 | 0.01 | 0.00 | 0.000 |
| 16 | rs8053839 | 48,390,512 | LONP2 | G | T | 457,145 | 0.458 | 0.014 | 0.002 | 1.0E-11 | 47.06 | 0.010 | 0.01 | 0.00 | 21.96 | 0.005 |
| 16 | rs12447324 | 50,089,038 | PAPD5 | A | C | 449,188 | 0.803 | 0.017 | 0.003 | 4.4E-11 | 44.41 | 0.010 | 0.02 | 0.00 | 15.45 | 0.003 |
| 16 | rs76219171 | 50,188,929 | | A | G | 450,771 | 0.942 | 0.036 | 0.004 | 7.8E-17 | 77.79 | 0.017 | 0.04 | 0.00 | 76.71 | 0.017 |
| 16 | rs139438549 | 67,692,863 | | C | T | 452,323 | 0.999 | 0.256 | 0.030 | 2.7E-17 | 71.11 | 0.016 | 0.26 | 0.03 | 68.70 | 0.015 |
| 16 | rs142507451 | 67,694,044 | | C | T | 461,205 | 0.997 | 0.142 | 0.019 | 1.6E-14 | 58.14 | 0.013 | 0.14 | 0.02 | 47.79 | 0.010 |
| 16 | rs528301822 | 69,403,012 | TERF2 | T | A | 464,229 | 0.711 | 0.024 | 0.002 | 1.7E-27 | 138.29 | 0.030 | 0.02 | 0.00 | 138.29 | 0.030 |
| 16 | rs62053340 | 69,987,764 | EXOSC6 | C | T | 457,598 | 0.617 | 0.021 | 0.002 | 1.2E-24 | 105.88 | 0.023 | 0.02 | 0.00 | 105.86 | 0.023 |

| | | | | | | | | Uncorrected for the winner's curse | | | | | Corrected for the winner's curse | | | |
|---|---|---|---|---|---|---|---|---|---|---|---|---|---|---|---|---|
| **Chr** | **Genetic Variant** | **BP_GRCh37** | **GENE** | **A1** | **A2** | **N_imp** | **FREQ (A1)** | **Beta** | **SE** | **Pvalue** | **F** | **%var explained** | **Beta** | **SE** | **F** | **%var explained** |
| 16 | rs34003787 | 73,071,381 | ZFHX3 | C | T | 449,493 | 0.912 | 0.024 | 0.004 | 1.9E-11 | 45.16 | 0.010 | 0.02 | 0.01 | 17.33 | 0.004 |
| 16 | rs11866592 | 74,654,396 | | A | G | 464,397 | 0.858 | 0.035 | 0.003 | 1.3E-33 | 155.56 | 0.033 | 0.03 | 0.00 | 155.56 | 0.033 |
| 16 | rs2303262 | 82,203,758 | MPHOSPH6 | C | T | 464,716 | 0.223 | 0.047 | 0.003 | 2.9E-85 | 339.43 | 0.073 | 0.05 | 0.00 | 339.43 | 0.073 |
| 16 | rs62046862 | 88,073,029 | BANP | A | C | 441,263 | 0.479 | 0.024 | 0.002 | 4.9E-31 | 138.29 | 0.031 | 0.02 | 0.00 | 138.29 | 0.031 |
| 16 | rs9923119 | 90,153,815 | PRDM7 | C | T | 446,260 | 0.750 | 0.018 | 0.002 | 1.8E-13 | 61.46 | 0.014 | 0.02 | 0.00 | 53.20 | 0.012 |
| 17 | rs7218033 | 1,694,247 | RPA1 | C | T | 460,449 | 0.747 | 0.023 | 0.002 | 1.0E-22 | 100.35 | 0.022 | 0.02 | 0.00 | 100.31 | 0.022 |
| 17 | rs4724 | 7,760,397 | CTC1 | G | A | 464,716 | 0.883 | 0.055 | 0.003 | 9.8E-69 | 322.79 | 0.069 | 0.06 | 0.00 | 322.79 | 0.069 |
| 17 | rs75664430 | 8,064,779 | | C | G | 462,922 | 0.752 | 0.024 | 0.002 | 3.6E-24 | 109.27 | 0.024 | 0.02 | 0.00 | 109.26 | 0.024 |
| 17 | rs111527438 | 29,252,703 | ADAP2 | C | T | 462,353 | 0.649 | 0.013 | 0.002 | 3.1E-09 | 32.06 | 0.007 | 0.00 | 0.01 | 0.01 | 0.000 |
| 17 | rs12941945 | 41,448,228 | | A | G | 458,692 | 0.832 | 0.026 | 0.003 | 3.0E-22 | 103.87 | 0.023 | 0.03 | 0.00 | 103.85 | 0.023 |
| 17 | rs34405642 | 65,741,012 | NOL11 | GA | G | 413,522 | 0.415 | 0.013 | 0.002 | 2.6E-09 | 40.58 | 0.010 | 0.01 | 0.00 | 6.63 | 0.002 |
| 17 | rs2069536 | 74,001,106 | TEN1 | A | G | 463,298 | 0.250 | 0.015 | 0.002 | 1.8E-10 | 42.68 | 0.009 | 0.01 | 0.00 | 11.31 | 0.002 |
| 17 | rs144204502 | 76,183,233 | | C | T | 447,012 | 0.987 | 0.101 | 0.009 | 3.4E-28 | 127.96 | 0.029 | 0.10 | 0.01 | 127.96 | 0.029 |
| 18 | rs3891167 | 658,423 | | A | G | 427,300 | 0.747 | 0.043 | 0.002 | 1.2E-70 | 350.76 | 0.082 | 0.04 | 0.00 | 350.76 | 0.082 |
| 18 | rs78694226 | 710,980 | | G | A | 452,168 | 0.992 | 0.066 | 0.011 | 6.8E-09 | 33.05 | 0.007 | 0.00 | 0.07 | 0.00 | 0.000 |
| 18 | rs8088824 | 42,151,261 | | T | C | 462,570 | 0.237 | 0.026 | 0.002 | 8.1E-28 | 128.24 | 0.028 | 0.03 | 0.00 | 128.24 | 0.028 |
| 18 | rs2276182 | 51,798,047 | POLI | G | C | 464,368 | 0.597 | 0.023 | 0.002 | 2.8E-30 | 127.01 | 0.027 | 0.02 | 0.00 | 127.01 | 0.027 |
| 18 | rs6565924 | 74,691,225 | ZNF236 | G | A | 461,318 | 0.639 | 0.013 | 0.002 | 2.9E-10 | 40.58 | 0.009 | 0.01 | 0.00 | 6.63 | 0.001 |
| 18 | rs1879100 | 77,985,740 | PARD6G | C | T | 452,390 | 0.138 | 0.019 | 0.003 | 8.1E-11 | 38.52 | 0.009 | 0.01 | 0.01 | 2.85 | 0.001 |
| 19 | rs80337039 | 4,105,089 | MAP2K2 | T | G | 388,380 | 0.994 | 0.081 | 0.014 | 2.3E-09 | 35.89 | 0.009 | 0.02 | 0.06 | 0.16 | 0.000 |
| 19 | rs35601737 | 13,220,703 | TRMT1 | C | G | 464,716 | 0.704 | 0.014 | 0.002 | 1.9E-10 | 47.06 | 0.010 | 0.01 | 0.00 | 21.96 | 0.005 |
| 19 | rs8105767 | 22,215,441 | ZNF208 | G | A | 461,895 | 0.705 | 0.033 | 0.002 | 2.5E-50 | 261.46 | 0.057 | 0.03 | 0.00 | 261.46 | 0.057 |
| 19 | rs4530278 | 33,752,994 | CEBPA | T | G | 457,460 | 0.402 | 0.014 | 0.002 | 1.5E-11 | 47.06 | 0.010 | 0.01 | 0.00 | 21.96 | 0.005 |
| 19 | rs11084431 | 56,708,667 | ZSCAN5B | G | A | 451,567 | 0.396 | 0.012 | 0.002 | 4.0E-09 | 34.57 | 0.008 | 0.00 | 0.01 | 0.01 | 0.000 |
| 19 | rs8102497 | 57,370,055 | PEG3 | G | A | 461,793 | 0.568 | 0.015 | 0.002 | 1.4E-13 | 54.02 | 0.012 | 0.01 | 0.00 | 38.31 | 0.008 |
| 20 | rs1291143 | 35,525,640 | SAMHD1 | C | A | 463,481 | 0.151 | 0.049 | 0.003 | 1.8E-69 | 304.90 | 0.066 | 0.05 | 0.00 | 304.90 | 0.066 |
| 20 | rs544699357 | 41,997,057 | SRSF6 | T | C | 373,245 | 0.998 | 0.169 | 0.025 | 1.1E-11 | 45.70 | 0.012 | 0.16 | 0.04 | 18.67 | 0.005 |
| 20 | rs577449057 | 62,236,709 | RTEL1 | A | G | 268,900 | 0.995 | 0.154 | 0.018 | 1.4E-17 | 72.29 | 0.027 | 0.15 | 0.02 | 70.22 | 0.026 |
| 20 | rs187013287 | 62,298,374 | | A | T | 370,530 | 0.997 | 0.283 | 0.022 | 7.9E-38 | 166.39 | 0.045 | 0.28 | 0.02 | 166.39 | 0.045 |
| 20 | rs35640778 | 62,321,128 | | G | A | 464,716 | 0.979 | 0.209 | 0.007 | 9.6E-195 | 856.12 | 0.184 | 0.21 | 0.01 | 856.12 | 0.184 |
| 20 | 20:62321690_GAGA_( | 62,321,690 | | G | GAGA | 349,451 | 0.996 | 0.118 | 0.017 | 1.3E-11 | 46.27 | 0.013 | 0.11 | 0.02 | 20.13 | 0.006 |
| 20 | rs115610405 | 62,325,833 | | C | A | 464,716 | 0.981 | 0.11 | 0.007 | 1.8E-50 | 221.08 | 0.048 | 0.11 | 0.01 | 221.08 | 0.048 |
| 20 | rs187577818 | 62,358,869 | | C | A | 373,318 | 0.997 | 0.298 | 0.021 | 6.6E-44 | 193.39 | 0.052 | 0.30 | 0.02 | 193.39 | 0.052 |
| 20 | rs111527478 | 62,678,100 | | A | G | 457,574 | 0.900 | 0.022 | 0.003 | 1.2E-10 | 44.01 | 0.010 | 0.02 | 0.01 | 14.48 | 0.003 |
| 22 | rs28502153 | 17,469,049 | GAB4 | C | A | 464,716 | 0.622 | 0.022 | 0.002 | 1.2E-25 | 116.20 | 0.025 | 0.02 | 0.00 | 116.20 | 0.025 |
| 22 | rs5845706 | 45,779,013 | SMC1B | CA | C | 459,636 | 0.644 | 0.015 | 0.002 | 2.2E-12 | 54.02 | 0.012 | 0.01 | 0.00 | 38.31 | 0.008 |
| 22 | rs131796 | 50,971,639 | TYMP | G | GA | 460,083 | 0.236 | 0.024 | 0.002 | 6.7E-25 | 109.27 | 0.024 | 0.02 | 0.00 | 109.26 | 0.024 |
| | | | | | | | | | | | | 3.619 | | | | 3.267 |
