## Supplement 2 for "Mid-life leukocyte telomere length and dementia risk: a prospective cohort study of 435,046 UK Biobank participants"

**eTable 1 |** UKB field IDs and ICD-10 codes for identification of AD/ADRD

**eTable 2 |** A selection of UK Biobank cognitive tests

**eTable 3 |** Selected imaging derived phenotypes

**eTable 4 |** Baseline characteristics of participants in the baseline or imaging cohort

**eTable 5 |** Participant characteristics at baseline of incident AD/ADRD cases versus normal controls

**eFigure 1 |** Relationship between baseline leukocyte telomere length and incidence of AD/ADRD

**eFigure 2 |** Associations between genetically determined telomere length and AD/ADRD using the Inverse Variance Weighted (IVW) method

**eFigure 3 |** Associations between genetically determined telomere length and cognitive function using the Inverse Variance Weighted (IVW) method

**eFigure 4 |** Associations between genetically determined telomere length and volumetric IDPs of AD signatures and white matter hyperintensities (WMH) using the Inverse Variance Weighted (IVW) method

**eFigure 5 |** Associations between genetically determined telomere length and weighted-mean fractional anisotropy IDPs using the Inverse Variance Weighted (IVW) method

**eFigure 6 |** Associations between genetically determined telomere length and weighted-mean mean diffusivity IDPs using the Inverse Variance Weighted (IVW) method

**eFigure 7 |** Associations between genetically determined telomere length and AD/ADRD, comparing the primary (IVW: inverse variance weighted) to secondary MR methods (Mendelian randomization robust adjusted profile score (MR-RAPS); weighted median based method; MR-Egger method)

**eFigure 8 |** Associations between genetically determined telomere length and cognitive function, comparing the primary (IVW: inverse variance weighted) to secondary MR methods (Mendelian randomization robust adjusted profile score (MR-RAPS); weighted median based method; MR-Egger method)

**eFigure 9 |** Associations between genetically determined telomere length and volumetric IDPs of AD signatures and white matter hyperintensities (WMH), comparing the primary (IVW: inverse variance weighted) to secondary MR methods (Mendelian randomization robust adjusted profile score (MR-RAPS); weighted median based method; MR-Egger method)

**eFigure 10 |** Associations between genetically determined telomere length and weighted-mean fractional anisotropy IDPs, comparing the primary (IVW: inverse variance weighted) to secondary MR methods (Mendelian randomization robust adjusted profile score (MR-RAPS); weighted median based method; MR-Egger method)

**eFigure 11 |** Associations between genetically determined telomere length and weighted-mean mean diffusivity IDPs, comparing the primary (IVW: inverse variance weighted) to secondary MR methods (Mendelian randomization robust adjusted profile score (MR-RAPS); weighted median based method; MR-Egger method)

### eTable 1. UKB field IDs and ICD-10 codes for identification of AD/ADRD

| AD/ADRD | UKB field ID | ICD-10 | Data release period currently available in England* |
|---|---|---|---|
| Alzheimer's disease (AD) | 131036 | G30 | Death data: April 2006-February 2021 |
| Dementia in AD | 130836 | F00 | |
| Vascular dementia | 130838 | F01 | Hospital inpatient data: 1997-March 2021 for half of the UKB cohort |
| Unspecified dementia | 130842 | F03 | |
| Other degenerative diseases of nervous system, not elsewhere classified, including frontotemporal dementia and Lewy body's dementia | 131038 | G31 | Primary care data: 1938-2017<br>Self-reported data: initial assessment visit (2006-2010), first repeat assessment visit (2012-2013), imaging visit (2014+), first repeat imaging visit (2019+) |

*Data release period varied but similar for different countries in the United Kingdom; participants from England account for over 95% of the cohort.

### eTable 2. A selection of UK Biobank cognitive tests

| Cognitive domain | Cognitive test | Field ID | Baseline visit | First imaging visit | Web link |
|---|---|---|---|---|---|
| **Processing speed** | Reaction time | 20023 | G √ | | https://biobank.ndph.ox.ac.uk/showcase/field.cgi?id=20023 |
| **Working memory** | Numeric memory | 4282 | G √ | | https://biobank.ndph.ox.ac.uk/showcase/field.cgi?id=4282 |
| **Verbal and numerical reasoning** | Fluid intelligence | 20016 | G √ | | https://biobank.ndph.ox.ac.uk/showcase/field.cgi?id=20016 |
| **Prospective memory** | Prospective memory | 20018 | G | | https://biobank.ctsu.ox.ac.uk/crystal/field.cgi?id=20018 |
| **Visual declarative memory** | Pairs matching | 399 | G | | https://biobank.ndph.ox.ac.uk/showcase/field.cgi?id=399 |
| **Processing speed** | Symbol digit substitution | 23324 | | √ | https://biobank.ndph.ox.ac.uk/showcase/field.cgi?id=23324 |
| **Executive function** | Trail making part B | 6350 | | √ | https://biobank.ndph.ox.ac.uk/showcase/field.cgi?id=6350 |
| **Non-verbal reasoning** | Matrix pattern completion | 6373 | | √ | https://biobank.ndph.ox.ac.uk/showcase/field.cgi?id=6373 |

G: general cognitive ability element

### eTable 3. Selected imaging derived phenotypes

| Imaging derived phenotype (IDP) | UKB Field ID |
|---|---|
| Volume of hippocampus (from T1 brain image) | 25019/25020 |
| Volume of precuneus generated by parcellation of the white surface using Desikan-Killiany parcellation | 26812/26913 |
| Volume of cuneus generated by parcellation of the white surface using Desikan-Killiany parcellation | 26792/26893 |
| Volume of entorhinal generated by parcellation of the white surface using Desikan-Killiany parcellation | 26793/26894 |
| Volume of inferiorparietal generated by parcellation of the white surface using Desikan-Killiany parcellation | 26795/26896 |
| Volume of parahippocampal generated by parcellation of the white surface using Desikan-Killiany parcellation | 26803/26904 |
| Total volume of white matter hyperintensities (from T1 and T2_FLAIR images) | 25781 |
| Mean FA (fractional anisotropy) in anterior corona radiata on FA skeleton | 25079/25078 |
| Mean FA (fractional anisotropy) in anterior limb of internal capsule on FA skeleton | 25073/25072 |
| Mean FA (fractional anisotropy) in body of corpus callosum on FA skeleton | 25059 |
| Mean FA (fractional anisotropy) in cerebral peduncle on FA skeleton | 25071/25070 |
| Mean FA (fractional anisotropy) in cingulum cingulate gyrus on FA skeleton | 25091/25090 |
| Mean FA (fractional anisotropy) in cingulum hippocampus on FA skeleton | 25093/25092 |
| Mean FA (fractional anisotropy) in corticospinal tract on FA skeleton | 25063/25062 |
| Mean FA (fractional anisotropy) in external capsule on FA skeleton | 25089/25088 |
| Mean FA (fractional anisotropy) in fornix cres+stria terminalis on FA skeleton | 25095/25094 |
| Mean FA (fractional anisotropy) in fornix on FA skeleton | 25061 |
| Mean FA (fractional anisotropy) in genu of corpus callosum on FA skeleton | 25058 |
| Mean FA (fractional anisotropy) in inferior cerebellar peduncle on FA skeleton | 25067/25066 |
| Mean FA (fractional anisotropy) in medial lemniscus on FA skeleton | 25065/25064 |
| Mean FA (fractional anisotropy) in middle cerebellar peduncle on FA skeleton | 25056 |
| Mean FA (fractional anisotropy) in pontine crossing tract on FA skeleton | 25057 |
| Mean FA (fractional anisotropy) in posterior corona radiata on FA skeleton | 25083/25082 |
| Mean FA (fractional anisotropy) in posterior limb of internal capsule on FA skeleton | 25075/25074 |
| Mean FA (fractional anisotropy) in posterior thalamic radiation on FA skeleton | 25085/25084 |
| Mean FA (fractional anisotropy) in retrolenticular part of internal capsule on FA skeleton | 25077/25076 |
| Mean FA (fractional anisotropy) in sagittal stratum on FA skeleton | 25087/25086 |
| Mean FA (fractional anisotropy) in splenium of corpus callosum on FA skeleton | 25060 |
| Mean FA (fractional anisotropy) in superior cerebellar peduncle on FA skeleton | 25069/25068 |
| Mean FA (fractional anisotropy) in superior corona radiata on FA skeleton | 25081/25080 |
| Mean FA (fractional anisotropy) in superior fronto-occipital fasciculus on FA skeleton | 25099/25098 |
| Mean FA (fractional anisotropy) in superior longitudinal fasciculus on FA skeleton | 25097/25096 |
| Mean FA (fractional anisotropy) in tapetum on FA skeleton | 25103/25102 |
| Mean FA (fractional anisotropy) in uncinate fasciculus on FA skeleton | 25101/25100 |
| Mean MD (mean diffusivity) in anterior corona radiata on FA (fractional anisotropy) skeleton | 25127/25126 |
| Mean MD (mean diffusivity) in anterior limb of internal capsule on FA (fractional anisotropy) skeleton | 25121/25120 |
| Mean MD (mean diffusivity) in body of corpus callosum on FA (fractional anisotropy) skeleton | 25107 |
| Mean MD (mean diffusivity) in cerebral peduncle on FA (fractional anisotropy) skeleton | 25119/25118 |
| Mean MD (mean diffusivity) in cingulum cingulate gyrus on FA (fractional anisotropy) skeleton | 25139/25138 |
| Mean MD (mean diffusivity) in cingulum hippocampus on FA (fractional anisotropy) skeleton | 25141/25140 |

| | |
|---|---|
| Mean MD (mean diffusivity) in corticospinal tract on FA (fractional anisotropy) skeleton | 25111/25110 |
| Mean MD (mean diffusivity) in external capsule on FA (fractional anisotropy) skeleton | 25137/25136 |
| Mean MD (mean diffusivity) in fornix cres+stria terminalis on FA (fractional anisotropy) skeleton | 25143/25142 |
| Mean MD (mean diffusivity) in fornix on FA (fractional anisotropy) skeleton | 25109 |
| Mean MD (mean diffusivity) in genu of corpus callosum on FA (fractional anisotropy) skeleton | 25106 |
| Mean MD (mean diffusivity) in inferior cerebellar peduncle on FA (fractional anisotropy) skeleton | 25115/25114 |
| Mean MD (mean diffusivity) in medial lemniscus on FA (fractional anisotropy) skeleton | 25113/25112 |
| Mean MD (mean diffusivity) in middle cerebellar peduncle on FA (fractional anisotropy) skeleton | 25104 |
| Mean MD (mean diffusivity) in pontine crossing tract on FA (fractional anisotropy) skeleton | 25105 |
| Mean MD (mean diffusivity) in posterior corona radiata on FA (fractional anisotropy) skeleton | 25131/25130 |
| Mean MD (mean diffusivity) in posterior limb of internal capsule on FA (fractional anisotropy) skeleton | 25123/25122 |
| Mean MD (mean diffusivity) in posterior thalamic radiation on FA (fractional anisotropy) skeleton | 25133/25132 |
| Mean MD (mean diffusivity) in retrolenticular part of internal capsule on FA (fractional anisotropy) skeleton | 25125/25124 |
| Mean MD (mean diffusivity) in sagittal stratum on FA (fractional anisotropy) skeleton | 25135/25134 |
| Mean MD (mean diffusivity) in splenium of corpus callosum on FA (fractional anisotropy) skeleton | 25108 |
| Mean MD (mean diffusivity) in superior cerebellar peduncle on FA (fractional anisotropy) skeleton | 25117/25116 |
| Mean MD (mean diffusivity) in superior corona radiata on FA (fractional anisotropy) skeleton | 25129/25128 |
| Mean MD (mean diffusivity) in superior fronto-occipital fasciculus on FA (fractional anisotropy) skeleton | 25147/25146 |
| Mean MD (mean diffusivity) in superior longitudinal fasciculus on FA (fractional anisotropy) skeleton | 25145/25144 |
| Mean MD (mean diffusivity) in tapetum on FA (fractional anisotropy) skeleton | 25151/25150 |
| Mean MD (mean diffusivity) in uncinate fasciculus on FA (fractional anisotropy) skeleton | 25149/25148 |

**eTable 4 Baseline characteristics of participants in the baseline or imaging cohort**

| Characteristics | Baseline Cohort (N=435,046) | Imaging Cohort (N=43,390) |
|---|---|---|
| **Baseline age, years (mean ± SD)** | 56.8 ± 8 | 55.3 ± 7.5 |
| **Sex, female (%)** | 236,117 (54%) | 22,373 (52%) |
| **Death (%)** | 30,504 (7%) | 425 (1%) |
| **Age at death (mean ± SD)** | 71.1 ± 7.3 | 71.6 ± 6.4 |
| **Education (%)** | | |
| **None** | 75,541 (18%) | 2,984 (7%) |
| **CSEs or equivalent** | 16,231 (4%) | 1,122 (3%) |
| **O levels/GCSEs or equivalent** | 59,497 (13%) | 4,705 (11%) |
| **A/AS levels/NVQ/HND/HNC** | 79,609 (18%) | 7,778 (18%) |
| **Other professional qualifications** | 64,394 (15%) | 6,946 (16%) |
| **College or university degree** | 137,941 (32%) | 19,721 (46%) |
| **Townsend deprivation index** | -1.5 ± 3 | -1.9 ± 2.7 |
| **BMI, kg/m$^2$ (mean ± SD)** | 27.4 ± 4.8 | 26.6 ± 4.3 |
| **Smoking status (%)** | | |
| **Never** | 234,004 (54%) | 26,022 (60%) |
| **Previous** | 154,156 (36%) | 14,596 (34%) |
| **Current** | 45,343 (10%) | 2,687 (6%) |
| **Alcohol intake frequency (%)** | | |
| **Never** | 29,060 (7%) | 1,847 (4%) |
| **Special occasions only** | 46,553 (11%) | 3,323 (8%) |
| **1-3 times a month** | 48,359 (11%) | 4,619 (11%) |
| **1-2 times a week** | 114,513 (26%) | 11,195 (26%) |
| **3-4 times a week** | 104,311 (24%) | 12,345 (28%) |
| **Daily or almost daily** | 91,947 (21%) | 10,053 (23%) |
| **IPAQ activity group (%)** | | |
| **Low** | 57,696 (14%) | 5,742 (14%) |
| **Moderate** | 177,073 (44%) | 18,504 (45%) |
| **High** | 163,435 (41%) | 16,902 (41%) |
| ***APOE* genotype (%)** | | |
| **e3e3** | 254,331 (58.46%) | 25,619 (59.04%) |
| **e2e3** | 53,573 (12.31%) | 5,388 (12.42%) |
| **e2e2** | 2,766 (0.64%) | 250 (0.58%) |
| **e2e4** | 10,931 (2.51%) | 1,036 (2.39%) |
| **e3e4** | 103,077 (23.69%) | 10,122 (23.33%) |
| **e4e4** | 10,350 (2.38%) | 975 (2.25%) |
| **e1e2** | 3 (<0.01%) | 0 (0.00%) |
| **Telomere length (T/S ratio), adjusting for technical parameters** | 0.83 ± 0.13 | 0.84 ± 0.13 |

Abbreviations: SD, standard deviation; *APOE*, apolipoprotein E; CSE, certificate of secondary education; GCSE, general certificate of secondary education; NVQ, national vocational qualification; HND, higher national diploma; HNC, higher national certificate. BMI, body mass index; IPAQ, International Physical Activity Questionnaire.

**eTable 5. Participant characteristics at baseline of incident AD/ADRD cases versus normal controls**

| Characteristics | AD/ADRD (N=6,424) | Normal Controls (N=428,622) | P-Value |
|---|---|---|---|
| **Baseline age, years (mean ± SD)** | 64.0 ± 5.1 | 56.7 ± 8.0 | $< 2.2\times10^{-16}$ |
| **Sex, female (%)** | 2,973 (46%) | 233,144 (54%) | $< 2.2\times10^{-16}$ |
| **Death (%)** | 3,009 (47%) | 27,495 (6%) | $< 2.2\times10^{-16}$ |
| **Age at death (mean ± SD)** | 74.7 ± 5.3 | 69.4 ± 7.3 | |
| **Education (%)** | | | $< 2.2\times10^{-16}$ |
| **None** | 2,276 (36%) | 73,265 (17%) | |
| **CSEs or equivalent** | 119 (2%) | 16,112 (4%) | |
| **O levels/GCSEs or equivalent** | 817 (13%) | 56,680 (13%) | |
| **A/AS levels/NVQ/HND/HNC** | 998 (16%) | 78,611 (19%) | |
| **Other professional qualifications** | 845 (13%) | 63,549 (15%) | |
| **College or university degree** | 1,246 (20%) | 136,695 (32%) | |
| **Townsend deprivation index** | -0.9 ± 3.3 | -1.5 ± 3.0 | $7.3\times10^{-38}$ |
| **BMI, kg/m$^2$ (mean ± SD)** | 27.8 ± 5.0 | 27.4 ± 4.8 | $9.3\times10^{-12}$ |
| **Smoking status (%)** | | | $< 2.2\times10^{-16}$ |
| **Never** | 2,842 (45%) | 231,162 (54%) | |
| **Previous** | 2,777 (44%) | 151,379 (35%) | |
| **Current** | 762 (12%) | 44,581 (10%) | |
| **Alcohol intake frequency (%)** | | | $< 2.2\times10^{-16}$ |
| **Never** | 826 (13%) | 28,234 (7%) | |
| **Special occasions only** | 875 (14%) | 45,678 (11%) | |
| **1-3 times a month** | 636 (10%) | 47,723 (11%) | |
| **1-2 times a week** | 1,471 (23%) | 113,042 (26%) | |
| **3-4 times a week** | 1,182 (18%) | 103,129 (24%) | |
| **Daily or almost daily** | 1,420 (22%) | 90,527 (21%) | |
| **IPAQ activity group (%)** | | | $7.6\times10^{-7}$ |
| **Low** | 925 (16%) | 56,771 (14%) | |
| **Moderate** | 2,556 (45%) | 174,517 (44%) | |
| **High** | 2,144 (38%) | 161,291 (41%) | |
| ***APOE* genotype (%)** | | | $< 2.2\times10^{-16}$ |
| **e3e3** | 2,651 (41.27%) | 251,680 (59.72%) | |
| **e2e3** | 473 (7.36%) | 53,100 (12.39%) | |
| **e2e2** | 30 (0.47%) | 2,736 (0.64%) | |
| **e2e4** | 159 (2.48%) | 10,772 (2.51%) | |
| **e3e4** | 2,449 (38.12%) | 100,628 (23.48%) | |
| **e4e4** | 662 (10.31%) | 9,688 (2.26%) | |
| **e1e2** | 0 (0.00%) | 3 (<0.01%) | |
| **Telomere length (T/S ratio), adjusting for technical parameters** | 0.80 ± 0.12 | 0.83 ± 0.13 | $< 2.2\times10^{-16}$ |

Abbreviations: SD, standard deviation; *APOE*, apolipoprotein E; CSE, certificate of secondary education; GCSE, general certificate of secondary education; NVQ, national vocational qualification; HND, higher national diploma; HNC, higher national certificate. BMI, body mass index; IPAQ, International Physical Activity Questionnaire.

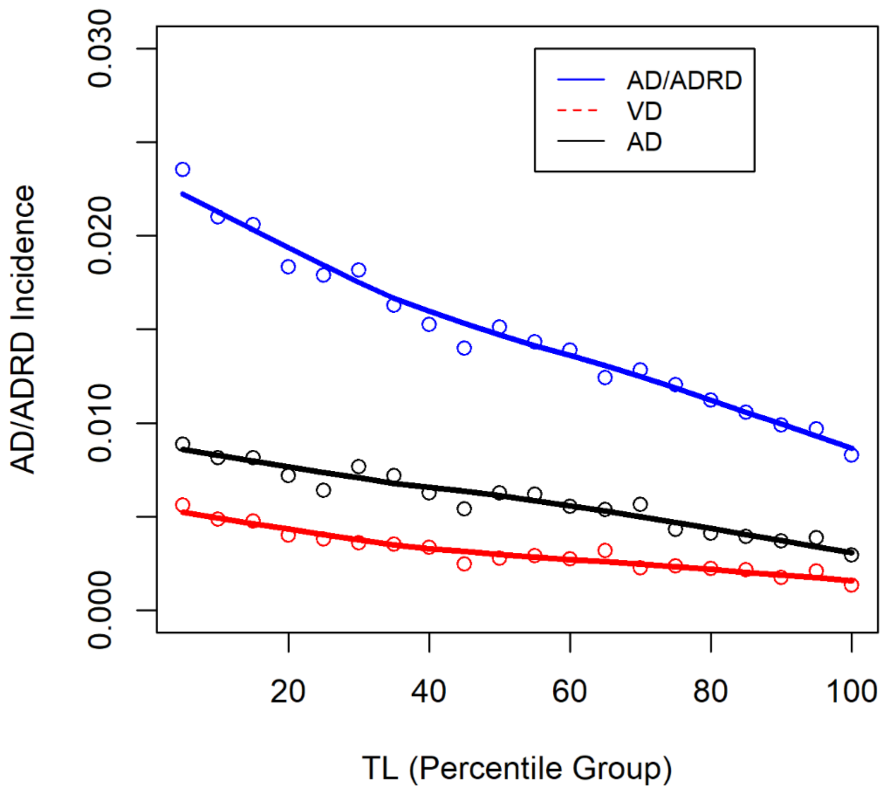


**eFigure 1. Relationship between baseline leukocyte telomere length and incidence of AD/ADRD.**

AD/ADRD incidence calculated for participants with TL between 0-2nd percentile, 3rd-4th percentile, …, or 98th-100th percentile and similarly for vascular dementia (VD) and Alzheimer's disease or dementia in Alzheimer's disease (AD).

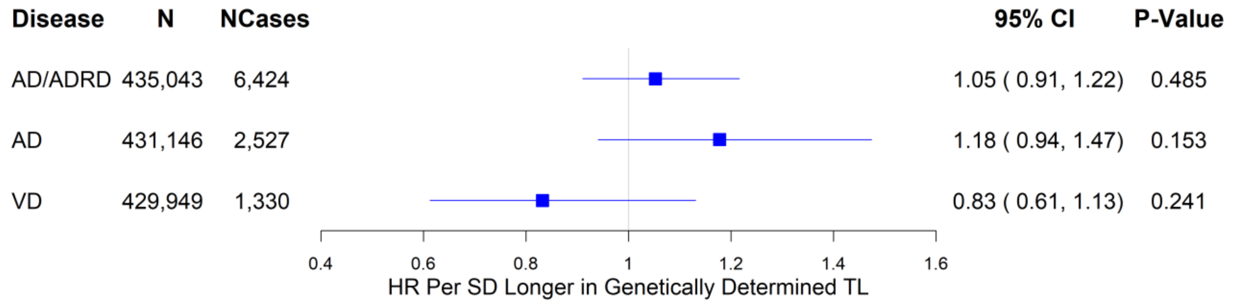


**eFigure 2.** Associations between genetically determined telomere length and AD/ADRD using the Inverse Variance Weighted (IVW) method

*Significant at the false discovery rate < 0.05 level; AD: Alzheimer's disease or dementia in Alzheimer's disease; VD: vascular dementia.

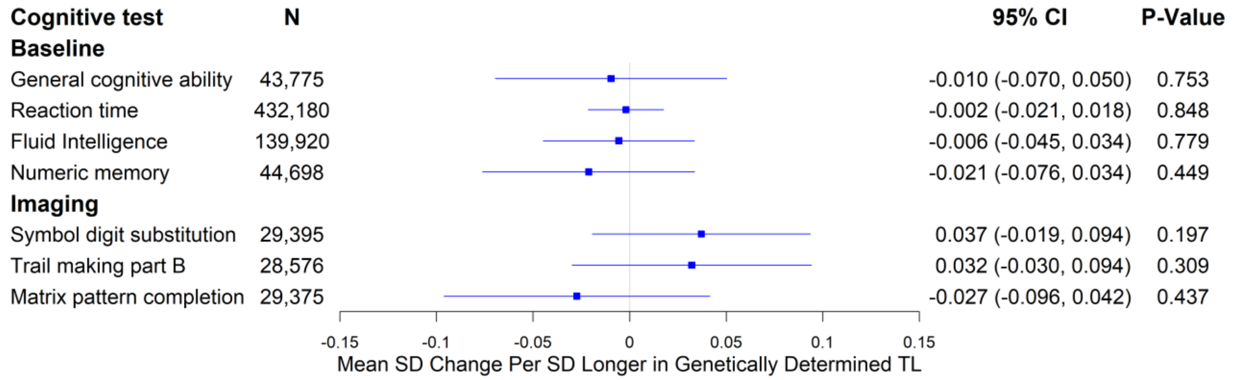


**eFigure 3.** Associations between genetically determined telomere length and cognitive function using the Inverse Variance Weighted (IVW) method

*Significant at the false discovery rate < 0.05 level.

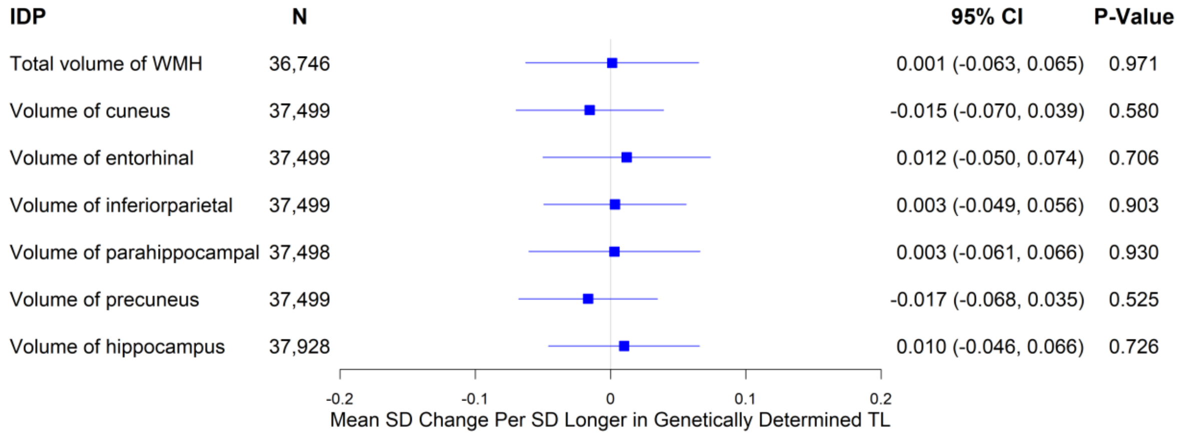


**eFigure 4.** Associations between genetically determined telomere length and volumetric IDPs of AD signatures and white matter hyperintensities (WMH) using the Inverse Variance Weighted (IVW) method

*Significant at the false discovery rate < 0.05 level.

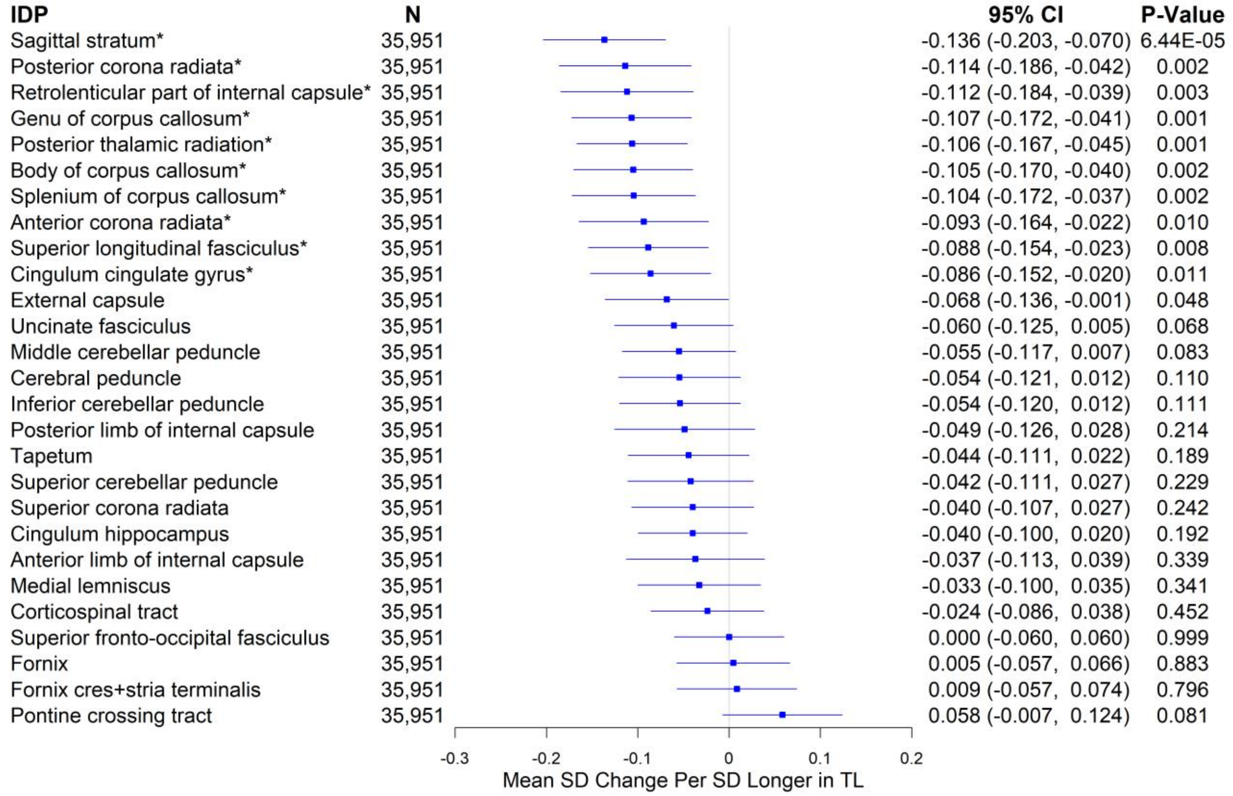


**eFigure 5.** Associations between genetically determined telomere length and weighted-mean fractional anisotropy IDPs using the Inverse Variance Weighted (IVW) method

*Significant at the false discovery rate < 0.05 level.

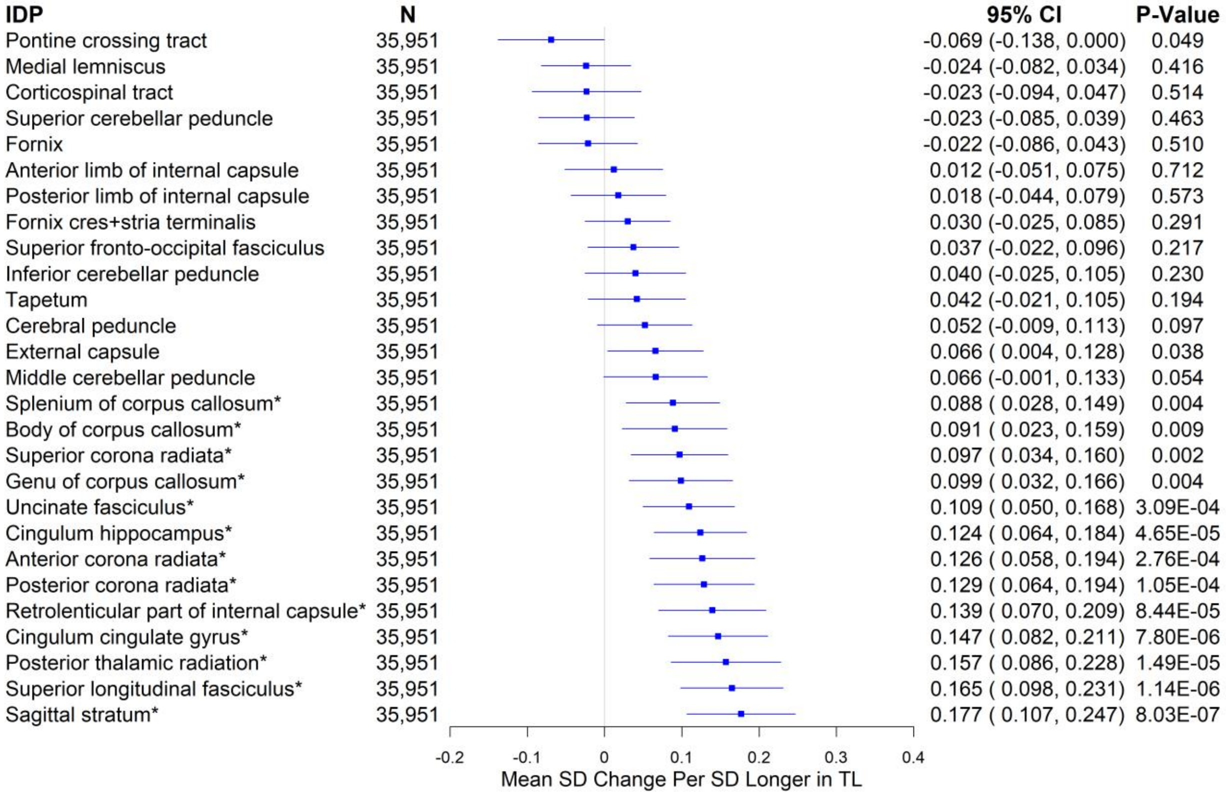


**eFigure 6.** Associations between genetically determined telomere length and weighted-mean mean diffusivity IDPs using the Inverse Variance Weighted (IVW) method

*Significant at the false discovery rate < 0.05 level.

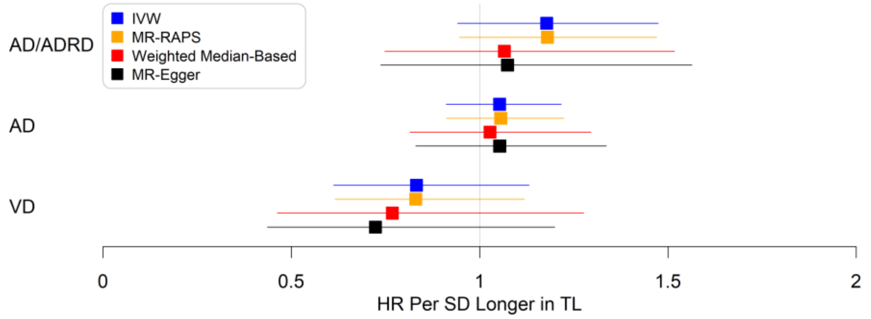


**eFigure 7.** Associations between genetically determined telomere length and AD/ADRD, comparing the primary (IVW: inverse variance weighted) to secondary MR methods (Mendelian randomization robust adjusted profile score (MR-RAPS); weighted median based method; MR-Egger method)

*Significant at the false discovery rate < 0.05 level using the IVW method; AD: Alzheimer's disease or dementia in Alzheimer's disease; VD: vascular dementia.

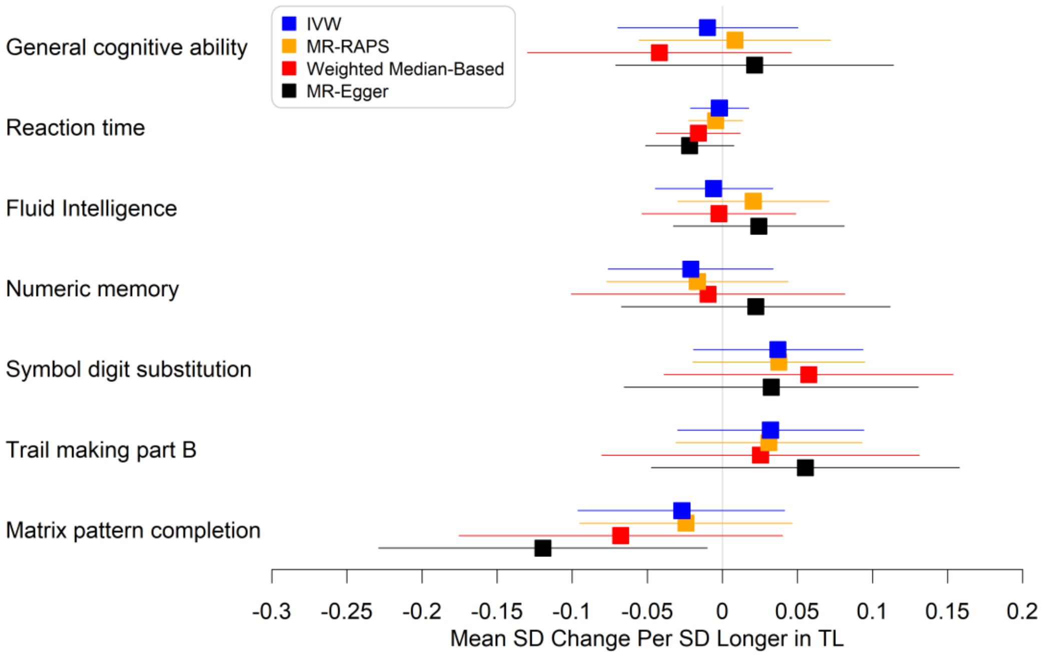


**eFigure 8.** Associations between genetically determined telomere length and cognitive function, comparing the primary (IVW: inverse variance weighted) to secondary MR methods (Mendelian randomization robust adjusted profile score (MR-RAPS); weighted median based method; MR-Egger method)

*Significant at the false discovery rate < 0.05 level using the IVW method.

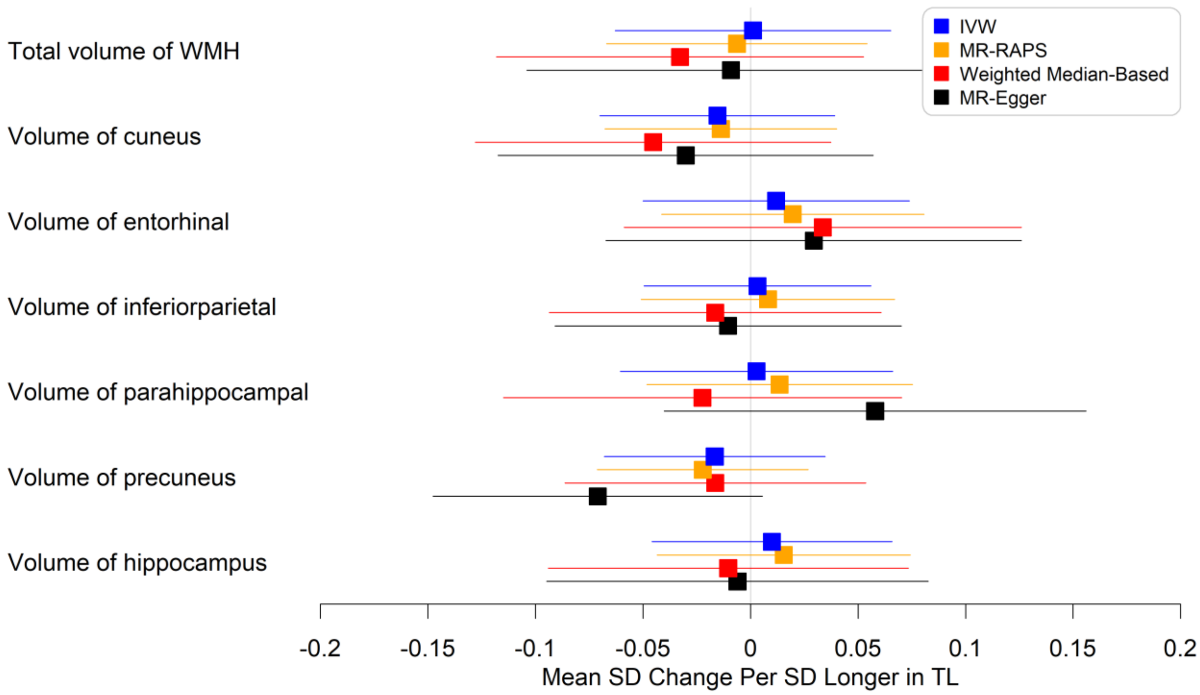


**eFigure 9.** Associations between genetically determined telomere length and volumetric IDPs of AD signatures and white matter hyperintensities (WMH), comparing the primary (IVW: inverse variance weighted) to secondary MR methods (Mendelian randomization robust adjusted profile score (MR-RAPS); weighted median based method; MR-Egger method)

*Significant at the false discovery rate < 0.05 level using the IVW method.

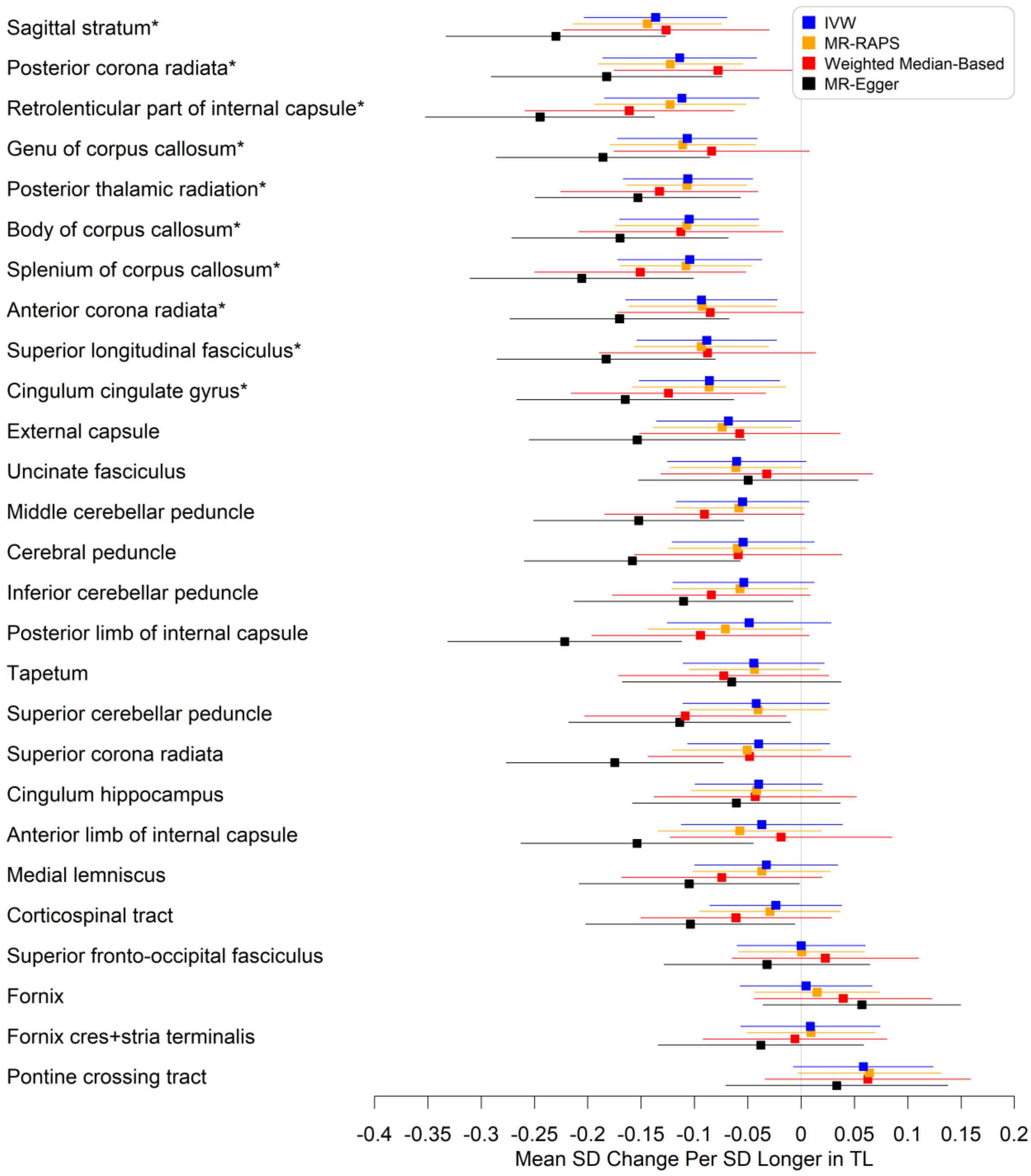


**eFigure 10.** Associations between genetically determined telomere length and weighted-mean fractional anisotropy IDPs, comparing the primary (IVW: inverse variance weighted) to secondary MR methods (Mendelian randomization robust adjusted profile score (MR-RAPS); weighted median based method; MR-Egger method)

*Significant at the false discovery rate < 0.05 level using the IVW method.

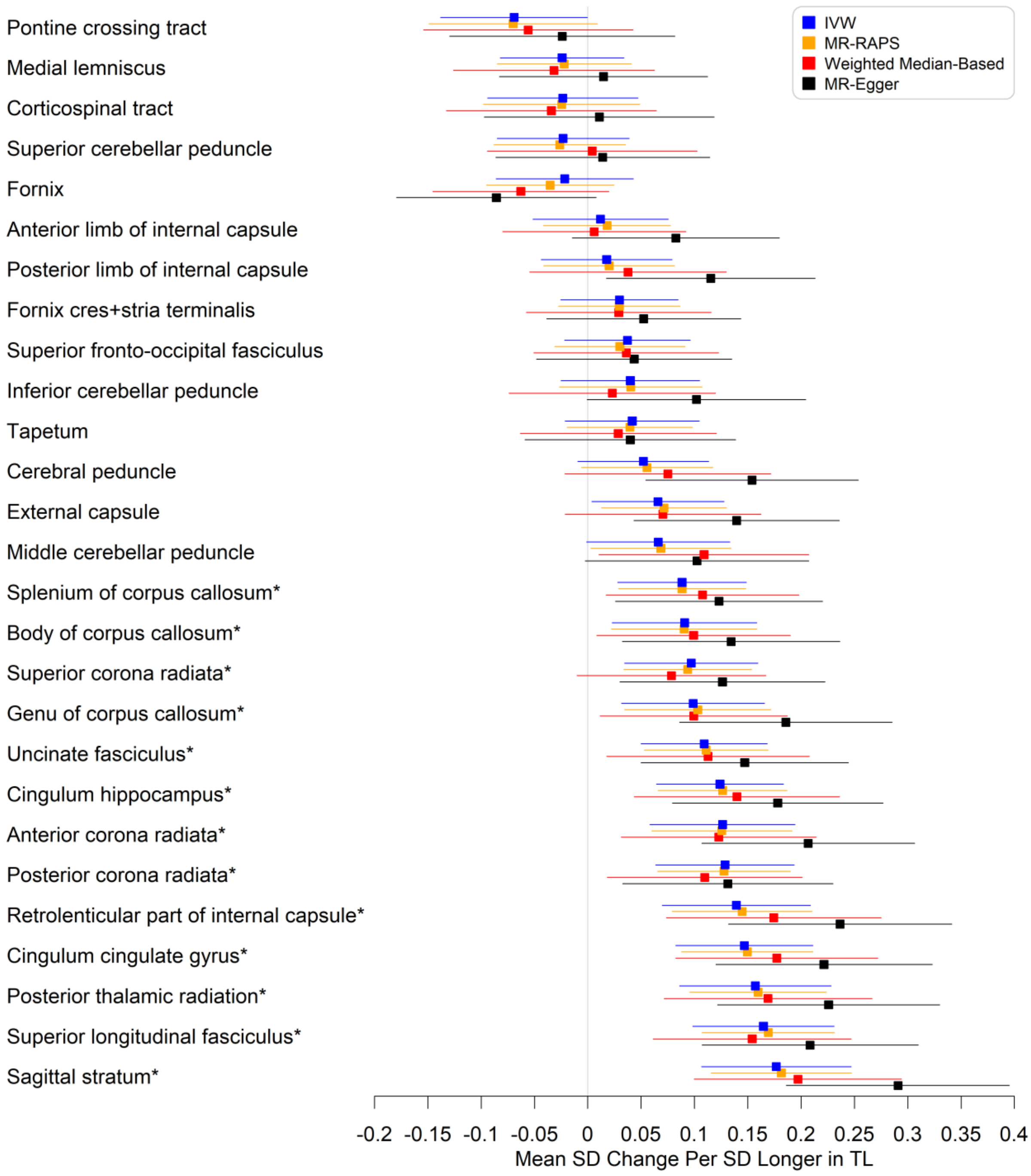


**eFigure 11.** Associations between genetically determined telomere length and weighted-mean mean diffusivity IDPs, comparing the primary (IVW: inverse variance weighted) to secondary MR methods (Mendelian randomization robust adjusted profile score (MR-RAPS); weighted median based method; MR-Egger method)

*Significant at the false discovery rate < 0.05 level using the IVW method.
