## Supplement 3 for "Mid-life leukocyte telomere length and dementia risk: a prospective cohort study of 435,046 UK Biobank participants"

**eMethods 1** | UK Biobank cognitive assessments

**eMethods 2** | Method to correct for winner's curse

**eMethods 3** | R code to correct for winner's curse

### eMethods 1. UK Biobank cognitive assessments

**Reaction time in the domain of processing speed.** The reaction time was measured as the average response time in milliseconds, used to correctly identify pairs of matching symbols across four trials. This test was administered at both the baseline and first imaging visits (n=432,183 in the baseline cohort).

**Numeric memory in the domain of working memory**. A number starting from two digits was briefly displayed on screen and then disappeared. After a short pause, participants were asked to enter their remembered number onto the screen. Each time the participant entered the correct digit sequence, the number displayed became 1-digit longer, with up to a maximum of 12 digits. Participants had up to five successive tries for 2-digit numbers and two successive tries for 3 or more-digit numbers. The score was the maximum number of digits correctly remembered. This test was administered at both the baseline and first imaging visits. Participants who chose to abandon the test were excluded from the analysis (n=968 of 45,666 in the baseline cohort).

**Fluid intelligence in the domain of verbal and numerical reasoning**. Fluid intelligence score was an unweighted sum of the number of questions answered correctly in two minutes out of thirteen questions. Test questions comprised 1) numeric addition test, 2) identifying largest number, 3) word interpolation, 4) positional arithmetic, 5) family relationship calculation, 6) conditional arithmetic, 7) synonym, chained arithmetic, 8) concept interpolation, 9) arithmetic 10) sequence recognition, 11) antonym, 12) square sequence recognition, and 13) subset inclusion logic. This test was administered at both baseline and first imaging visits. At the start of the test, participants who checked "I am not able to try this." rather than "Begin check." were excluded from analysis (n=1,856 of 141,776 in the baseline cohort).

**Prospective memory in the domain of prospective memory.** Before the start of all other cognitive tests, participants were presented with the following instruction: "At the end of the games we will show you four colored symbols and ask you to touch the Blue Square. However, to test your memory, we want you to actually touch the Orange Circle instead." Participants then proceeded to complete all other cognitive assessments. At the end of the cognitive tests, participants were shown a screen with four shapes (blue square, pink star, grey cross, orange circle) and were instructed to touch the Blue Square. Participants were scored as one if they remembered to correctly touch the orange circle on their first attempt, or zero if they failed their first attempt.

**Pairs matching in the domain of visual declarative memory.** Participants were shown a screen of six cards with three pairs of matching symbols for 3 seconds before the cards were turned over. Participants were asked to correctly identify the pairs of cards with matching symbols using the fewest number of attempts. All participants completed two trials, one with 6 cards consisting of 3 matching pairs, and another trial with 12 cards consisting of 6 matching pairs. The score for each trial was the total number of errors made during this task and we used the second trial score for analyses.

**Symbol digit substitution in the domain of processing speed**. A key with a set of eight paired symbols with numbers was presented to the participants. Underneath the key, a row of randomly shuffled symbols was then displayed, and participants were asked to select the number that paired with the corresponding symbol using the key displayed above. Once completed, a new set of symbol-number pairs appeared. A practice trial was completed before the test started. The score was the number of correct symbol-digit matches within one minute. This test was administered at the first imaging visit only (n=29,413 in the imaging cohort).

**Trail making part B in the domain of executive function**. Participants were instructed to connect nodes containing numbers 1-13 and letters A-L in alternate ascending order, with a number followed by a letter (e.g., 1 A 2 B 3 C and so on). The score was the time taken to complete each part in deciseconds. This trail making test was first administered during the first imaging visit. Participants who failed to complete the trail were excluded from analysis (n=1,130 of 29,722 in the imaging cohort).

**Matrix pattern completion in the domain of non-verbal reasoning**. Participants were presented with a matrix puzzle with a piece missing and a set of possible selection pieces at the bottom of the puzzle. The task was to select the piece which best completes the matrix design in a logical manner. Participants had two examples to practice before starting the test. There were 15 total patterns presented in ascending order of difficulty. The score was calculated as the number of correct pieces answered in three minutes with a range of 0 to15. This test was administered at the first imaging visit only (n=29,392 in the imaging cohort).

### eMethods 2. Method to correct for winner's curse

Denoted by $\hat{\beta}_j$ the discovery association estimate of the $j$-th genetic instrument with the standard error $\sigma_j$, both subject to winner's curse due to the selection of $p<8.31\times10^{-9}$ (which is equivalent to the z-test statistic threshold $\delta$=5.76). To correct for winner's curse, we maximized the likelihood penalized on the probability of being genome-wide significant:

$$logf\left(\beta_j \middle| \hat{\beta}_j, \sigma_j\right) - log\left[\Phi\left(\frac{\beta_j}{\sigma_j} - \delta\right) + \Phi\left(-\frac{\beta_j}{\sigma_j} - \delta\right)\right]$$

with respect to $\beta_j$ using numerical maximization, where $\Phi$ is the cumulative distribution function of the standard normal distribution. This method is based on the technique of Zhong and Prentice (2010), yielding a corrected point estimate and standard error for the $j$-th genetic instrument. To further improve the stability of the estimate, we replaced $\delta$ with $0.9\delta$ in this estimation to avoid excessive over-correction in a small number of cases (R code provided in Text SC.2 in supporting information).

### eMethods 3. R code to correct for winner's curse

Code requires a vector of SNP-exposure associations estimates and their standard errors (`BetaXG' and `seBetaXG') and a p-value threshold 'p'. It will produce a vector of corrected estimates and standard errors BetaXGc and seBetaXGc. Note the code has a stability parameter S. Ideally, it would be 1, you will need to play around with it to see how close you can get it to 1 without getting a NaN in the standard errors.

**Example input data:**

```
BetaXG <- c(0.091,0.018,0.026)

seBetaXG <- c(0.015051297,0.002040854,0.004081708)

wc_corrected <- function(BetaXG,seBetaXG){

Isq = function(y,s){

 k = length(y)

 w = 1/s^2; sum.w = sum(w)

 mu.hat = sum(y*w)/sum.w

 Q = sum(w*(y-mu.hat)^2)

 Isq = (Q - (k-1))/Q

 Isq = max(0,Isq)

 return(Isq)

}

S = 0.9

p = 0.00000000831

t = qnorm(1-p/2)

L = length(BetaXG)

BetaXGc = NULL

seBetaXGc = NULL
```

```
Wcorrect = function(a){
  gamma = a[1]
  f1 = dnorm(b,gamma,se)
  f2 = pnorm((gamma/se) - (S*t))
  l = -log(f1/f2)
}
for(i in 1:L){
  b = BetaXG[i]
  se = seBetaXG[i]
  results = optim(b, Wcorrect,method = "BFGS", hessian = TRUE)
  BetaXGc[i] = results$par
  seBetaXGc[i] = sqrt(1/results$hessian)
}
F_original = BetaXG^2/seBetaXG^2; mF_original = mean(BetaXG^2/seBetaXG^2)
F_wc = BetaXGc^2/seBetaXGc^2; mF_wc = mean(BetaXGc^2/seBetaXGc^2)
I2_original = Isq(BetaXG,seBetaXG)
I2_wc = Isq(BetaXGc,seBetaXGc)
Data = data.frame(BetaXG, seBetaXG, BetaXGc, seBetaXGc, F_original, F_wc)
FIstats = c(mF_original, mF_wc, I2_original, I2_wc)
return(Data)
}
```
