## Supplement 4 for "Mid-life leukocyte telomere length and dementia risk: a prospective cohort study of 435,046 UK Biobank participants"

**eFigure 1** | SNP-AD/ADRD association plotted against SNP-telomere length association, labelled by the mapped gene, with MR slope estimates shown

**eFigure 2** | SNP-AD association plotted against SNP-telomere length association, labelled by the mapped gene, with MR slope estimates shown

**eFigure 3** | SNP-vascular dementia association plotted against SNP-telomere length association, labelled by the mapped gene, with MR slope estimates shown

**eFigure 4** | SNP-general cognitive ability association plotted against SNP-telomere length association, labelled by the mapped gene, with MR slope estimates shown

**eFigure 5** | SNP-reaction time association plotted against SNP-telomere length association, labelled by the mapped gene, with MR slope estimates shown

**eFigure 6** | SNP-fluid intelligence association plotted against SNP-telomere length association, labelled by the mapped gene, with MR slope estimates shown

**eFigure 7** | SNP-numeric memory association plotted against SNP-telomere length association, labelled by the mapped gene, with MR slope estimates shown

**eFigure 8** | SNP-symbol digit substitution association plotted against SNP-telomere length association, labelled by the mapped gene, with MR slope estimates shown

**eFigure 9** | SNP-trail making part B association plotted against SNP-telomere length association, labelled by the mapped gene, with MR slope estimates shown

**eFigure 10** | SNP-matrix pattern completion association plotted against SNP-telomere length association, labelled by the mapped gene, with MR slope estimates shown

**eFigure 11** | SNP-total volume of white matter hyperintensities association plotted against SNP-telomere length association, labelled by the mapped gene, with MR slope estimates shown

**eFigure 12** | SNP-cuneus volume association plotted against SNP-telomere length association, labelled by the mapped gene, with MR slope estimates shown

**eFigure 13** | SNP-entorhinal volume association plotted against SNP-telomere length association, labelled by the mapped gene, with MR slope estimates shown

**eFigure 14** | SNP-inferiorparietal volume association plotted against SNP-telomere length association, labelled by the mapped gene, with MR slope estimates shown

**eFigure 15** | SNP-parahippocampal volume association plotted against SNP-telomere length association, labelled by the mapped gene, with MR slope estimates shown

**eFigure 16** | SNP-precuneus volume association plotted against SNP-telomere length association, labelled by the mapped gene, with MR slope estimates shown

**eFigure 17** | SNP-hippocampus volume association plotted against SNP-telomere length association, labelled by the mapped gene, with MR slope estimates shown

**eFigure 18** | SNP-mean FA in anterior corona radiata association plotted against SNP-telomere length association, labelled by the mapped gene, with MR slope estimates shown

**eFigure 19** | SNP-mean FA in anterior limb of internal capsule association plotted against SNP-telomere length association, labelled by the mapped gene, with MR slope estimates shown

**eFigure 20** | SNP-mean FA in body of corpus callosum association plotted against SNP-telomere length association, labelled by the mapped gene, with MR slope estimates shown

**eFigure 21** | SNP-mean FA in cerebral peduncle association plotted against SNP-telomere length association, labelled by the mapped gene, with MR slope estimates shown

**eFigure 22** | SNP-mean FA in cingulum cingulate gyrus association plotted against SNP-telomere length association, labelled by the mapped gene, with MR slope estimates shown

**eFigure 23** | SNP-mean FA in cingulum hippocampus association plotted against SNP-telomere length association, labelled by the mapped gene, with MR slope estimates shown

**eFigure 24** | SNP-mean FA in corticospinal tract association plotted against SNP-telomere length association, labelled by the mapped gene, with MR slope estimates shown

**eFigure 25** | SNP-mean FA in external capsule association plotted against SNP-telomere length association, labelled by the mapped gene, with MR slope estimates shown

**eFigure 26** | SNP-mean FA in fornix association plotted against SNP-telomere length association, labelled by the mapped gene, with MR slope estimates shown

**eFigure 27** | SNP-mean FA in fornix cres+stria terminalis association plotted against SNP-telomere length association, labelled by the mapped gene, with MR slope estimates shown

**eFigure 28** | SNP-mean FA in genu of corpus callosum association plotted against SNP-telomere length association, labelled by the mapped gene, with MR slope estimates shown

**eFigure 29** | SNP-mean FA in inferior cerebellar peduncle association plotted against SNP-telomere length association, labelled by the mapped gene, with MR slope estimates shown

**eFigure 30** | SNP-mean FA in medial lemniscus association plotted against SNP-telomere length association, labelled by the mapped gene, with MR slope estimates shown

**eFigure 31** | SNP-mean FA in middle cerebellar peduncle association plotted against SNP-telomere length association, labelled by the mapped gene, with MR slope estimates shown

**eFigure 32** | SNP-mean FA in pontine crossing tract association plotted against SNP-telomere length association, labelled by the mapped gene, with MR slope estimates shown

**eFigure 33** | SNP-mean FA in posterior corona radiata association plotted against SNP-telomere length association, labelled by the mapped gene, with MR slope estimates shown

**eFigure 34** | SNP-mean FA in posterior limb of internal capsule association plotted against SNP-telomere length association, labelled by the mapped gene, with MR slope estimates shown

**eFigure 35** | SNP-mean FA in posterior thalamic radiation association plotted against SNP-telomere length association, labelled by the mapped gene, with MR slope estimates shown

**eFigure 36** | SNP-mean FA in retrolenticular part of internal capsule association plotted against SNP-telomere length association, labelled by the mapped gene, with MR slope estimates shown

**eFigure 37** | SNP-mean FA in sagittal stratum association plotted against SNP-telomere length association, labelled by the mapped gene, with MR slope estimates shown

**eFigure 38** | SNP-mean FA in splenium of corpus callosum association plotted against SNP-telomere length association, labelled by the mapped gene, with MR slope estimates shown

**eFigure 39** | SNP-mean FA in superior cerebellar peduncle association plotted against SNP-telomere length association, labelled by the mapped gene, with MR slope estimates shown

**eFigure 40** | SNP-mean FA in superior corona radiata association plotted against SNP-telomere length association, labelled by the mapped gene, with MR slope estimates shown

**eFigure 41** | SNP-mean FA in superior fronto-occipital fasciculus association plotted against SNP-telomere length association, labelled by the mapped gene, with MR slope estimates shown

**eFigure 42** | SNP-mean FA in superior longitudinal fasciculus association plotted against SNP-telomere length association, labelled by the mapped gene, with MR slope estimates shown

**eFigure 43** | SNP-mean FA in tapetum association plotted against SNP-telomere length association, labelled by the mapped gene, with MR slope estimates shown

**eFigure 44** | SNP-mean FA in uncinate fasciculus association plotted against SNP-telomere length association, labelled by the mapped gene, with MR slope estimates shown

**eFigure 45** | SNP-mean MD in anterior corona radiata association plotted against SNP-telomere length association, labelled by the mapped gene, with MR slope estimates shown

**eFigure 46** | SNP-mean MD in anterior limb of internal capsule association plotted against SNP-telomere length association, labelled by the mapped gene, with MR slope estimates shown

**eFigure 47** | SNP-mean MD in body of corpus callosum association plotted against SNP-telomere length association, labelled by the mapped gene, with MR slope estimates shown

**eFigure 48** | SNP-mean MD in cerebral peduncle association plotted against SNP-telomere length association, labelled by the mapped gene, with MR slope estimates shown

**eFigure 49** | SNP-mean MD in cingulum cingulate gyrus association plotted against SNP-telomere length association, labelled by the mapped gene, with MR slope estimates shown

**eFigure 50** | SNP-mean MD in cingulum hippocampus association plotted against SNP-telomere length association, labelled by the mapped gene, with MR slope estimates shown

**eFigure 51** | SNP-mean MD in corticospinal tract association plotted against SNP-telomere length association, labelled by the mapped gene, with MR slope estimates shown

**eFigure 52** | SNP-mean MD in external capsule association plotted against SNP-telomere length association, labelled by the mapped gene, with MR slope estimates shown

**eFigure 53** | SNP-mean MD in fornix association plotted against SNP-telomere length association, labelled by the mapped gene, with MR slope estimates shown

**eFigure 54** | SNP-mean MD in fornix cres+stria terminalis association plotted against SNP-telomere length association, labelled by the mapped gene, with MR slope estimates shown

**eFigure 55** | SNP-mean MD in genu of corpus callosum association plotted against SNP-telomere length association, labelled by the mapped gene, with MR slope estimates shown

**eFigure 56** | SNP-mean MD in inferior cerebellar peduncle association plotted against SNP-telomere length association, labelled by the mapped gene, with MR slope estimates shown

**eFigure 57** | SNP-mean MD in medial lemniscus association plotted against SNP-telomere length association, labelled by the mapped gene, with MR slope estimates shown

**eFigure 58** | SNP-mean MD in middle cerebellar peduncle association plotted against SNP-telomere length association, labelled by the mapped gene, with MR slope estimates shown

**eFigure 59** | SNP-mean MD in pontine crossing tract association plotted against SNP-telomere length association, labelled by the mapped gene, with MR slope estimates shown

**eFigure 60** | SNP-mean MD in posterior corona radiata association plotted against SNP-telomere length association, labelled by the mapped gene, with MR slope estimates shown

**eFigure 61** | SNP-mean MD in posterior limb of internal capsule association plotted against SNP-telomere length association, labelled by the mapped gene, with MR slope estimates shown

**eFigure 62** | SNP-mean MD in posterior thalamic radiation association plotted against SNP-telomere length association, labelled by the mapped gene, with MR slope estimates shown

**eFigure 63** | SNP-mean MD in retrolenticular part of internal capsule association plotted against SNP-telomere length association, labelled by the mapped gene, with MR slope estimates shown

**eFigure 64** | SNP-mean MD in sagittal stratum association plotted against SNP-telomere length association, labelled by the mapped gene, with MR slope estimates shown

**eFigure 65** | SNP-mean MD in splenium of corpus callosum association plotted against SNP-telomere length association, labelled by the mapped gene, with MR slope estimates shown

**eFigure 66** | SNP-mean MD in superior cerebellar peduncle association plotted against SNP-telomere length association, labelled by the mapped gene, with MR slope estimates shown

**eFigure 67** | SNP-mean MD in superior corona radiata association plotted against SNP-telomere length association, labelled by the mapped gene, with MR slope estimates shown

**eFigure 68** | SNP-mean MD in superior fronto-occipital fasciculus association plotted against SNP-telomere length association, labelled by the mapped gene, with MR slope estimates shown

**eFigure 69** | SNP-mean MD in superior longitudinal fasciculus association plotted against SNP-telomere length association, labelled by the mapped gene, with MR slope estimates shown

**eFigure 70** | SNP-mean MD in tapetum association plotted against SNP-telomere length association, labelled by the mapped gene, with MR slope estimates shown

**eFigure 71** | SNP-mean MD in uncinate fasciculus association plotted against SNP-telomere length association, labelled by the mapped gene, with MR slope estimates shown

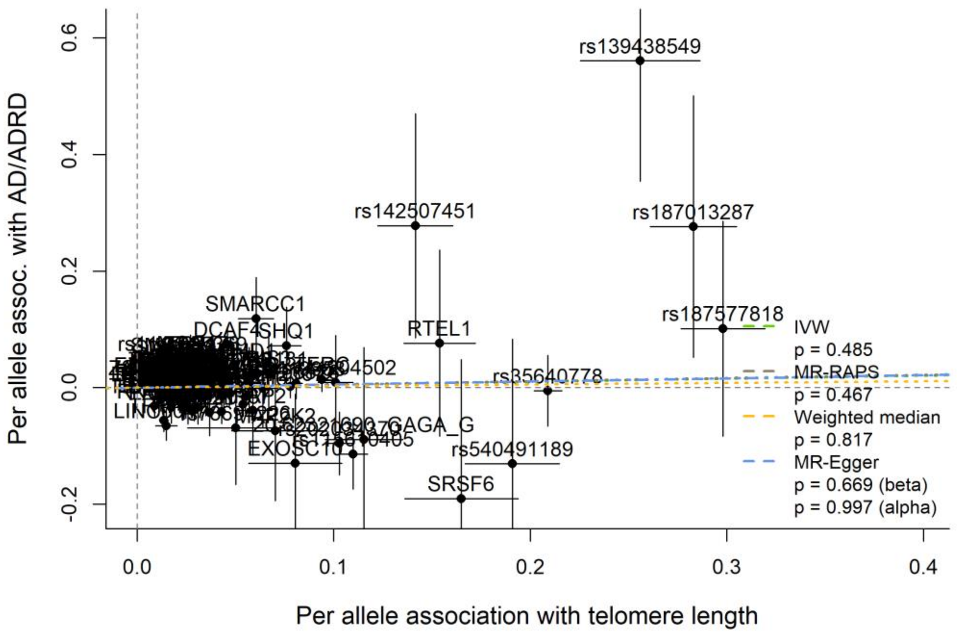


**eFigure 1.** SNP-AD/ADRD association plotted against SNP-telomere length association, labelled by the mapped gene, with MR slope estimates shown

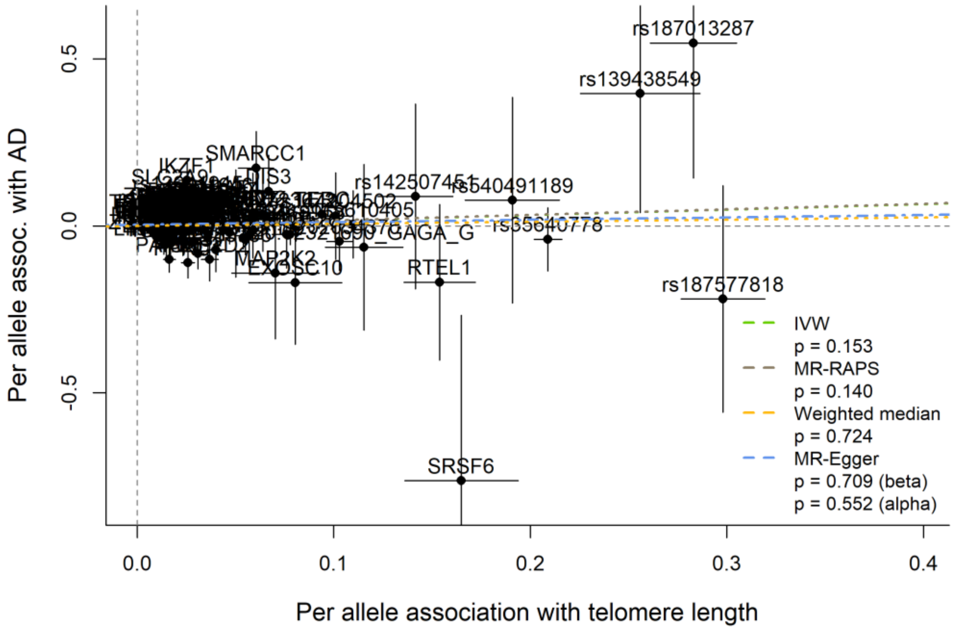


**eFigure 2.** SNP-AD association plotted against SNP-telomere length association , labelled by the mapped gene, with MR slope estimates shown

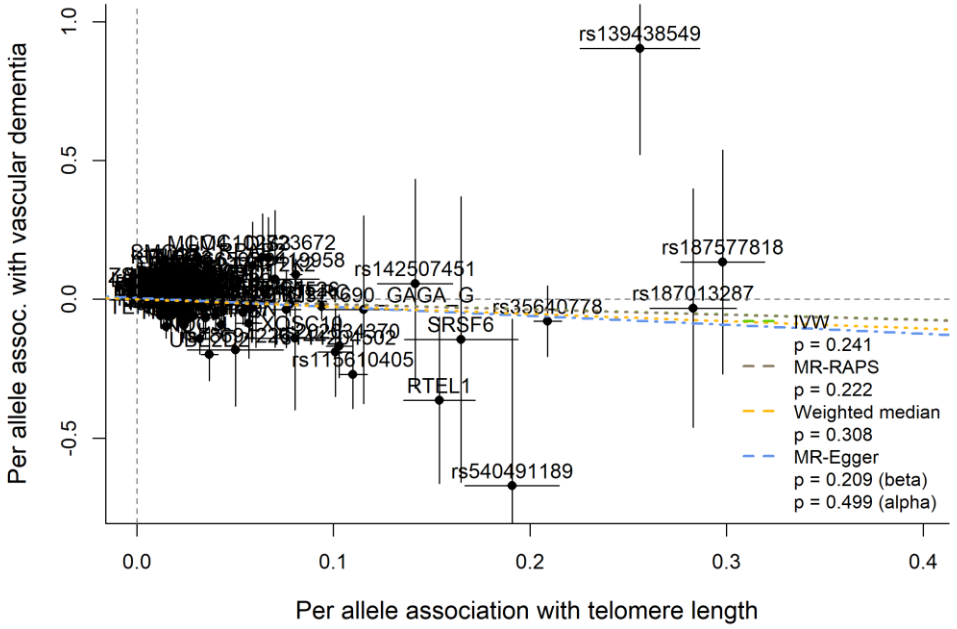


**eFigure 3.** SNP-vascular dementia association plotted against SNP-telomere length association, labelled by the mapped gene, with MR slope estimates shown

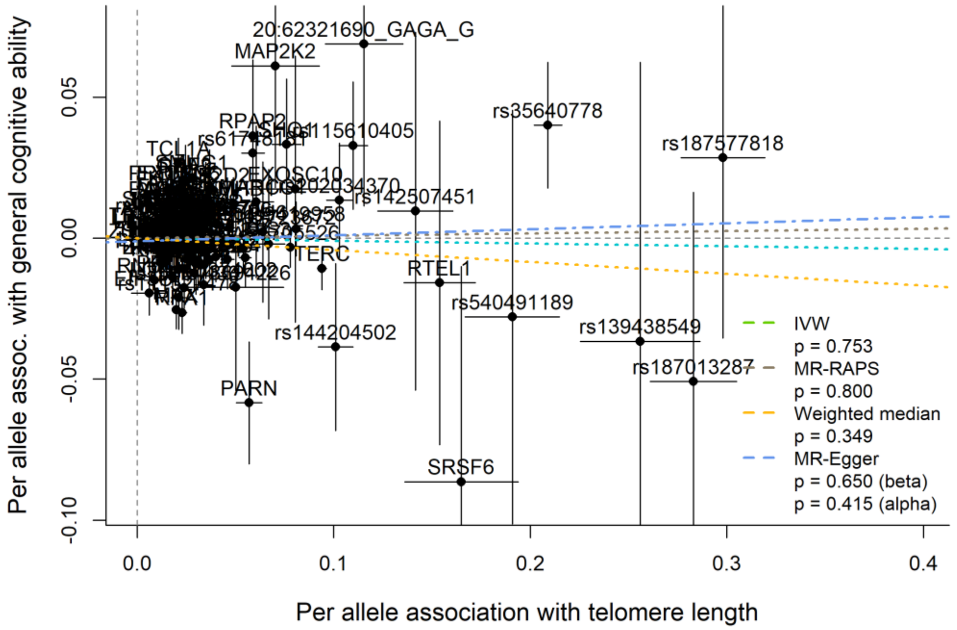


**eFigure 4.** SNP-general cognitive ability association plotted against SNP-telomere length association, labelled by the mapped gene, with MR slope estimates shown

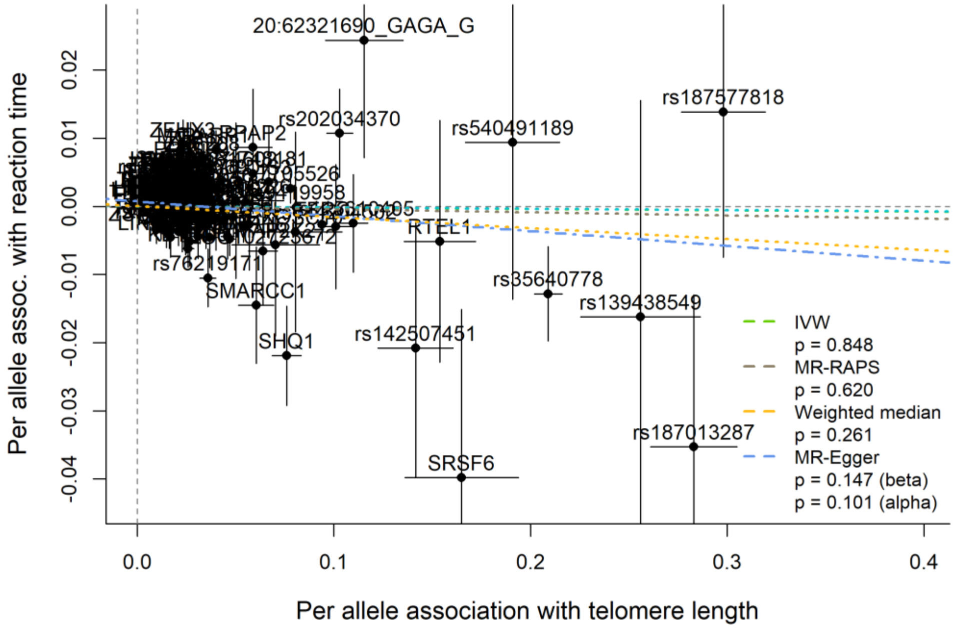


**eFigure 5.** SNP-reaction time association plotted against SNP-telomere length association, labelled by the mapped gene, with MR slope estimates shown

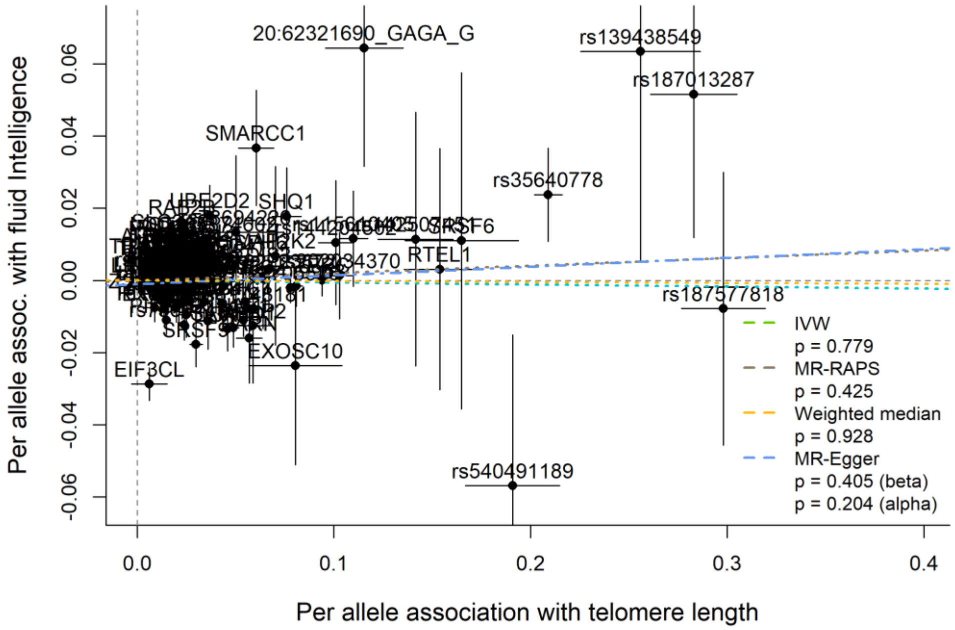


**eFigure 6.** SNP-fluid intelligence association plotted against SNP-telomere length association, labelled by the mapped gene, with MR slope estimates shown

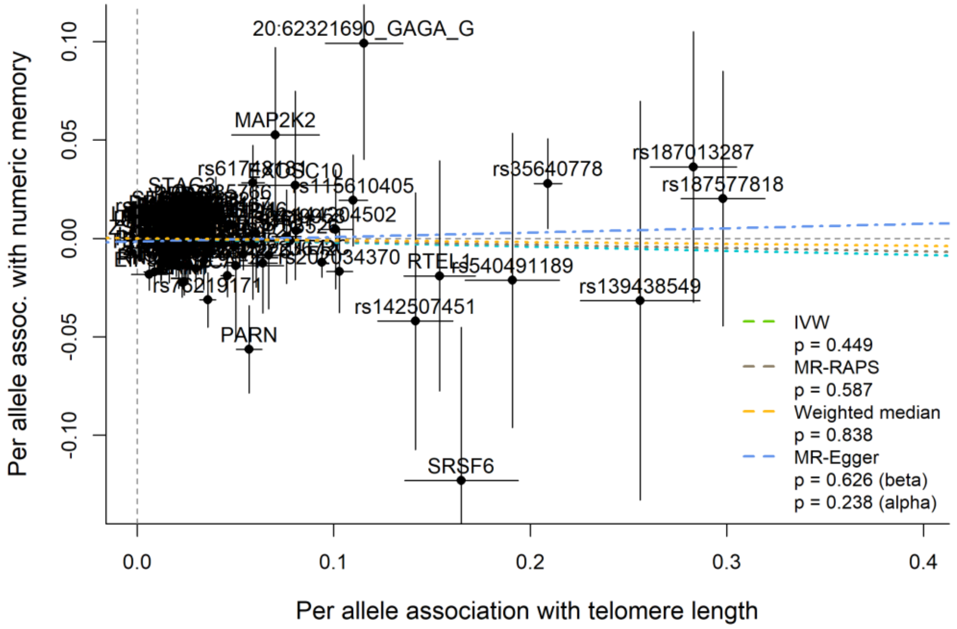


**eFigure 7.** SNP-numeric memory association plotted against SNP-telomere length association, labelled by the mapped gene, with MR slope estimates shown

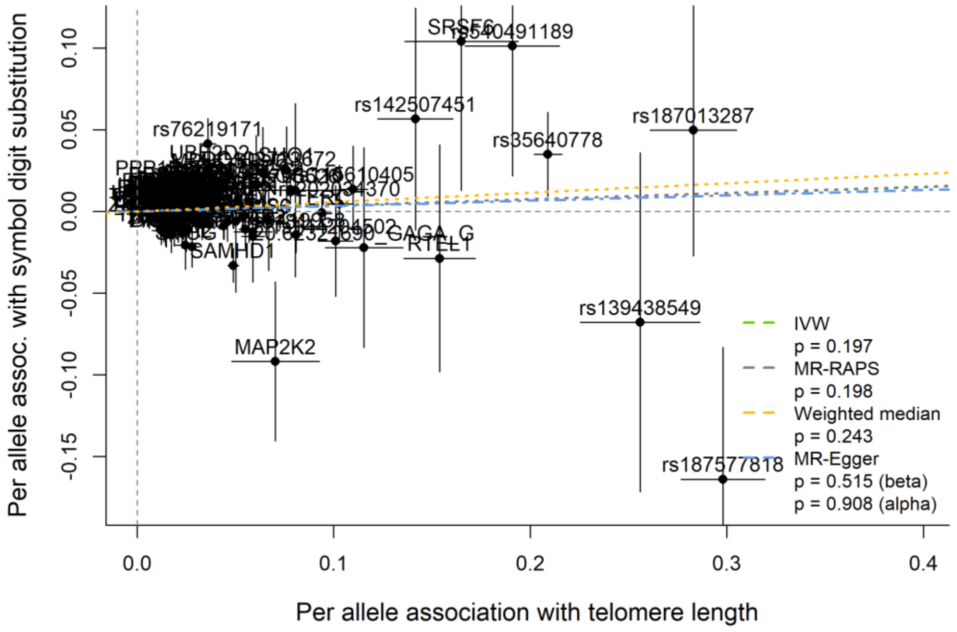


**eFigure 8.** SNP-symbol digit substitution association plotted against SNP-telomere length association, labelled by the mapped gene, with MR slope estimates shown

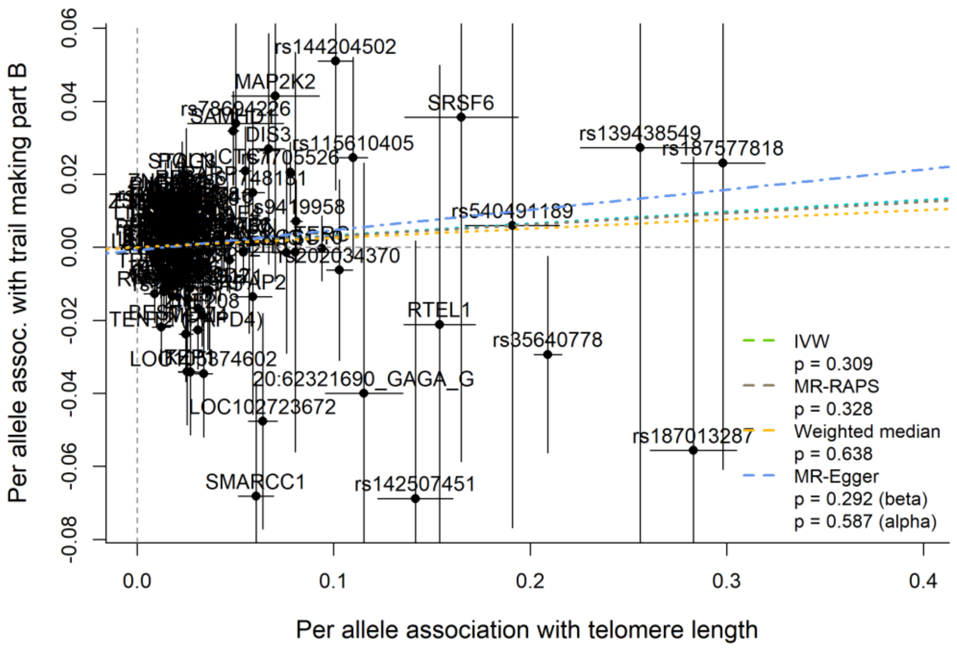


**eFigure 9.** SNP-trail making part B association plotted against SNP-telomere length association, labelled by the mapped gene, with MR slope estimates shown

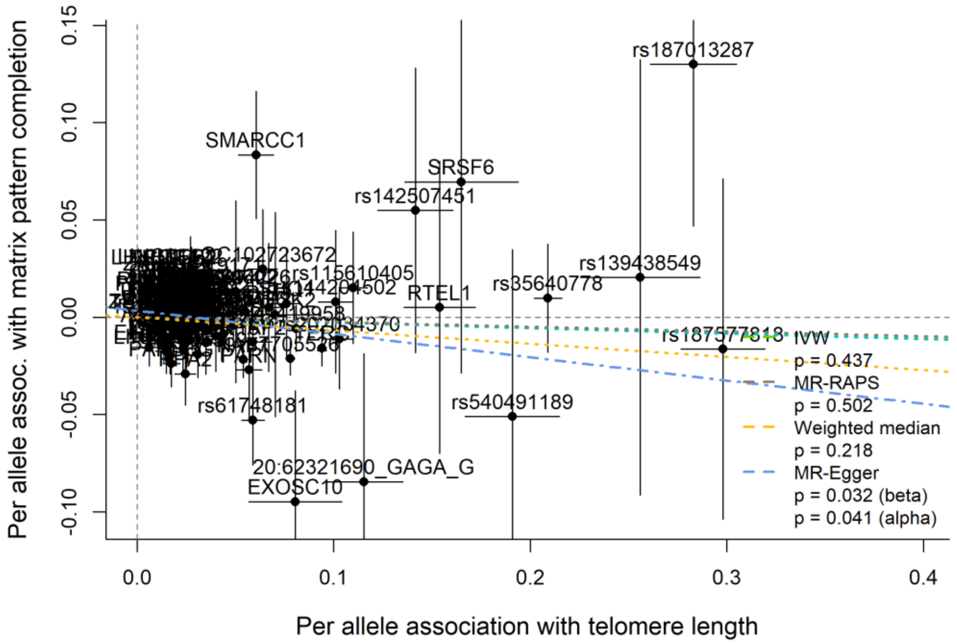


**eFigure 10.** SNP-matrix pattern completion association plotted against SNP-telomere length association, labelled by the mapped gene, with MR slope estimates shown

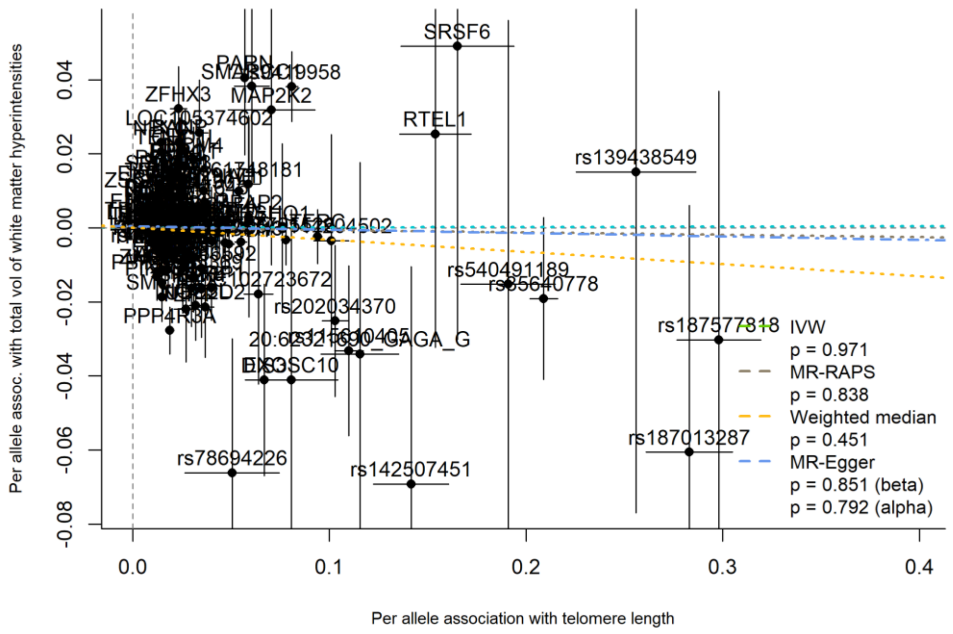


**eFigure 11.** SNP-total volume of white matter hyperintensities association plotted against SNP-telomere length association, labelled by the mapped gene, with MR slope estimates shown

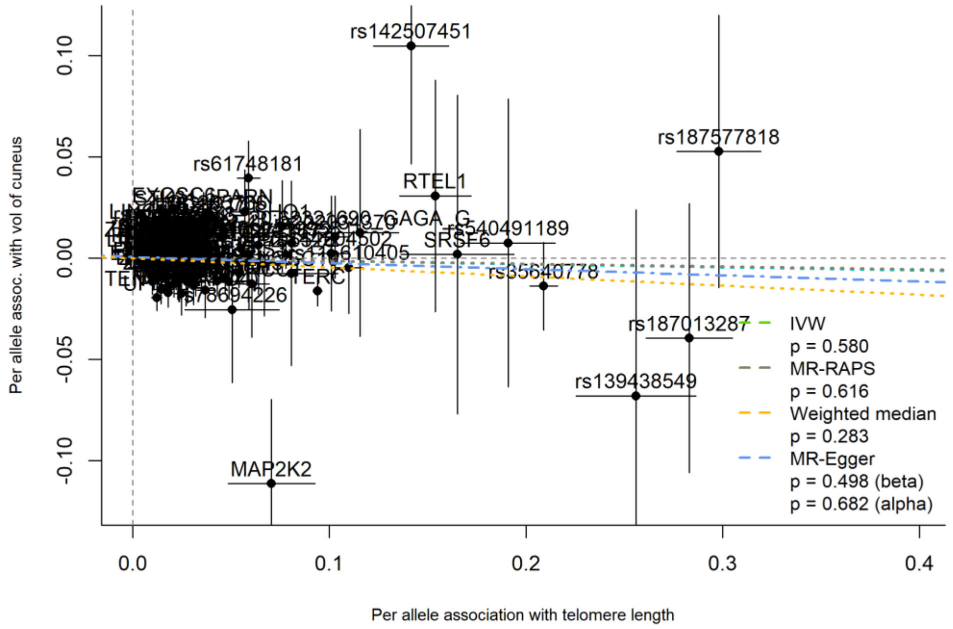


**eFigure 12.** SNP-cuneus volume association plotted against SNP-telomere length association, labelled by the mapped gene, with MR slope estimates shown

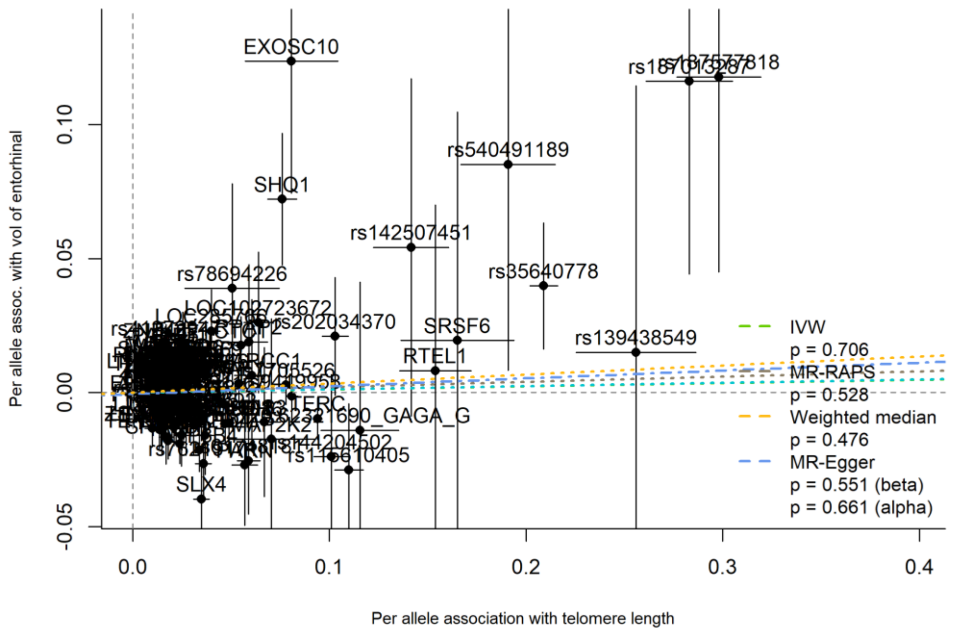


**eFigure 13.** SNP-entorhinal volume association plotted against SNP-telomere length association, labelled by the mapped gene, with MR slope estimates shown

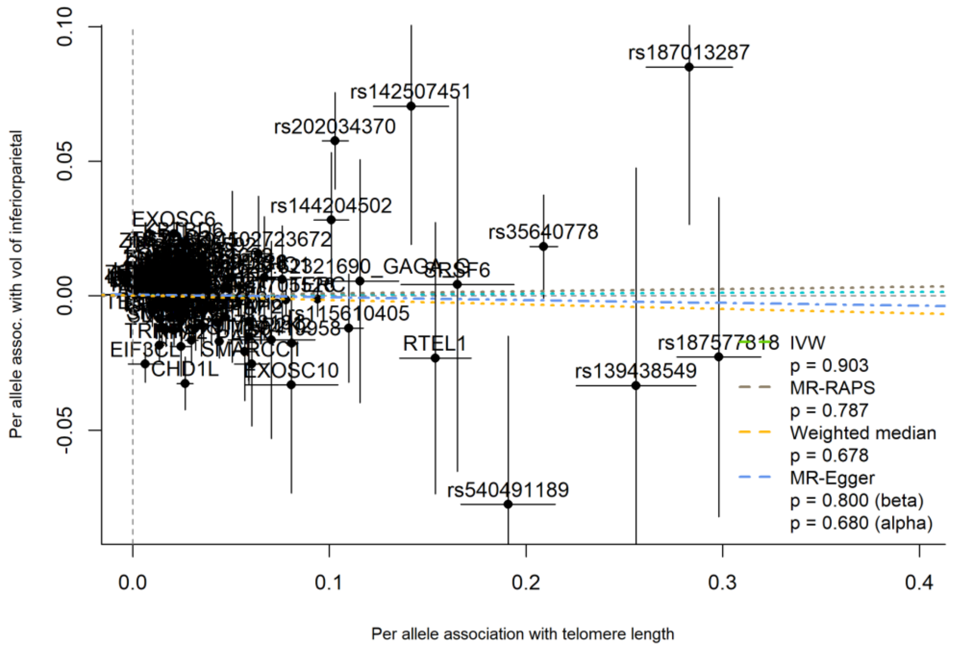


**eFigure 14.** SNP-inferiorparietal volume association plotted against SNP-telomere length association, labelled by the mapped gene, with MR slope estimates shown

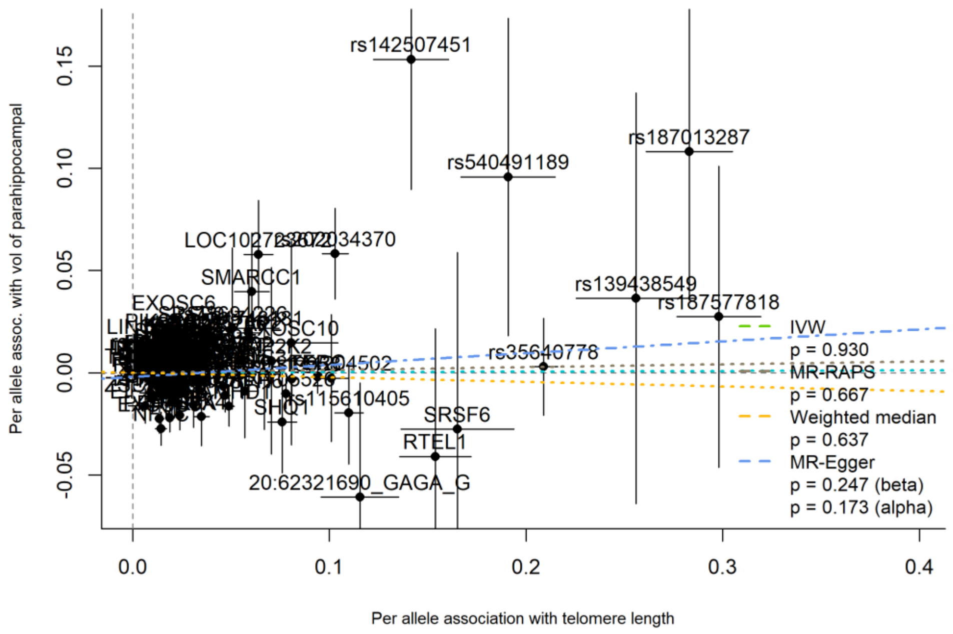


**eFigure 15.** SNP-parahippocampal volume association plotted against SNP-telomere length association, labelled by the mapped gene, with MR slope estimates shown

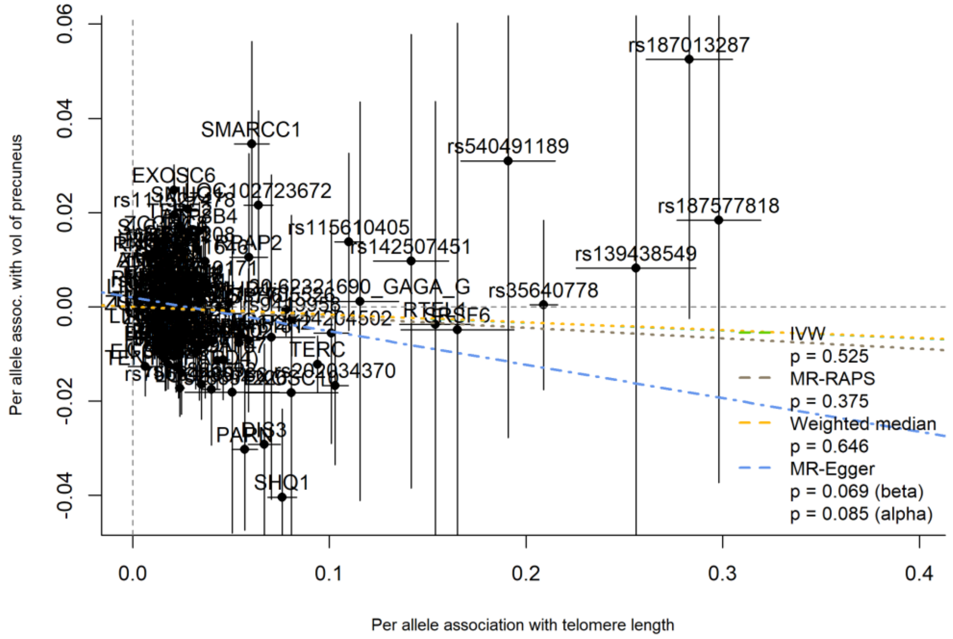


**eFigure 16.** SNP-precuneus volume association plotted against SNP-telomere length association, labelled by the mapped gene, with MR slope estimates shown

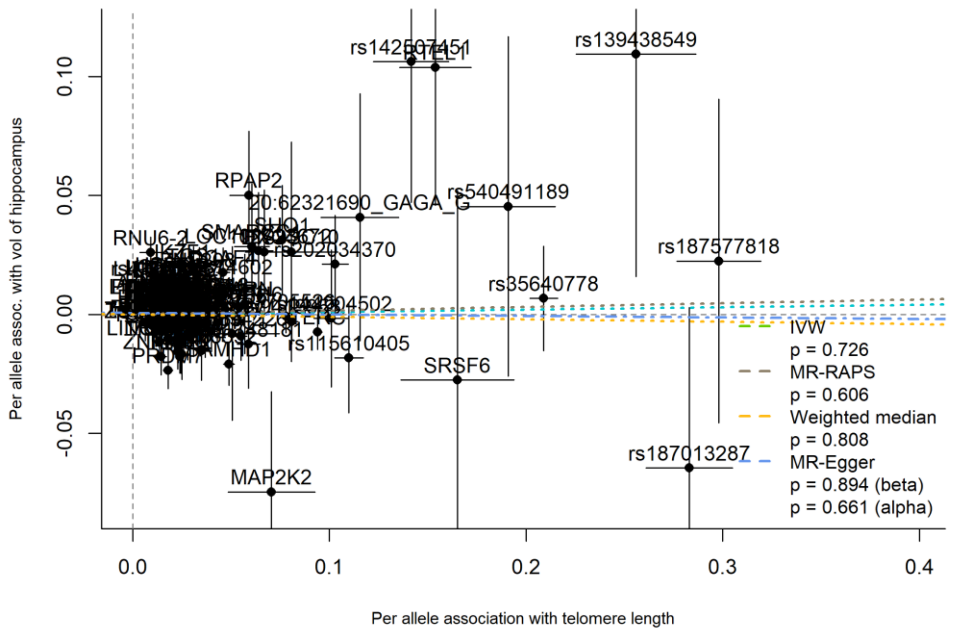


**eFigure 17.** SNP-hippocampus volume association plotted against SNP-telomere length association, labelled by the mapped gene, with MR slope estimates shown

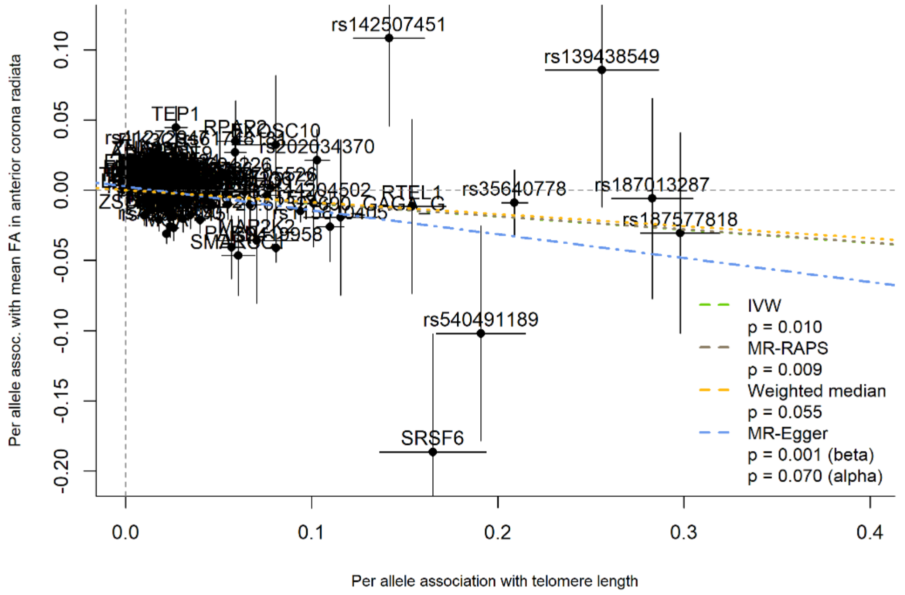


**eFigure 18.** SNP-mean FA in anterior corona radiata association plotted against SNP-telomere length association, labelled by the mapped gene, with MR slope estimates shown

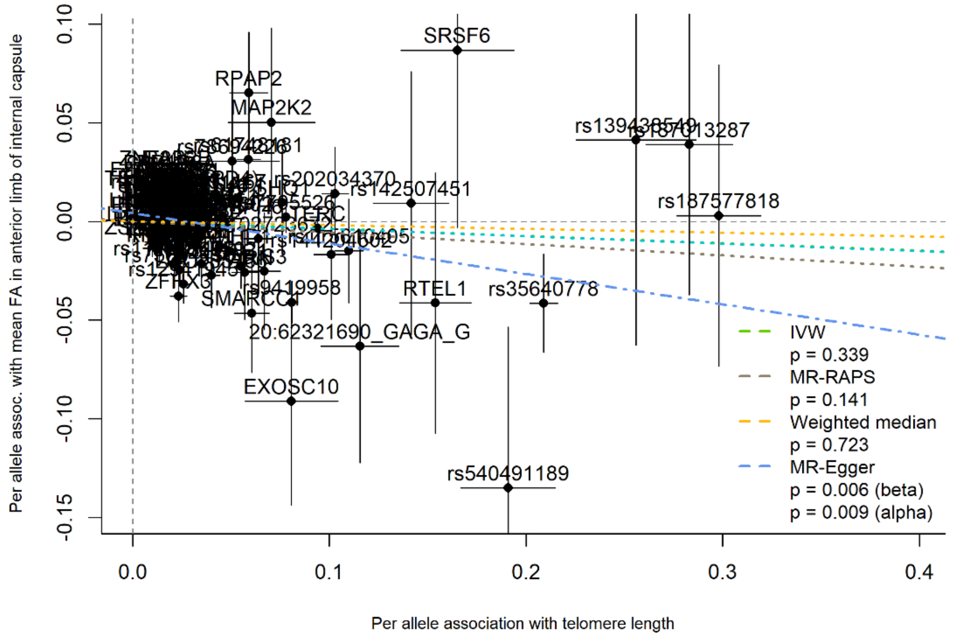


**eFigure 19.** SNP-mean FA in anterior limb of internal capsule association plotted against SNP-telomere length association, labelled by the mapped gene, with MR slope estimates shown

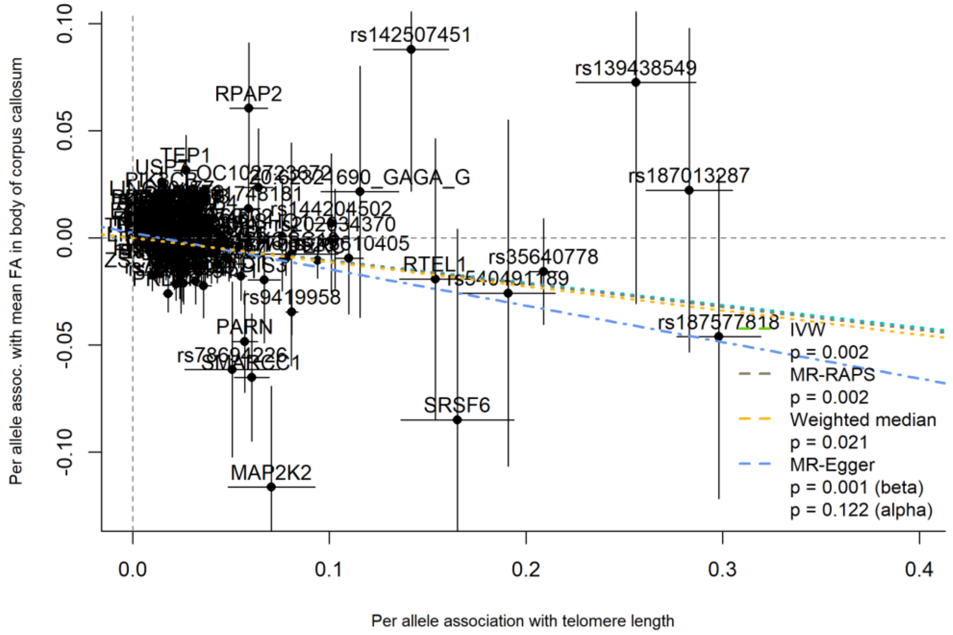


**eFigure 20.** SNP-mean FA in body of corpus callosum association plotted against SNP-telomere length association, labelled by the mapped gene, with MR slope estimates shown

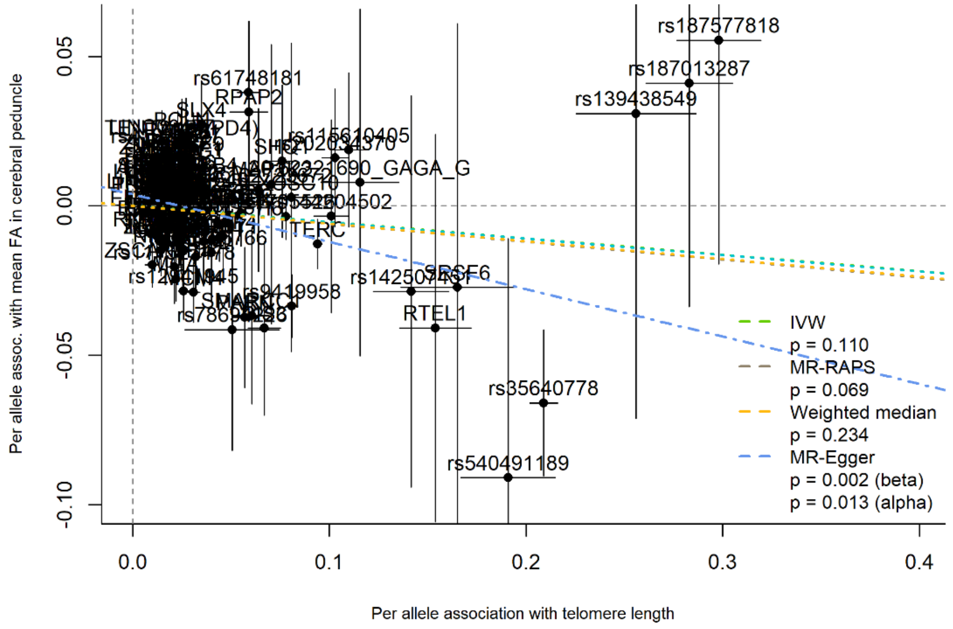


**eFigure 21.** SNP-mean FA in cerebral peduncle association plotted against SNP-telomere length association, labelled by the mapped gene, with MR slope estimates shown

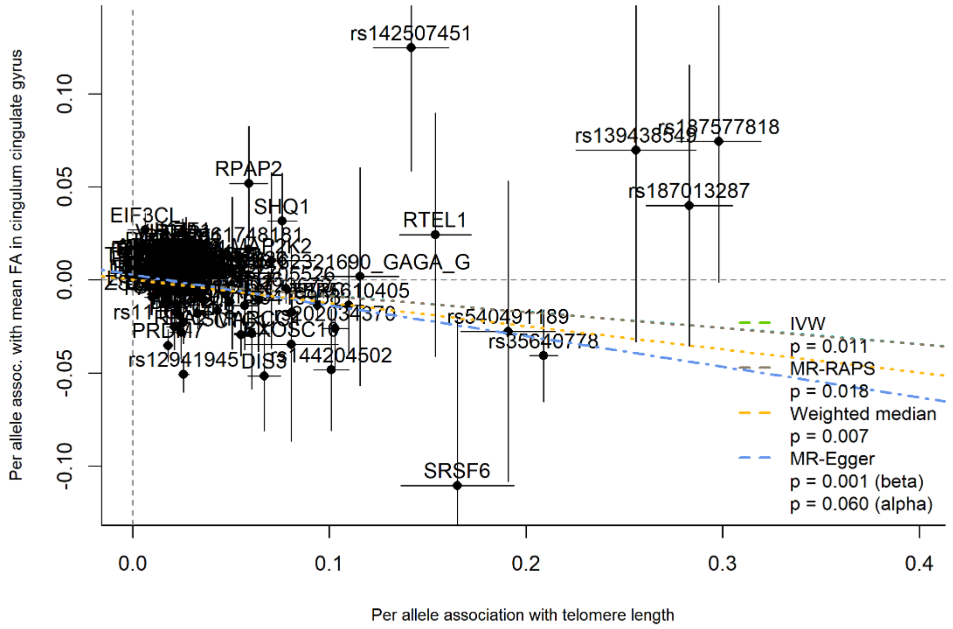


**eFigure 22.** SNP-mean FA in cingulum cingulate gyrus association plotted against SNP-telomere length association, labelled by the mapped gene, with MR slope estimates shown

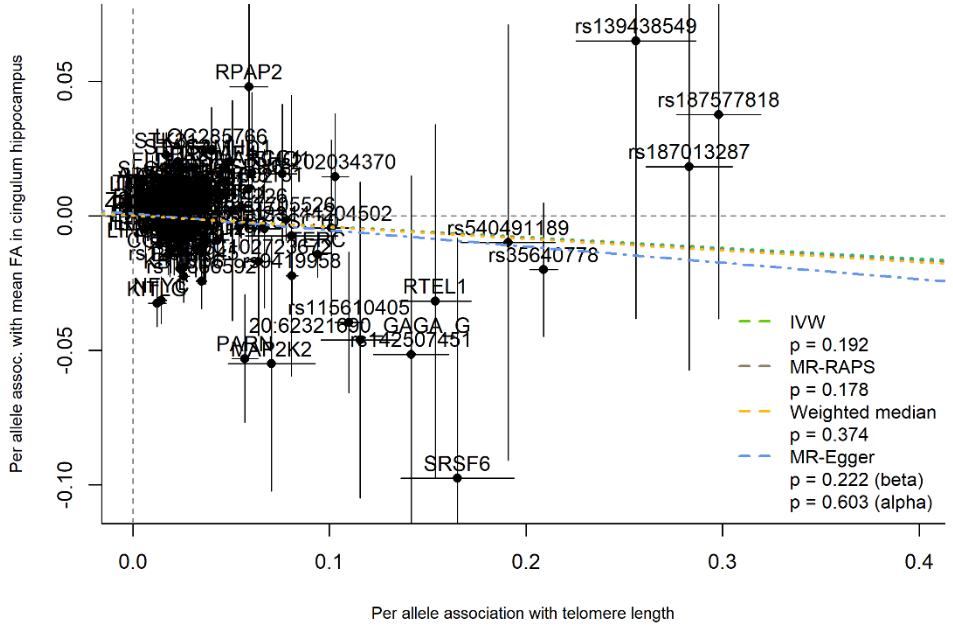


**eFigure 23.** SNP-mean FA in cingulum hippocampus association plotted against SNP-telomere length association, labelled by the mapped gene, with MR slope estimates shown

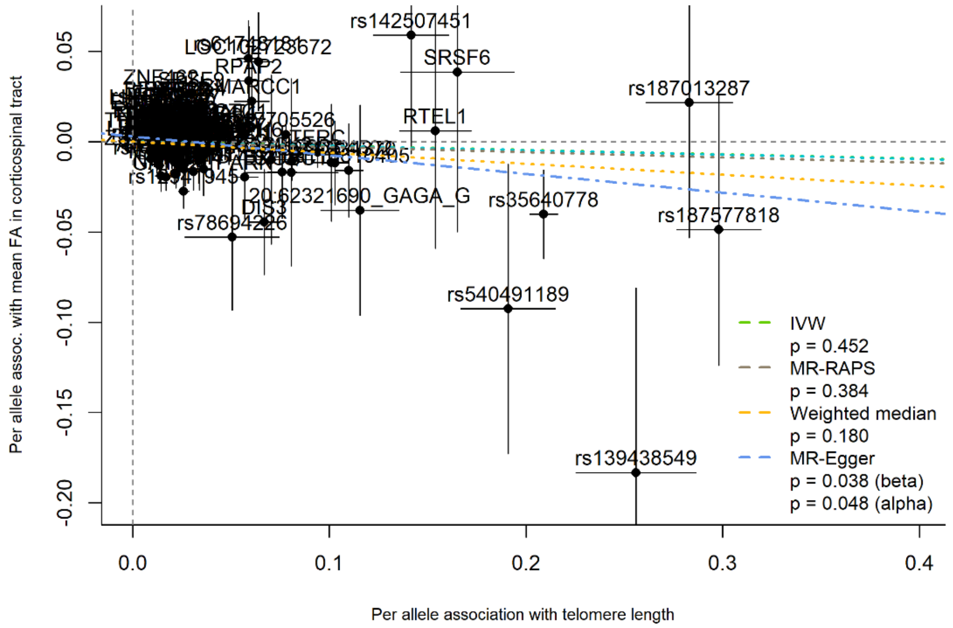


**eFigure 24.** SNP-mean FA in corticospinal tract association plotted against SNP-telomere length association, labelled by the mapped gene, with MR slope estimates shown

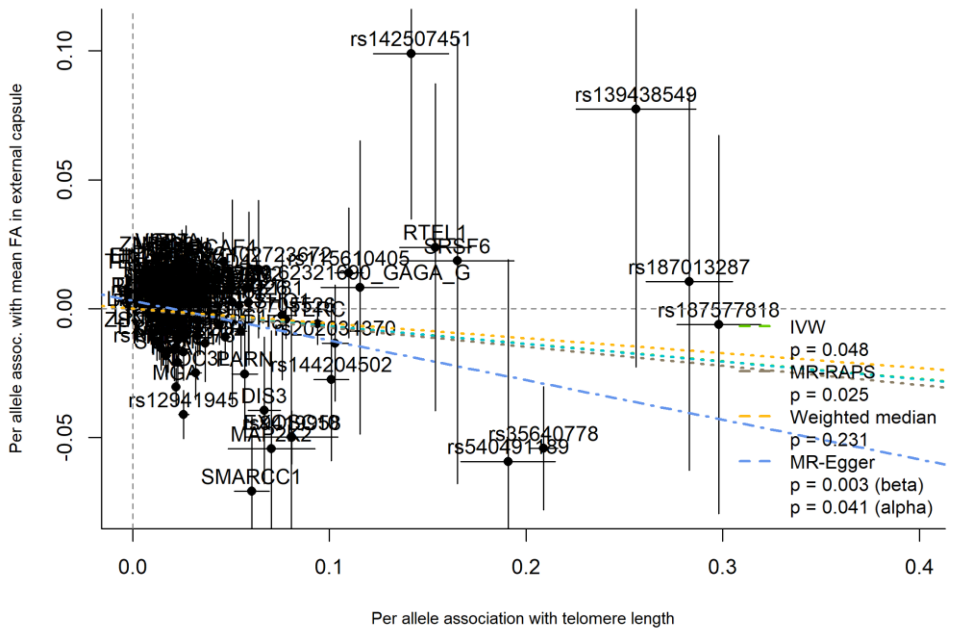


**eFigure 25.** SNP-mean FA in external capsule association plotted against SNP-telomere length association, labelled by the mapped gene, with MR slope estimates shown

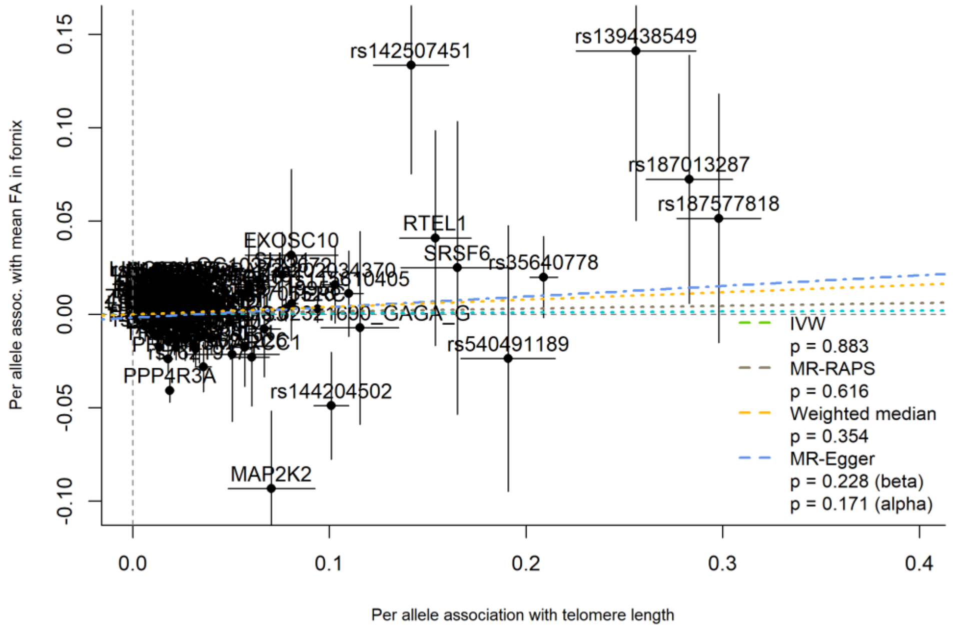


**eFigure 26.** SNP-mean FA in fornix association plotted against SNP-telomere length association, labelled by the mapped gene, with MR slope estimates shown

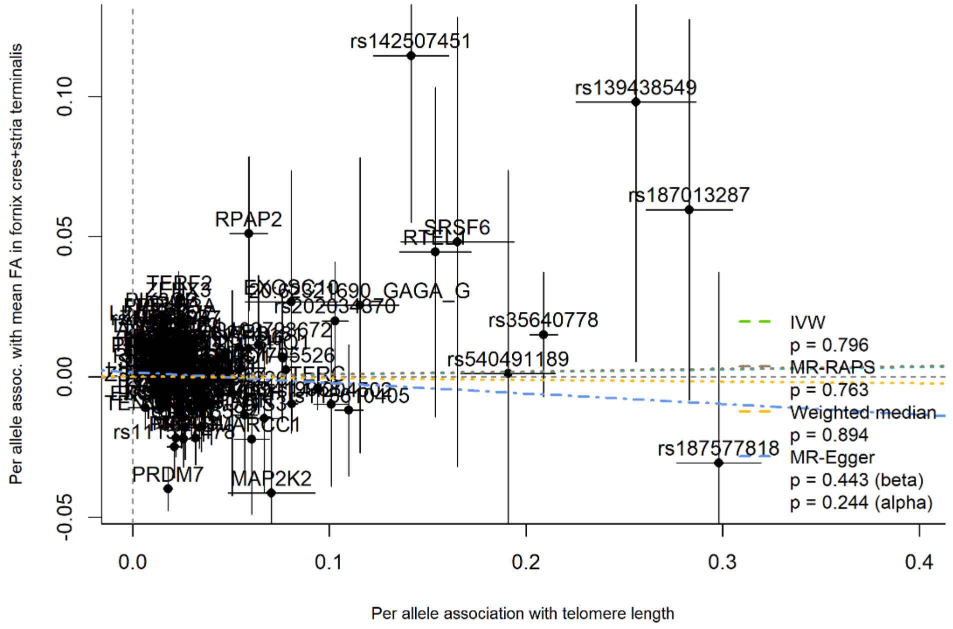


**eFigure 27.** SNP-mean FA in fornix cres+stria terminalis association plotted against SNP-telomere length association, labelled by the mapped gene, with MR slope estimates shown

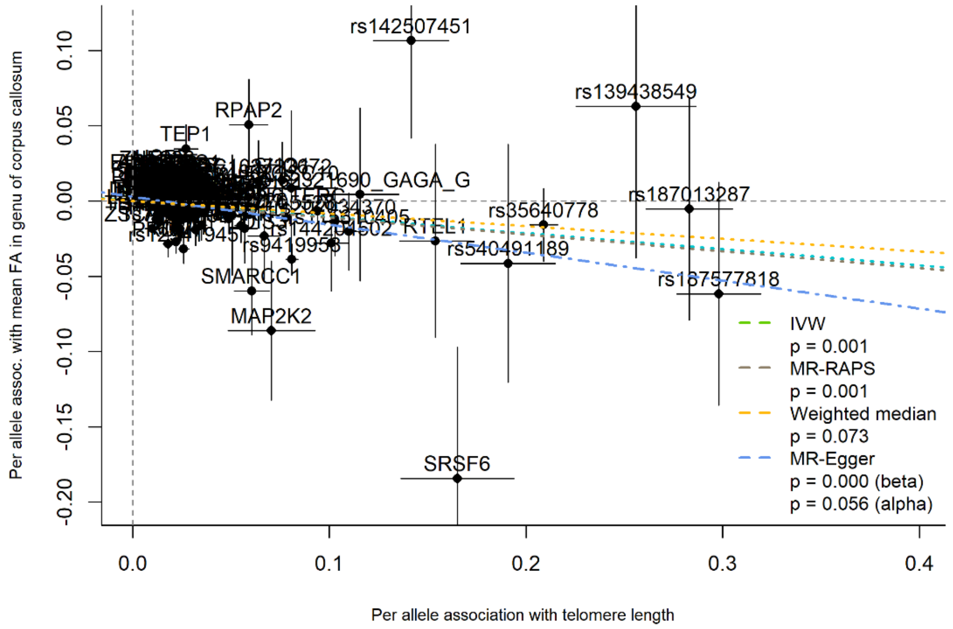


**eFigure 28.** SNP-mean FA in genu of corpus callosum association plotted against SNP-telomere length association, labelled by the mapped gene, with MR slope estimates shown

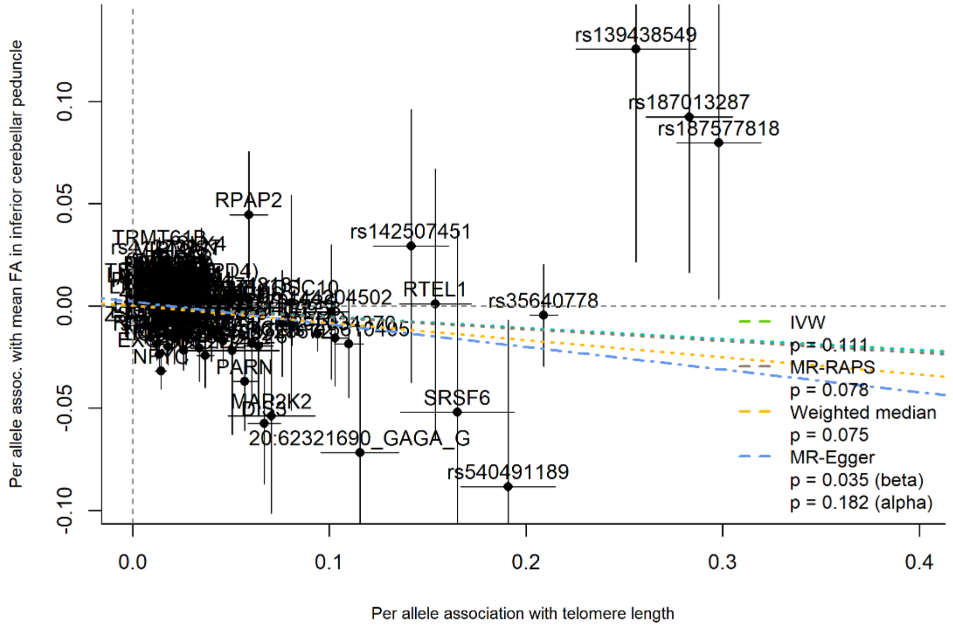


**eFigure 29.** SNP-mean FA in inferior cerebellar peduncle association plotted against SNP-telomere length association, labelled by the mapped gene, with MR slope estimates shown

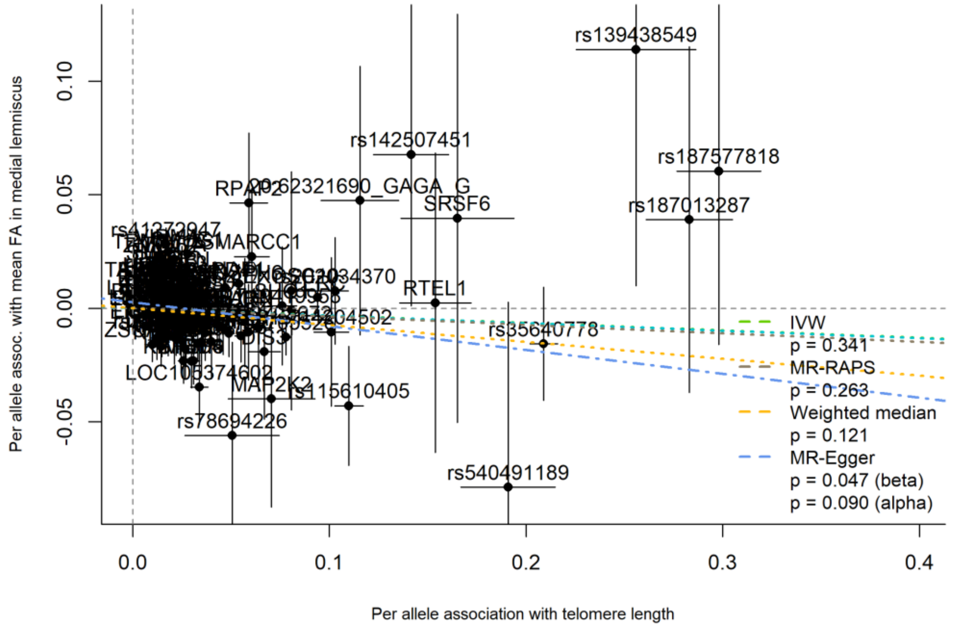


**eFigure 30.** SNP-mean FA in medial lemniscus association plotted against SNP-telomere length association, labelled by the mapped gene, with MR slope estimates shown

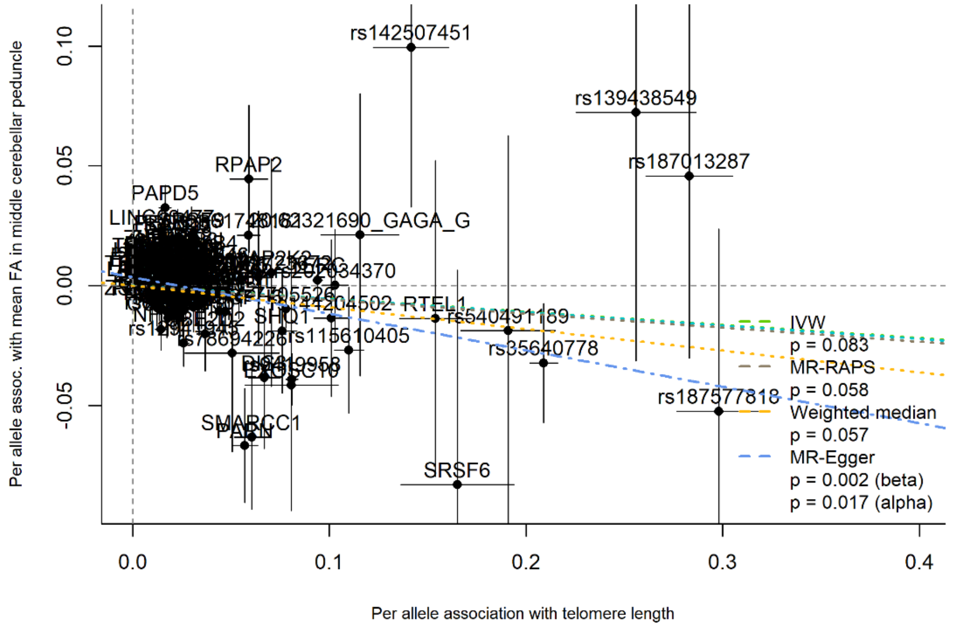


**eFigure 31.** SNP-mean FA in middle cerebellar peduncle association plotted against SNP-telomere length association, labelled by the mapped gene, with MR slope estimates shown

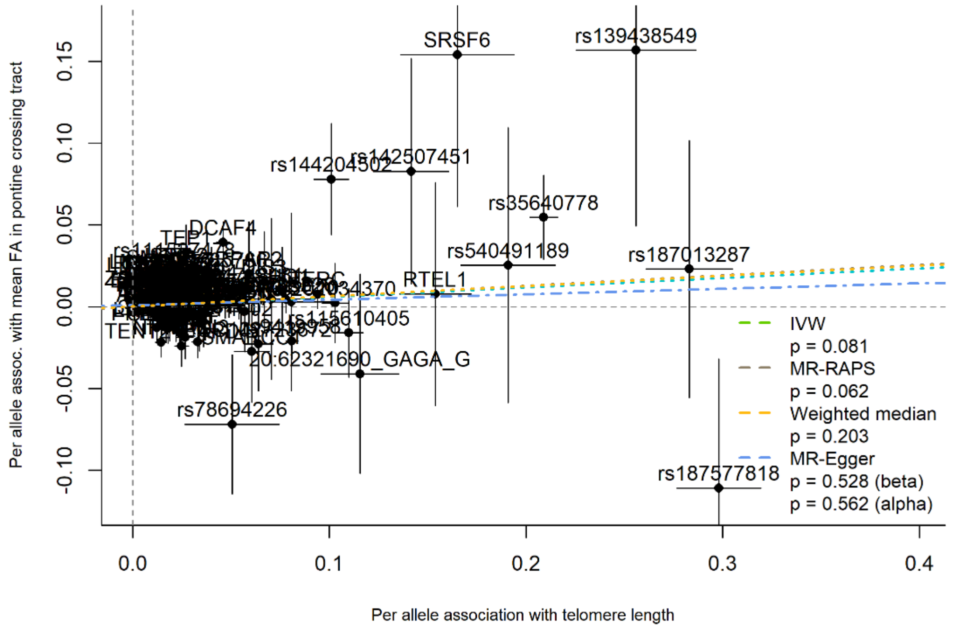


**eFigure 32.** SNP-mean FA in pontine crossing tract association plotted against SNP-telomere length association, labelled by the mapped gene, with MR slope estimates shown

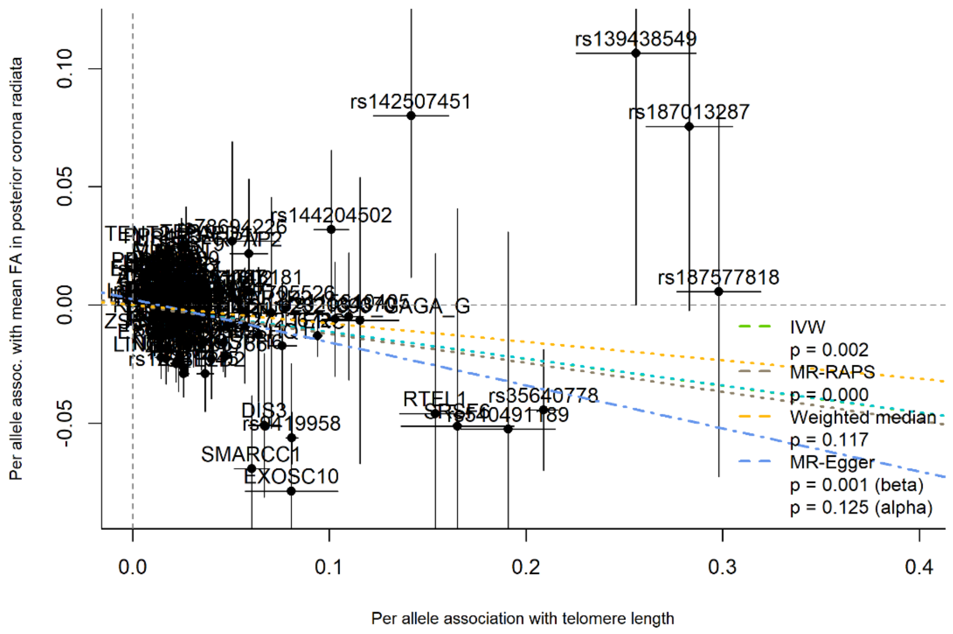


**eFigure 33.** SNP-mean FA in posterior corona radiata association plotted against SNP-telomere length association, labelled by the mapped gene, with MR slope estimates shown

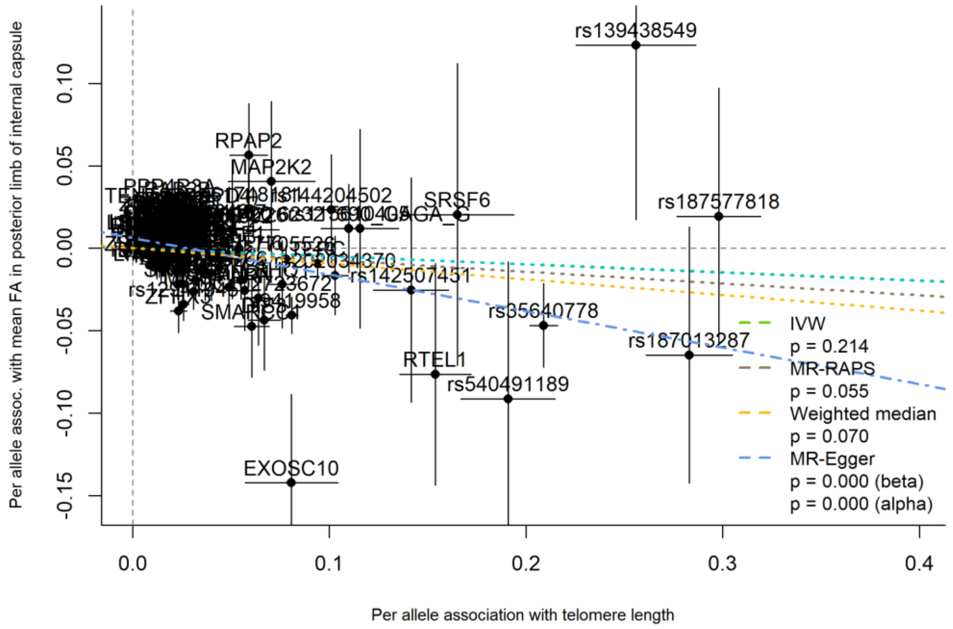


**eFigure 34.** SNP-mean FA in posterior limb of internal capsule association plotted against SNP-telomere length association, labelled by the mapped gene, with MR slope estimates shown

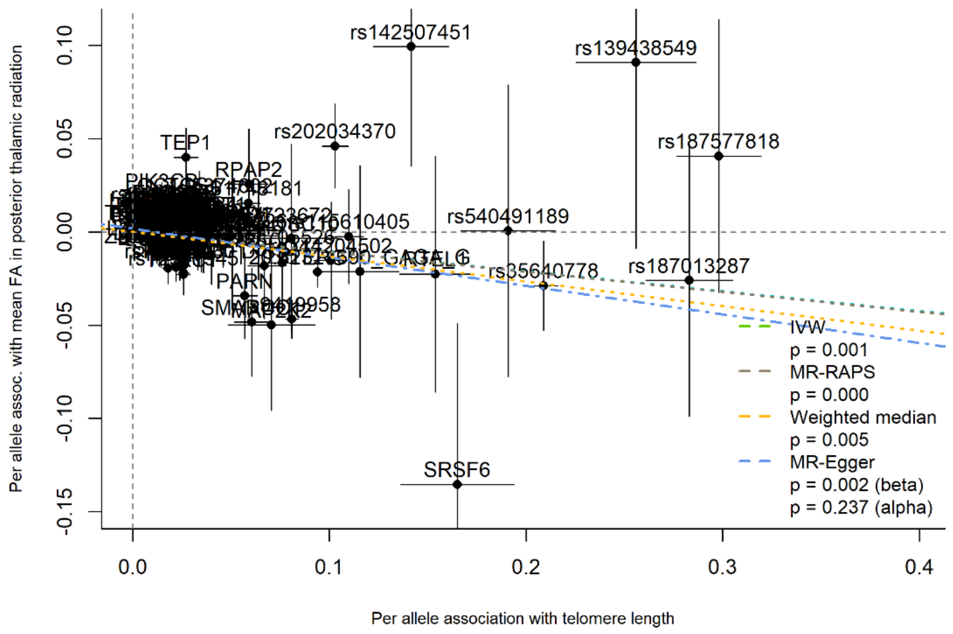


**eFigure 35.** SNP-mean FA in posterior thalamic radiation association plotted against SNP-telomere length association, labelled by the mapped gene, with MR slope estimates shown

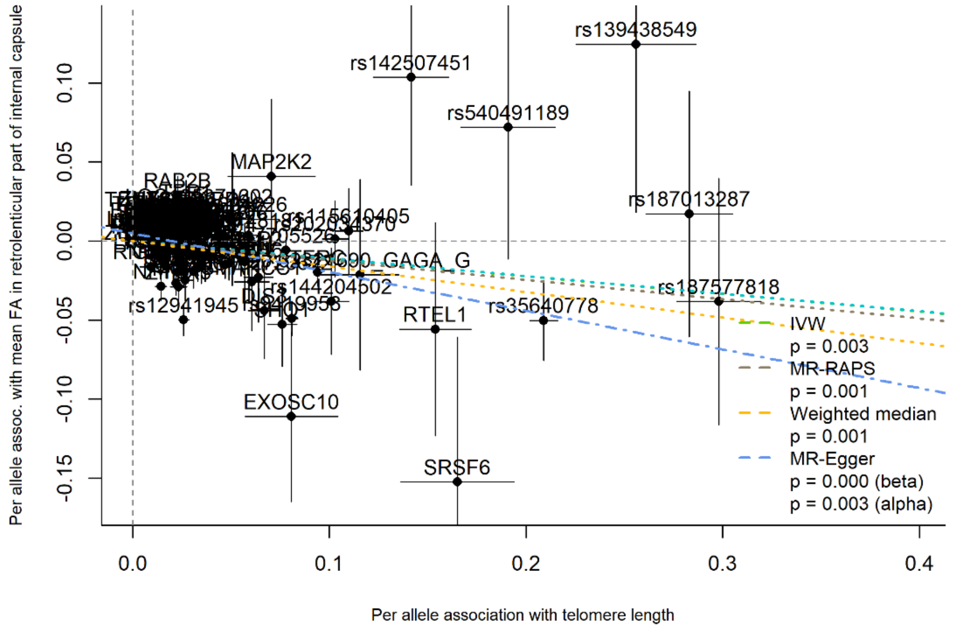


**eFigure 36.** SNP-mean FA in retrolenticular part of internal capsule association plotted against SNP-telomere length association, labelled by the mapped gene, with MR slope estimates shown

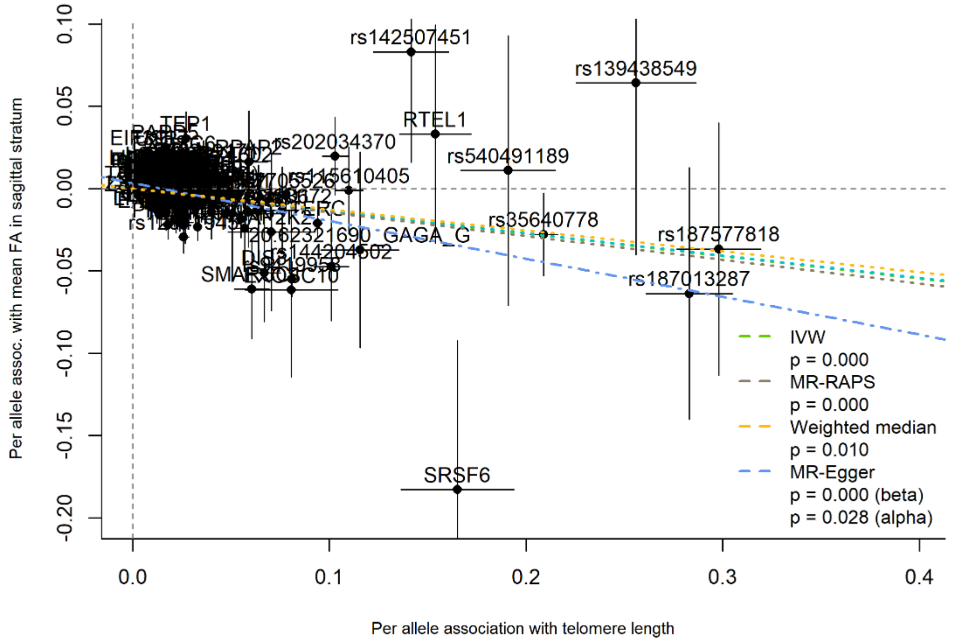


**eFigure 37.** SNP-mean FA in sagittal stratum association plotted against SNP-telomere length association, labelled by the mapped gene, with MR slope estimates shown

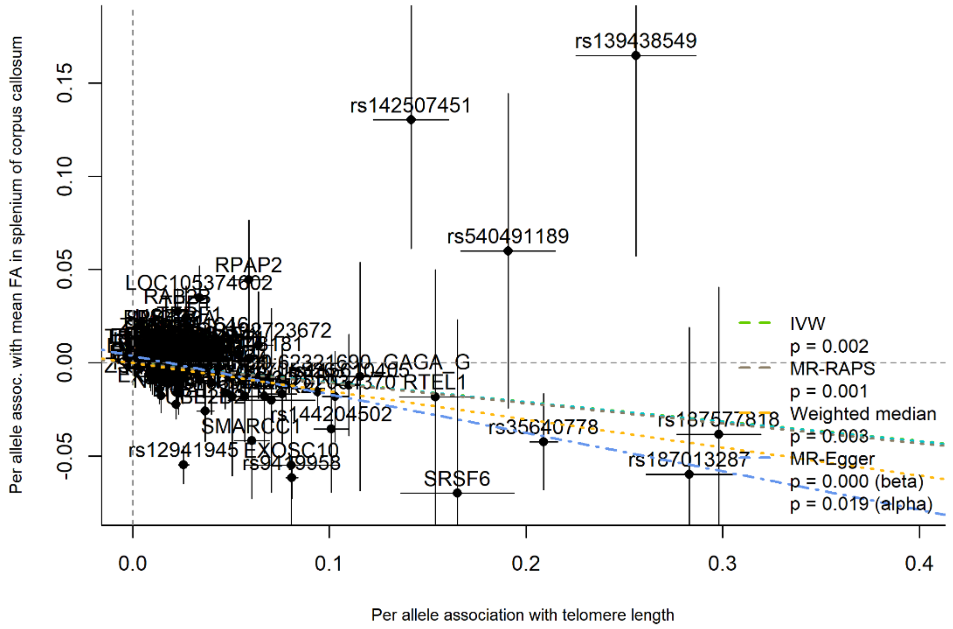


**eFigure 38.** SNP-mean FA in splenium of corpus callosum association plotted against SNP-telomere length association, labelled by the mapped gene, with MR slope estimates shown

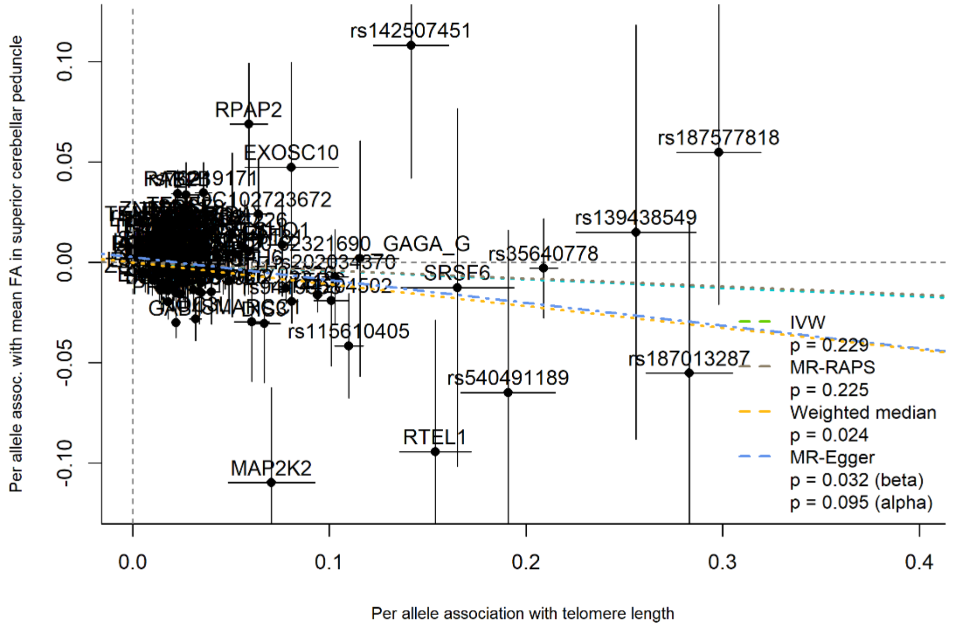


**eFigure 39.** SNP-mean FA in superior cerebellar peduncle association plotted against SNP-telomere length association, labelled by the mapped gene, with MR slope estimates shown

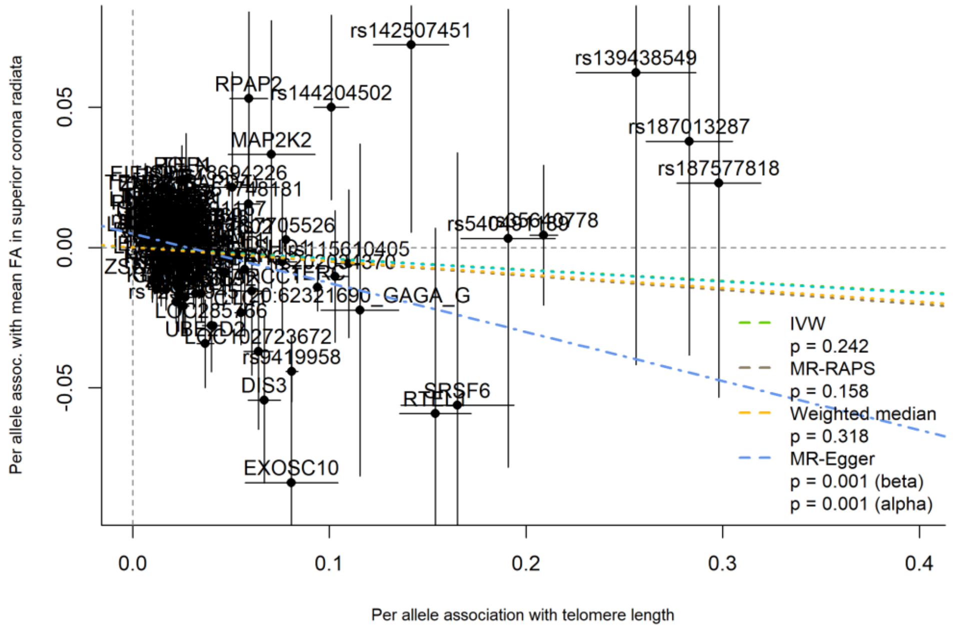


**eFigure 40.** SNP-mean FA in superior corona radiata association plotted against SNP-telomere length association, labelled by the mapped gene, with MR slope estimates shown

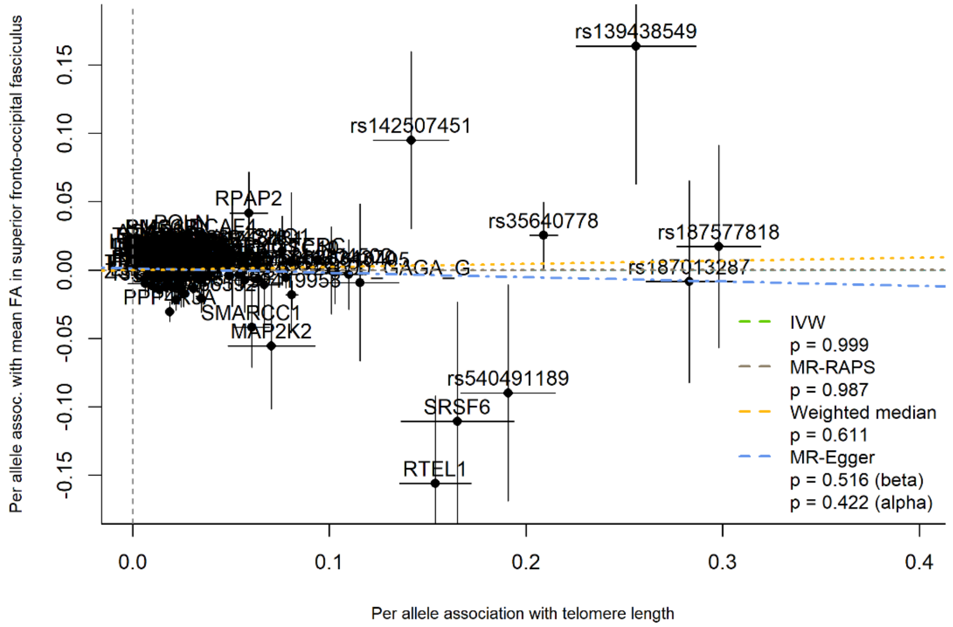


**eFigure 41.** SNP-mean FA in superior fronto-occipital fasciculus association plotted against SNP-telomere length association, labelled by the mapped gene, with MR slope estimates shown

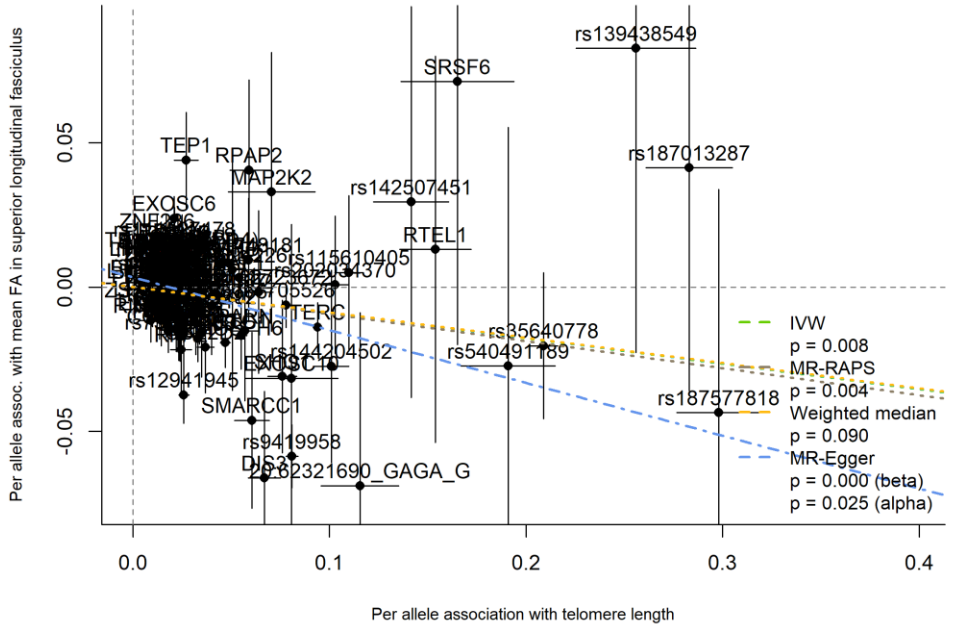


**eFigure 42.** SNP-mean FA in superior longitudinal fasciculus association plotted against SNP-telomere length association, labelled by the mapped gene, with MR slope estimates shown

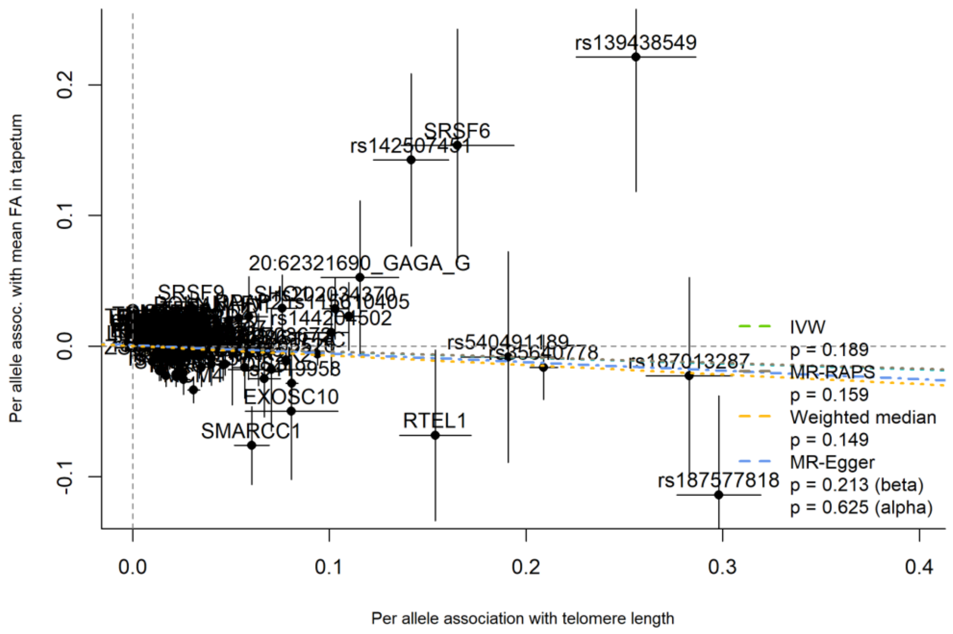


**eFigure 43.** SNP-mean FA in tapetum association plotted against SNP-telomere length association, labelled by the mapped gene, with MR slope estimates shown

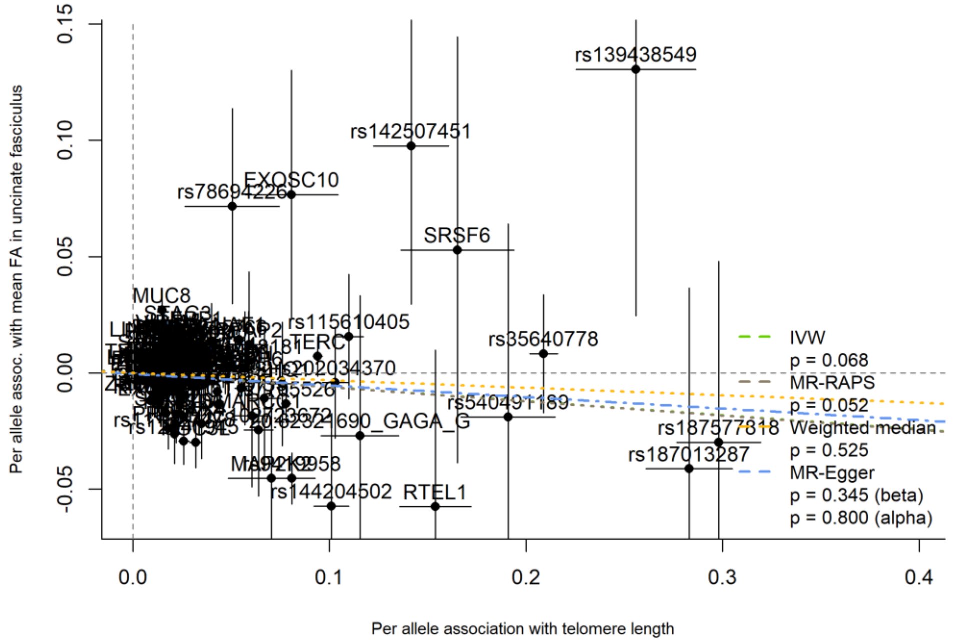


**eFigure 44.** SNP-mean FA in uncinate fasciculus association plotted against SNP-telomere length association, labelled by the mapped gene, with MR slope estimates shown

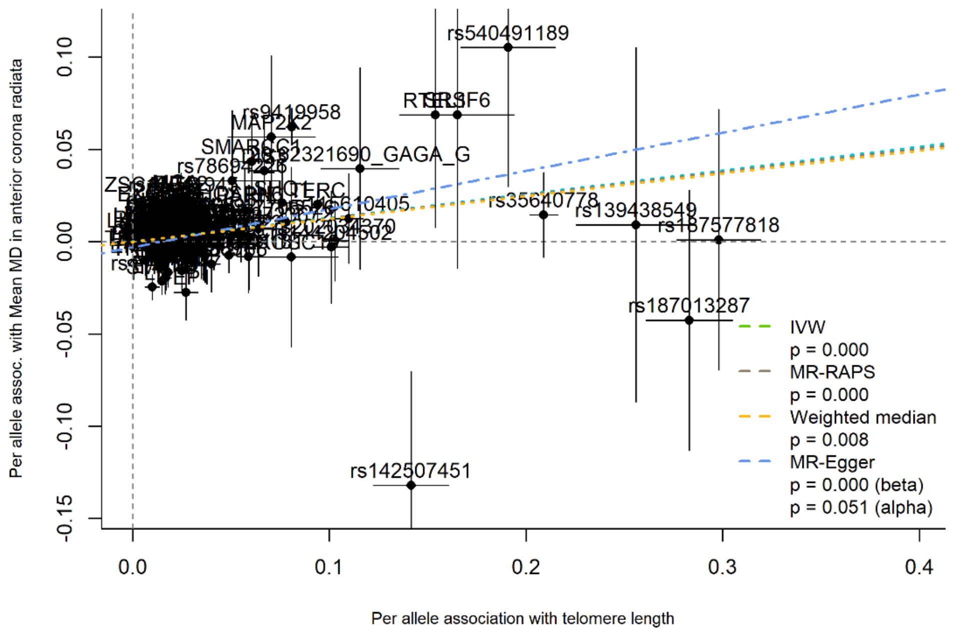


**eFigure 45.** SNP-mean MD in anterior corona radiata association plotted against SNP-telomere length association, labelled by the mapped gene, with MR slope estimates shown

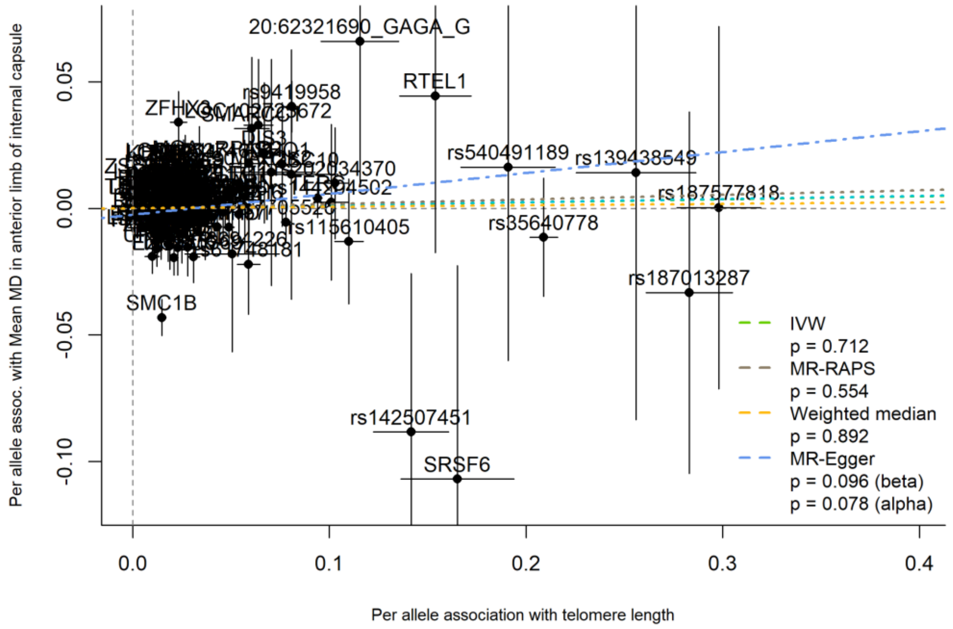


**eFigure 46.** SNP-mean MD in anterior limb of internal capsule association plotted against SNP-telomere length association, labelled by the mapped gene, with MR slope estimates shown

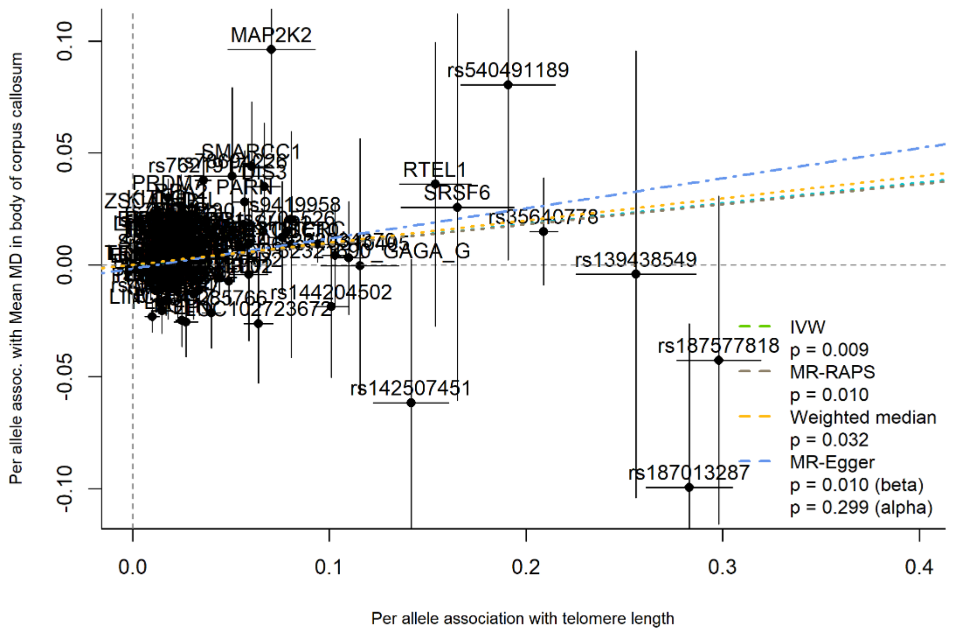


**eFigure 47.** SNP-mean MD in body of corpus callosum association plotted against SNP-telomere length association, labelled by the mapped gene, with MR slope estimates shown

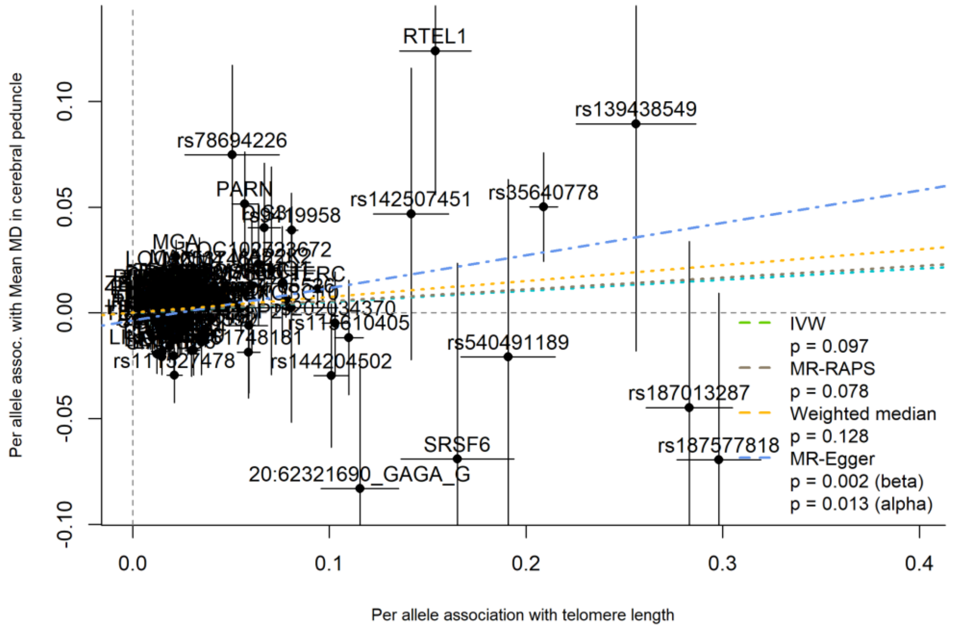


**eFigure 48.** SNP-mean MD in cerebral peduncle association plotted against SNP-telomere length association, labelled by the mapped gene, with MR slope estimates shown

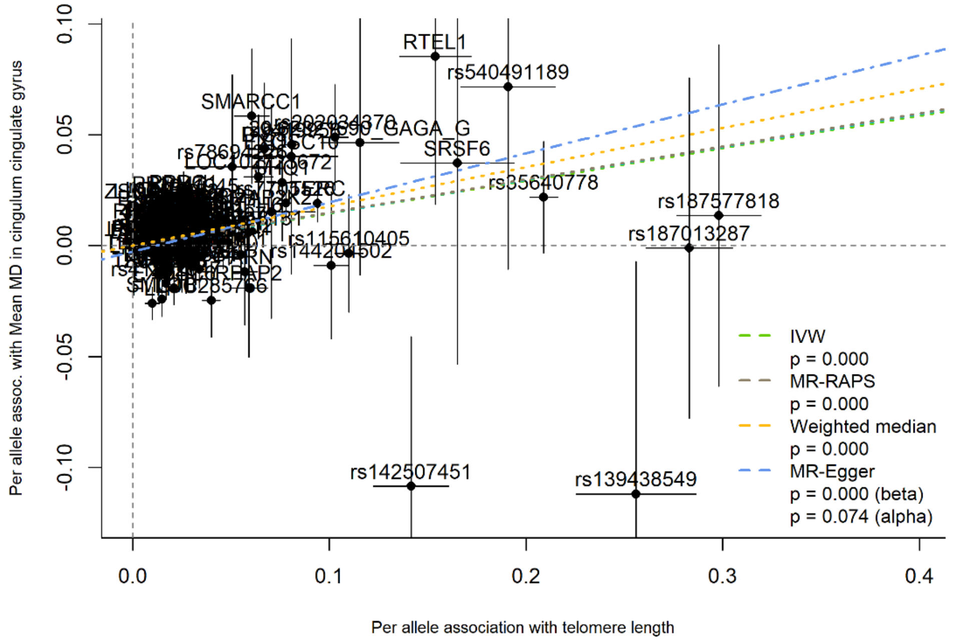


**eFigure 49.** SNP-mean MD in cingulum cingulate gyrus association plotted against SNP-telomere length association, labelled by the mapped gene, with MR slope estimates shown

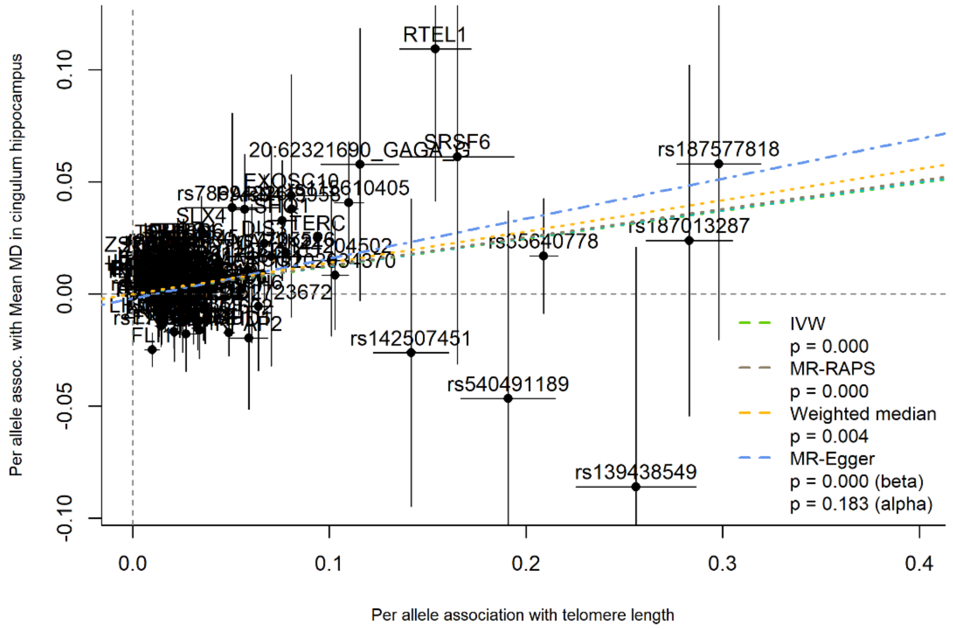


**eFigure 50.** SNP-mean MD in cingulum hippocampus association plotted against SNP-telomere length association, labelled by the mapped gene, with MR slope estimates shown

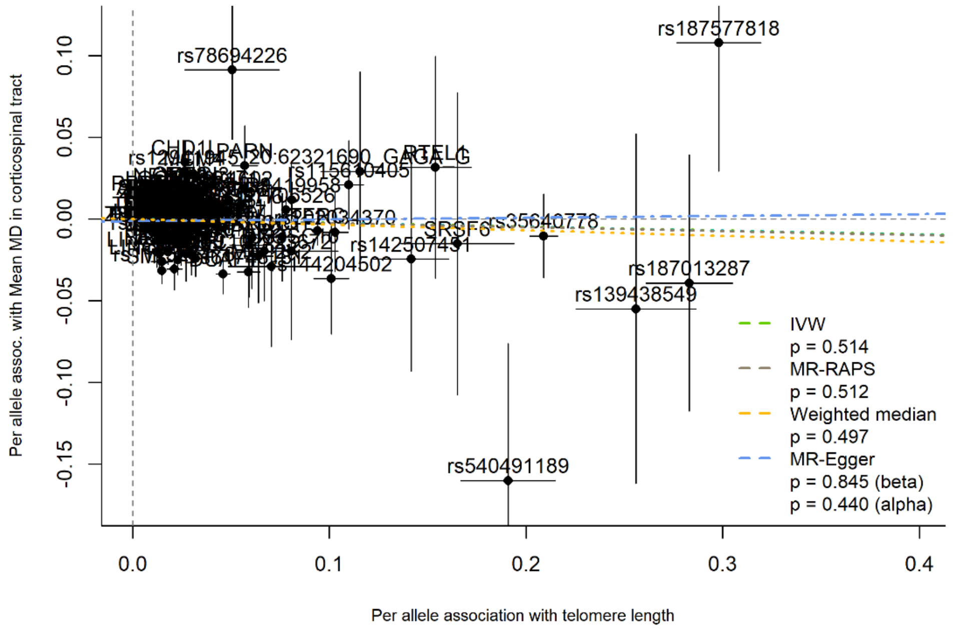


**eFigure 51.** SNP-mean MD in corticospinal tract association plotted against SNP-telomere length association, labelled by the mapped gene, with MR slope estimates shown

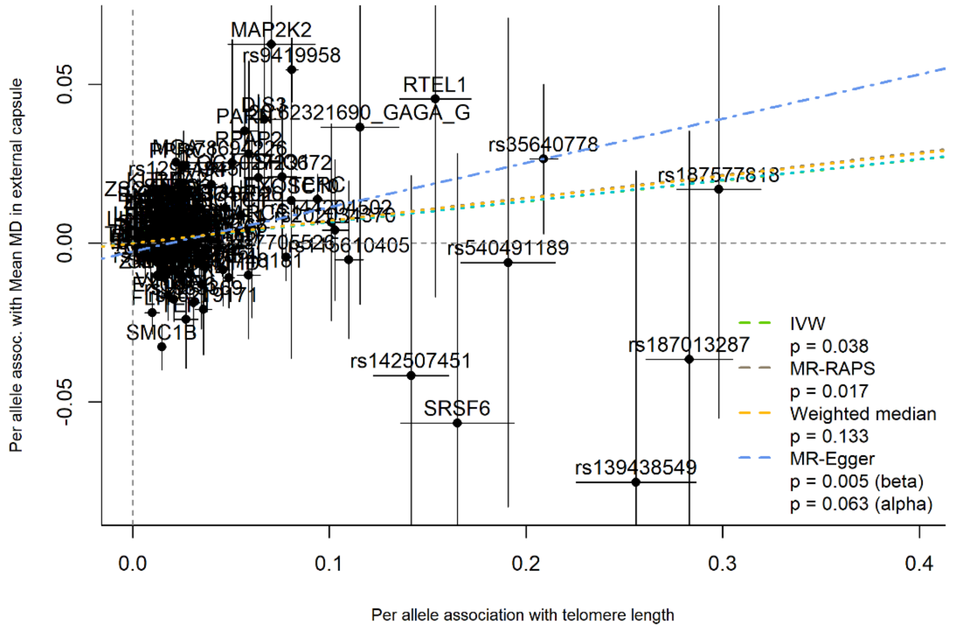


**eFigure 52.** SNP-mean MD in external capsule association plotted against SNP-telomere length association, labelled by the mapped gene, with MR slope estimates shown

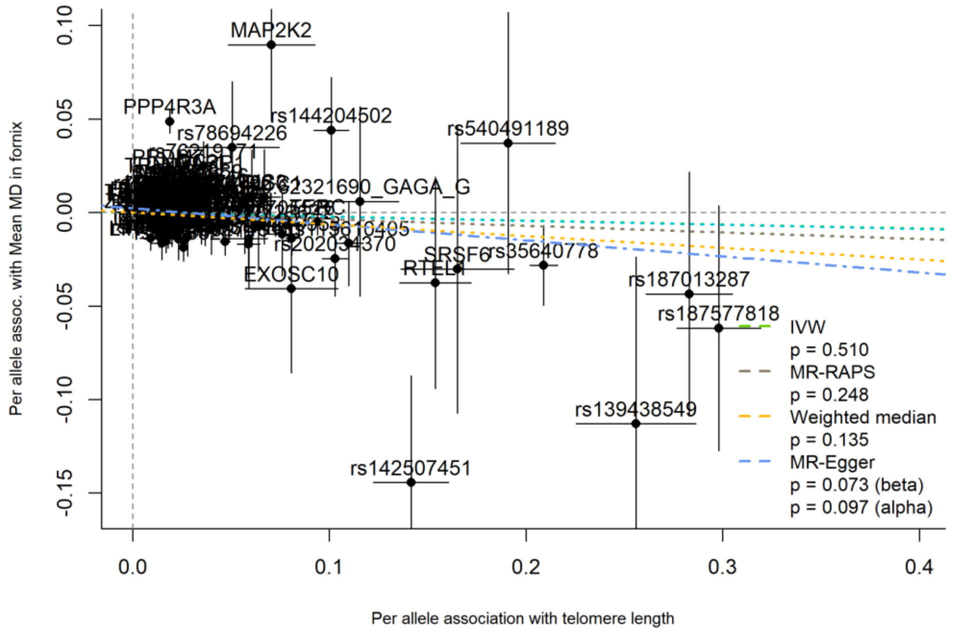


**eFigure 53.** SNP-mean MD in fornix association plotted against SNP-telomere length association, labelled by the mapped gene, with MR slope estimates shown

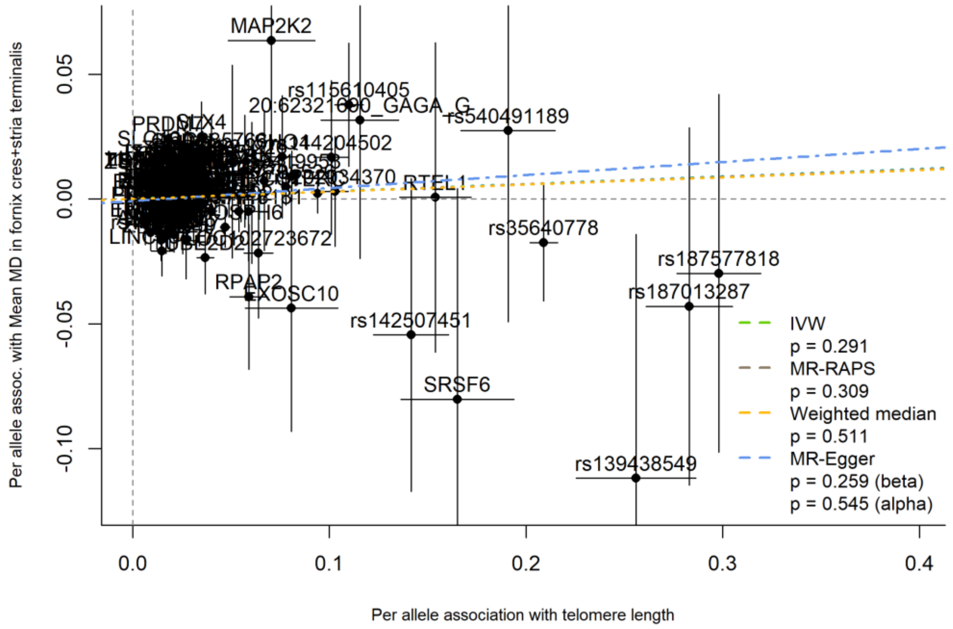


**eFigure 54.** SNP-mean MD in fornix cres+stria terminalis association plotted against SNP-telomere length association, labelled by the mapped gene, with MR slope estimates shown

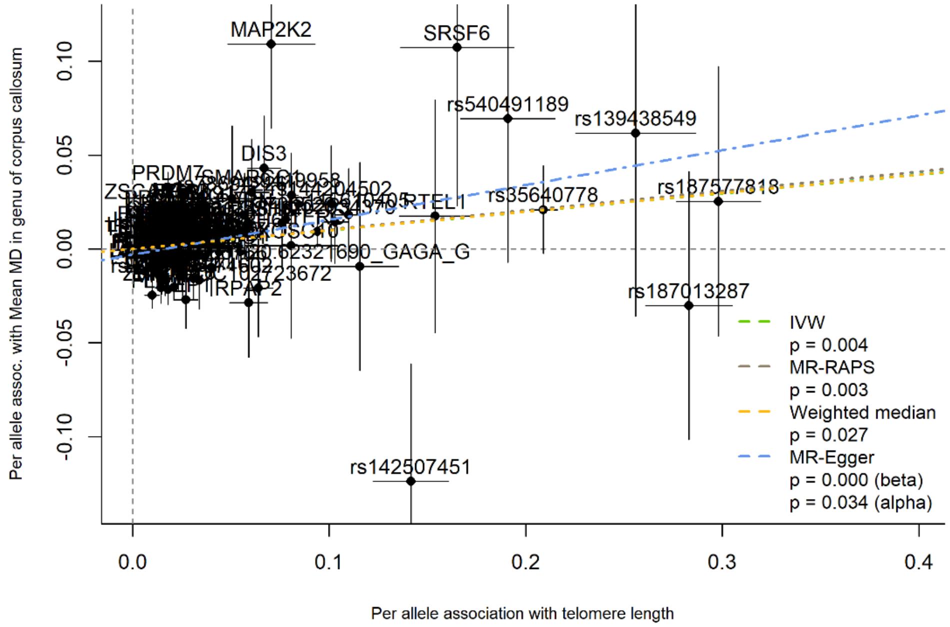


**eFigure 55.** SNP-mean MD in genu of corpus callosum association plotted against SNP-telomere length association, labelled by the mapped gene, with MR slope estimates shown

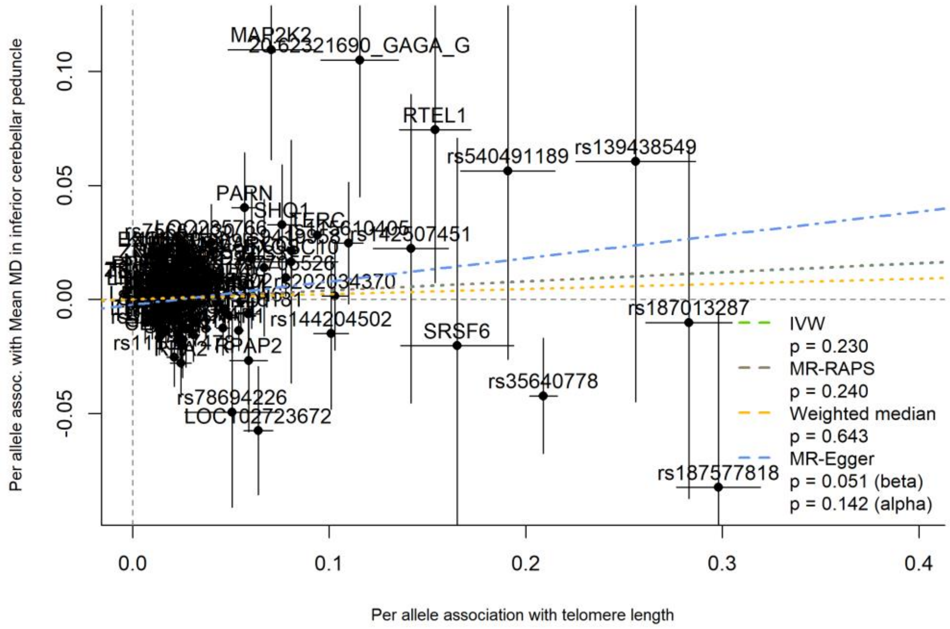


**eFigure 56.** SNP-mean MD in inferior cerebellar peduncle association plotted against SNP-telomere length association, labelled by the mapped gene, with MR slope estimates shown

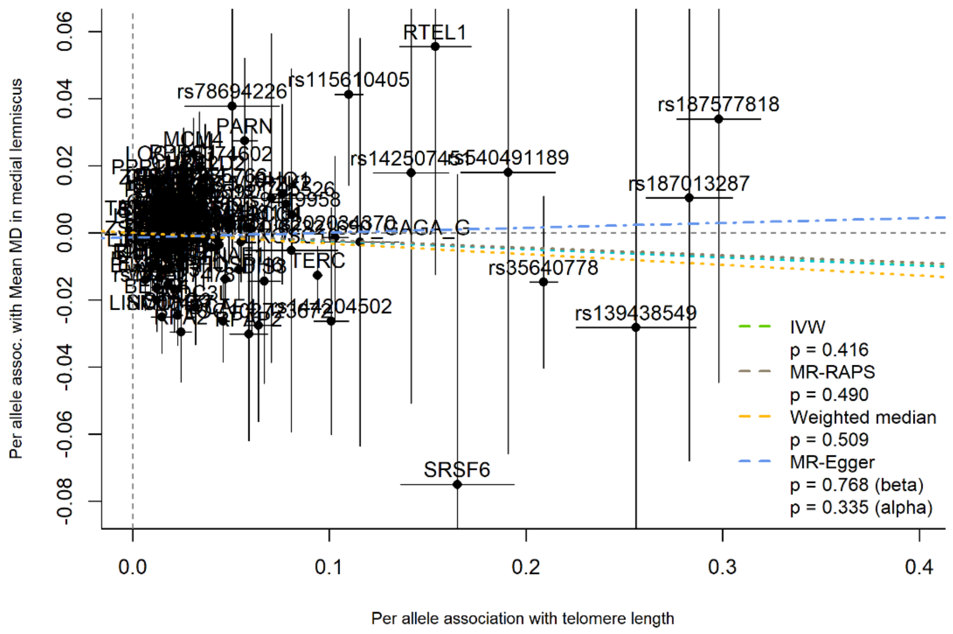


**eFigure 57.** SNP-mean MD in medial lemniscus association plotted against SNP-telomere length association, labelled by the mapped gene, with MR slope estimates shown

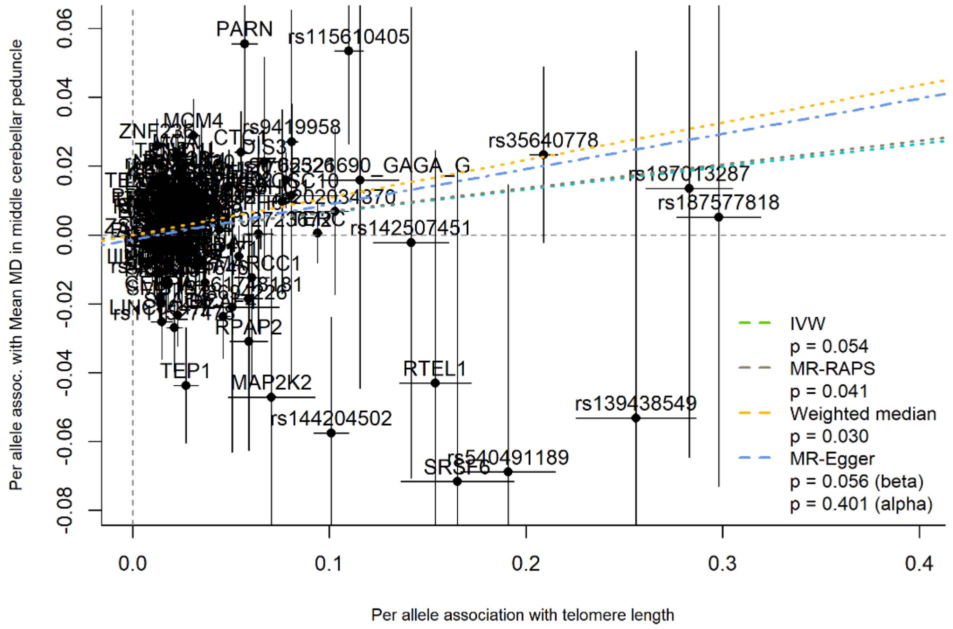


**eFigure 58.** SNP-mean MD in middle cerebellar peduncle association plotted against SNP-telomere length association, labelled by the mapped gene, with MR slope estimates shown

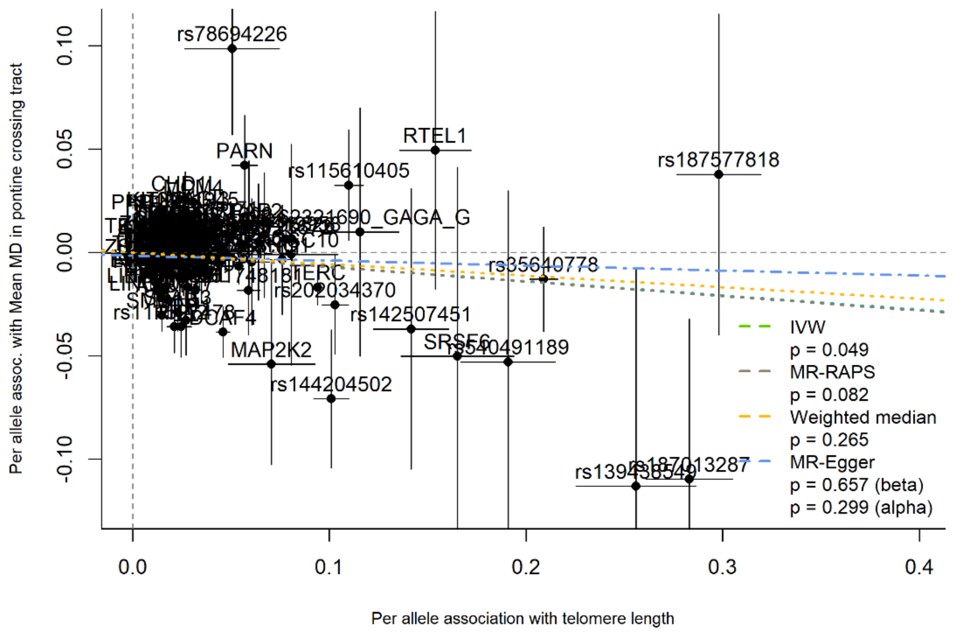


**eFigure 59.** SNP-mean MD in pontine crossing tract association plotted against SNP-telomere length association, labelled by the mapped gene, with MR slope estimates shown

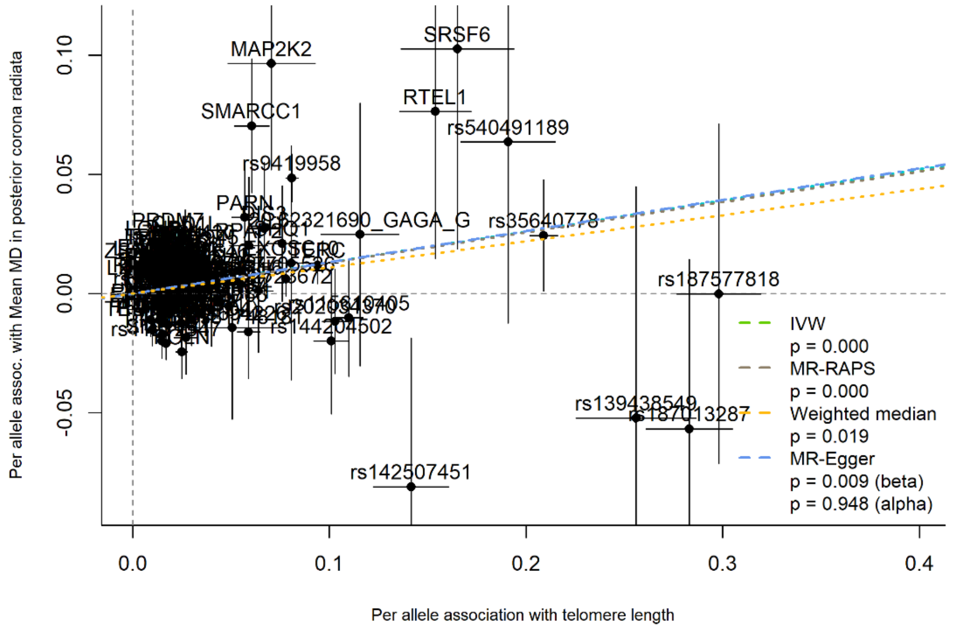


**eFigure 60.** SNP-mean MD in posterior corona radiata association plotted against SNP-telomere length association, labelled by the mapped gene, with MR slope estimates shown

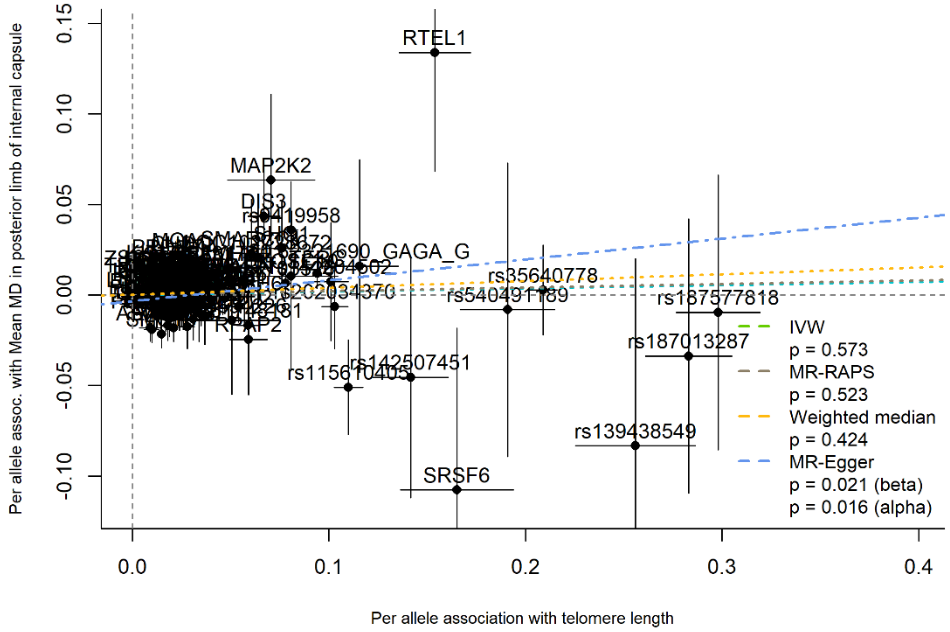


**eFigure 61.** SNP-mean MD in posterior limb of internal capsule association plotted against SNP-telomere length association, labelled by the mapped gene, with MR slope estimates shown

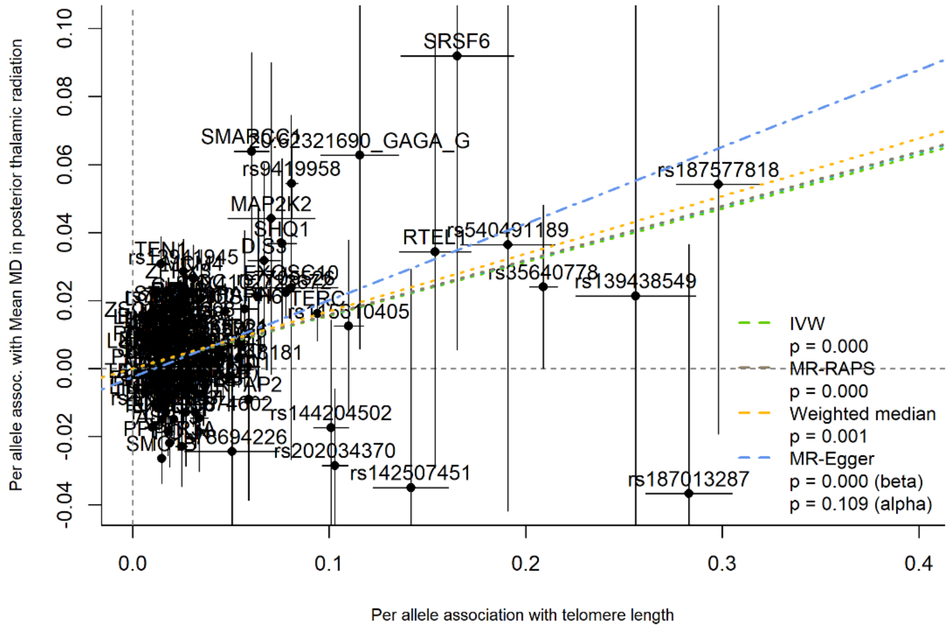


**eFigure 62.** SNP-mean MD in posterior thalamic radiation association plotted against SNP-telomere length association, labelled by the mapped gene, with MR slope estimates shown

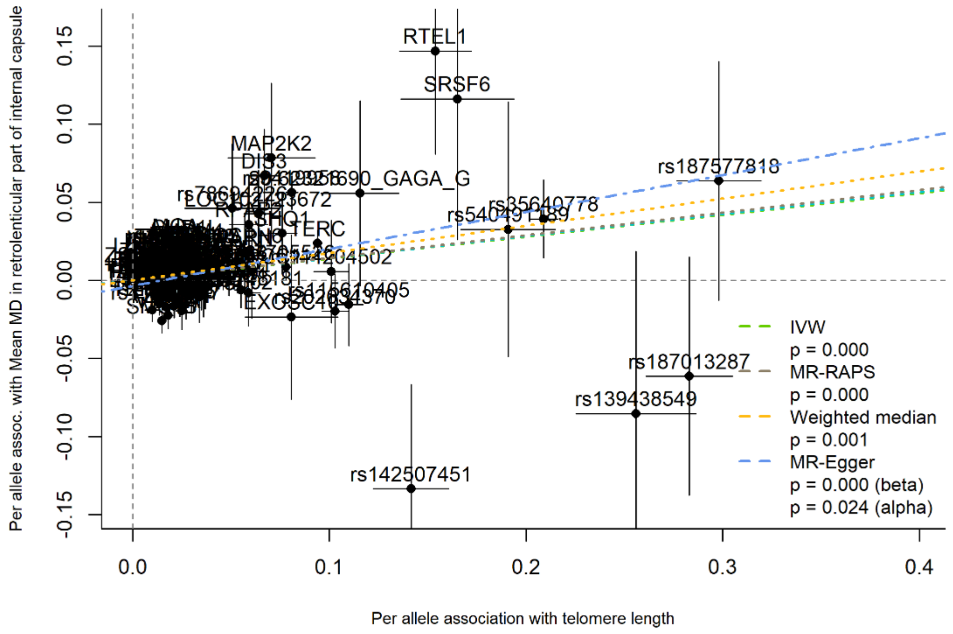


**eFigure 63.** SNP-mean MD in retrolenticular part of internal capsule association plotted against SNP-telomere length association, labelled by the mapped gene, with MR slope estimates shown

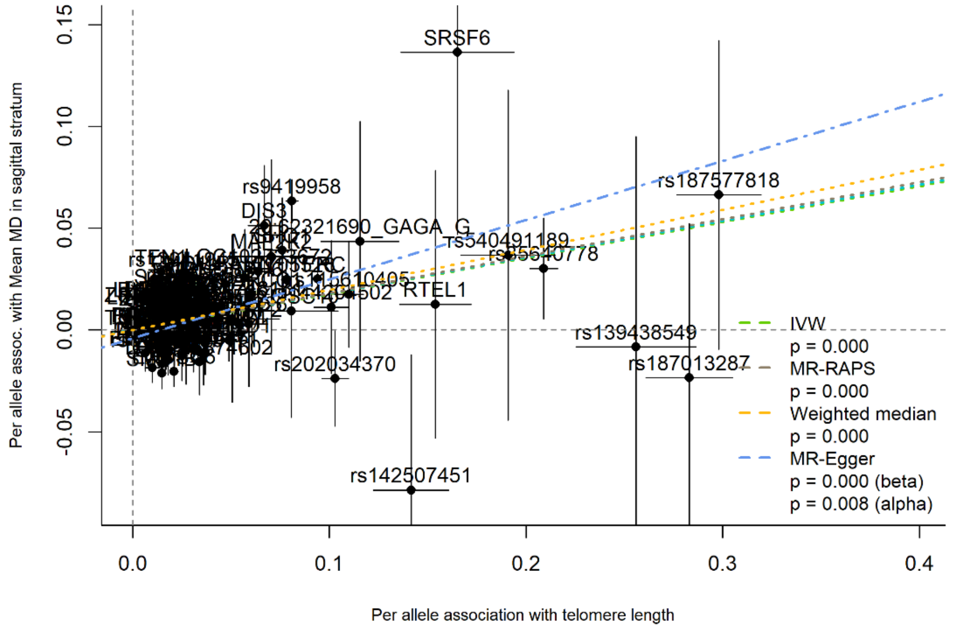


**eFigure 64.** SNP-mean MD in sagittal stratum association plotted against SNP-telomere length association, labelled by the mapped gene, with MR slope estimates shown

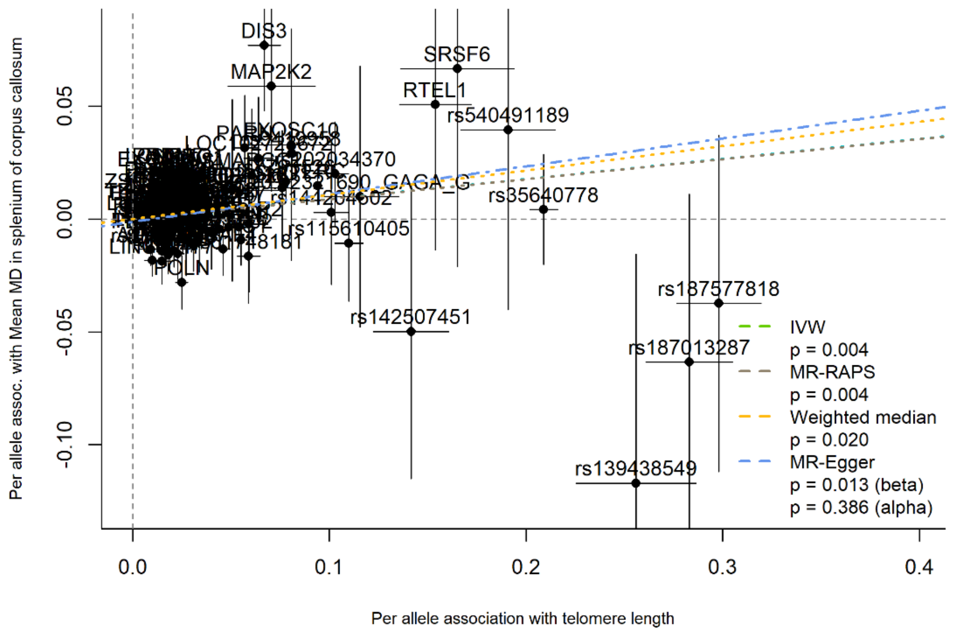


**eFigure 65.** SNP-mean MD in splenium of corpus callosum association plotted against SNP-telomere length association, labelled by the mapped gene, with MR slope estimates shown

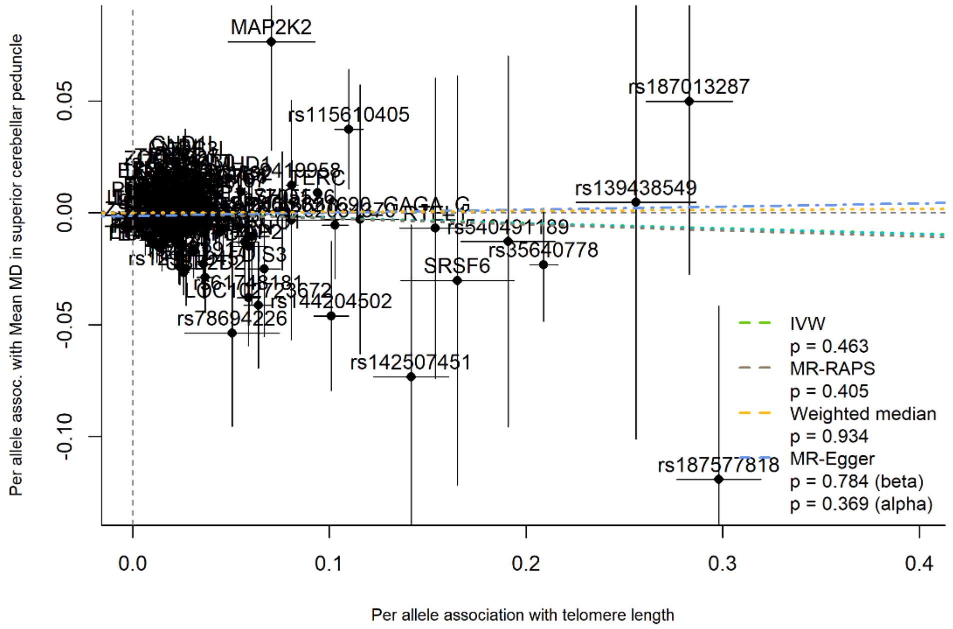


**eFigure 66.** SNP-mean MD in superior cerebellar peduncle association plotted against SNP-telomere length association, labelled by the mapped gene, with MR slope estimates shown

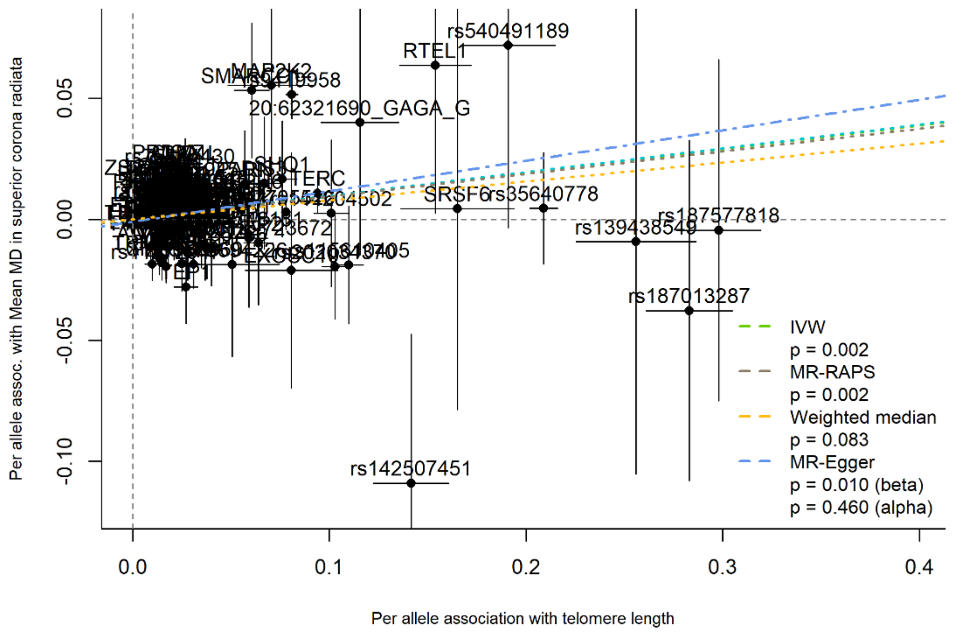


**eFigure 67.** SNP-mean MD in superior corona radiata association plotted against SNP-telomere length association, labelled by the mapped gene, with MR slope estimates shown

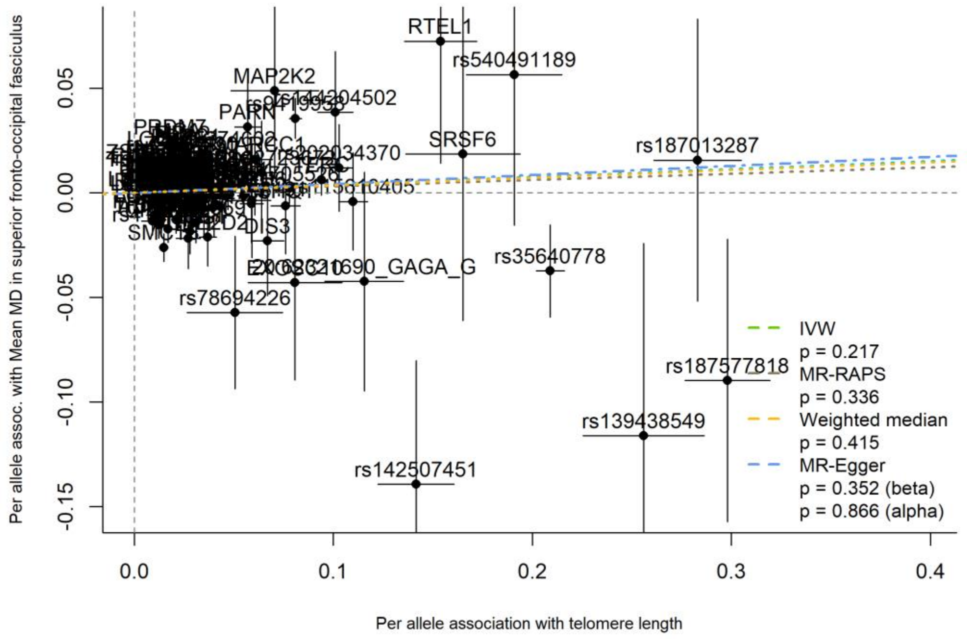


**eFigure 68.** SNP-mean MD in superior fronto-occipital fasciculus association plotted against SNP-telomere length association, labelled by the mapped gene, with MR slope estimates shown

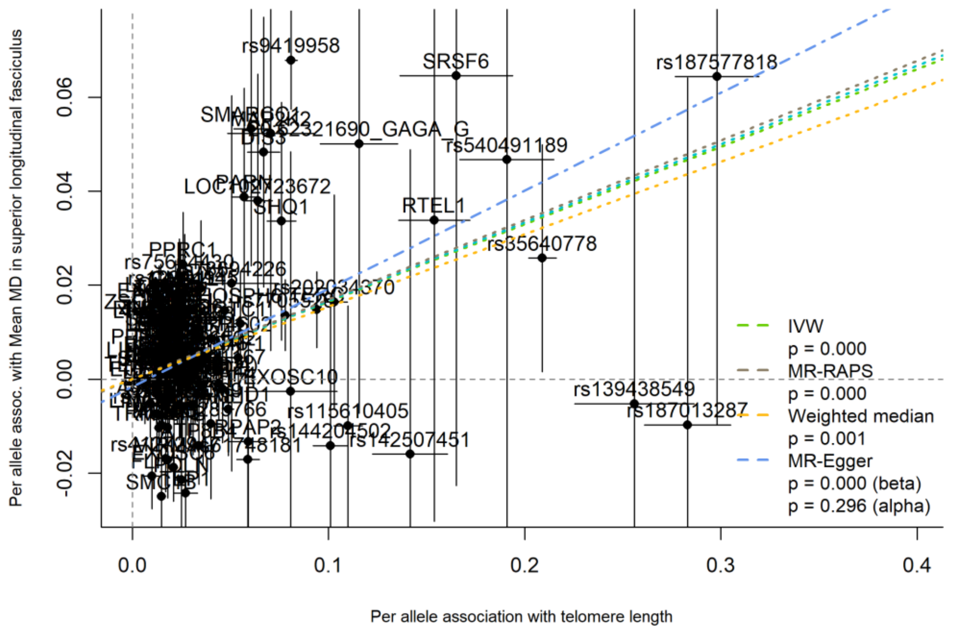


**eFigure 69.** SNP-mean MD in superior longitudinal fasciculus association plotted against SNP-telomere length association, labelled by the mapped gene, with MR slope estimates shown

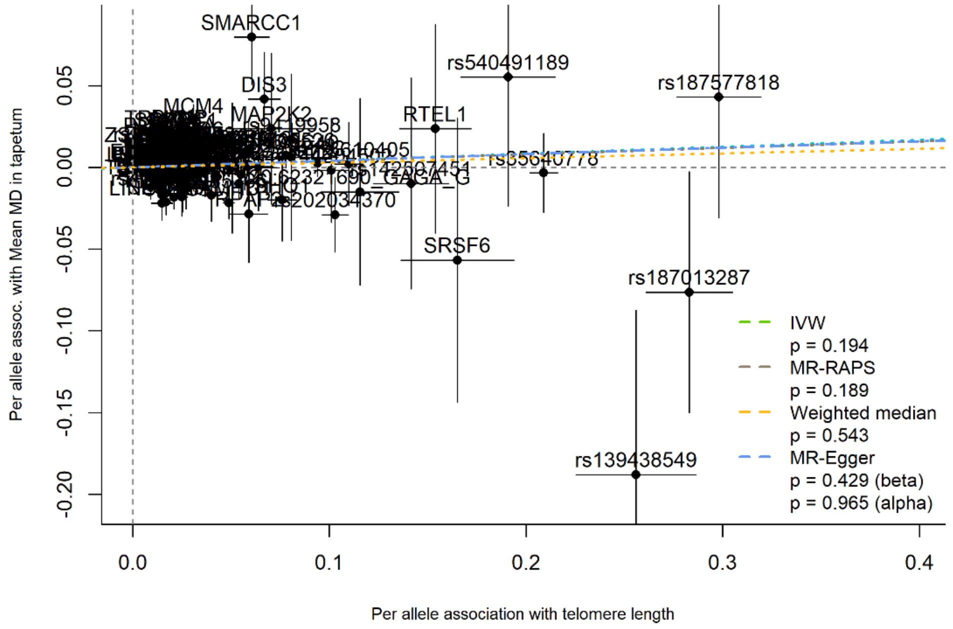


**eFigure 70.** SNP-mean MD in tapetum association plotted against SNP-telomere length association, labelled by the mapped gene, with MR slope estimates shown

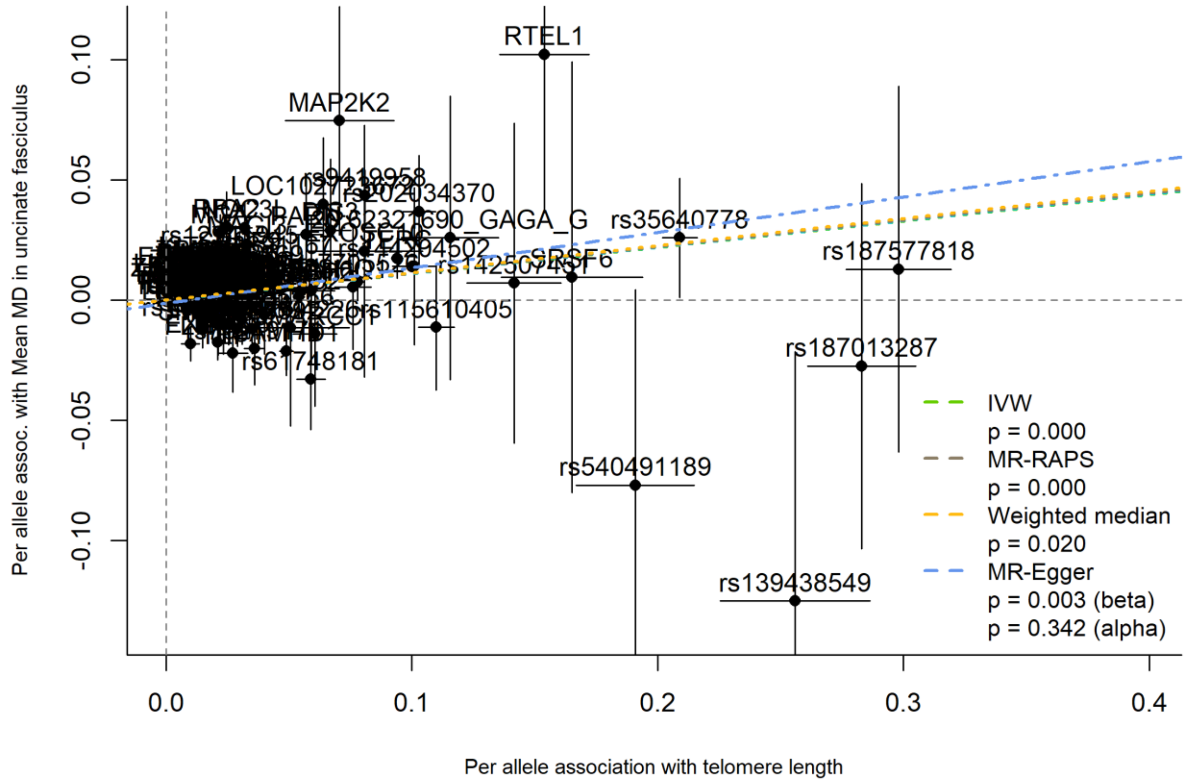


**eFigure 71.** SNP-mean MD in uncinate fasciculus association plotted against SNP-telomere length association, labelled by the mapped gene, with MR slope estimates shown
